# Benchmarking local genetic correlation estimation methods using summary statistics from genome-wide association studies

**DOI:** 10.1101/2023.06.01.23290835

**Authors:** Chi Zhang, Yiliang Zhang, Yunxuan Zhang, Hongyu Zhao

## Abstract

Local genetic correlation evaluates the correlation of genetic effects between different traits across genetic variants in a local region. It has been proven informative for understanding the genetic similarities of complex traits beyond that captured by global genetic correlation calculated across the whole genome. Several summary-statistics-based approaches have been developed for estimating local genetic correlation, including ***ρ***-hess, SUPERGNOVA, and LAVA. However, there has not been a comprehensive evaluation of these methods to offer practical guidelines on the choices of these methods. In this study, we conduct benchmark comparisons of the performance of these three methods through extensive simulation and real data analyses. We focus on two technical difficulties in estimating local genetic correlation: sample overlaps across traits and local linkage disequilibrium (LD) estimates when only the external reference panels are available. Our simulations suggest that the type-I error and estimation accuracy are highly dependent on the estimation of the local LD matrix. These observations are corroborated by real data analyses of 31 complex traits. Overall, our results offer insights into post-GWAS local correlation studies and highlight issues that demand future methodology developments.

## 1 Introduction

In recent years, genome-wide association analyses (GWAS) have identified tens of thousands of genetic variants associated with numerous complex traits and diseases[1–4]. Various post-GWAS approaches[5], such as fine mapping, genetic correlation, functional enrichment, and polygenic risk score (PRS), are routinely conducted to gain a further understanding of the genetic variants and biological mechanisms behind the observed statistical associations. Estimating genetic correlation using GWAS data is an essential part of the post-GWAS analysis that can quantify genetic similarities and uncover shared genetic basis of complex traits and disorders. Additionally, genetic correlation results can help increase statistical power in genetic association studies [6, 7], and improve polygenic risk score prediction accuracy [8–12]. Genetic correlations can be characterized globally, summarizing the average correlation of genetic effects across the genome, or locally highlighting specific regions having correlated effects on different traits.

While global genetic correlations have been extensively studied [13] with many methods developed for their estimations using GWAS data [14–17], they may not capture local genetic correlations that may be distinct across different regions [18–22]. These include the existence of opposing correlations in different regions, which can lead to a non-significant global genetic correlation. Furthermore, global genetic correlations provide limited insight into the shared biological mechanisms, when different genomic regions have different correlation levels. For example, when investigating shared genetic architecture between COVID-19 severity and idiopathic pulmonary fibrosis (IPF), the global genetic correlation between these two diseases was 0.35 (p = 0.001), however, the effect of MUC5B and ATP11A revealed opposing effects for these two diseases[23]. In order to capture local correlation patterns, several methods have been developed for estimating or detecting local genetic correlation including *ρ*-HESS[24], SUPERGNOVA[20], and LAVA[21].

*ρ*-hess[19] and SUPERGNOVA[20] focus on evaluating bivariate local genetic correlations, whereas LAVA[21] used partial correlation and multiple regression to estimate bivariate and multivariate genetic correlations. These methods also differ in model assumptions relating genetic variants to their effects on traits. Whereas *ρ*-hess[19] and LAVA[21] are based on fixed effects models, SUPERGNOVA[20] is based on a random effects model. Although there was earlier research evaluating the performance of these approaches[20, 21], there has not been a comprehensive study of their performance through extensive simulation and real data analyses. Given the importance of inferring local genetic correlations, there is a need for objectively benchmarking the performance of these three methods with user-defined genome partitions in realistic settings. As all three methods both infer shared genetic effects and estimate local heritability in a local region, we evaluate their performances for both tasks, i.e. local genetic covariance/correlation estimation between two traits and local heritability estimation.

We conducted simulations using the observed genotype data from the UK Biobank (UKB)[25] and compared different methods using both in-sample and external reference panels to estimate local linkage disequilibrium (LD) structure. We used genotype data from 1KG Phase 3[26] as the external reference panel. We considered binary and continuous traits with varying sample overlaps and region sizes. We assessed the robustness of each method against both infinitesimal and non-infinitesimal models, and whether the effect sizes follow the underlying assumption of the random effect method, SUPERGNOVA. Additionally, we investigated the stability of *ρ*-hess and LAVA with different reference panels. After simulations, we applied these methods to analyze 31 complex traits with publicly available GWAS summary statistics. To validate the accuracy of these methods in real data, we applied LDSC[14] to estimate global genetic covariances and heritability and compared these estimates with the sum of local heritability and local genetic covariance. For these real data, we also assessed the stability of the point estimates and inferences using different reference panels and conducted polygenic risk score analyses using markers from regions with significant positive and negative correlations. The observations from our simulation and real data analyses offer valuable insights into the statistical properties, advantages, and limitations of each method.

## 2 Methods

### 2.1 Study population and quality control of genotype data

The UKB is a large, prospective study that aims to examine complex traits and diseases in middle-aged adults. We performed simulations using imputed genotype data from UKB and selected samples from genetically unrelated participants of White British ancestry (n=276,731). For real data analysis, we used phenotype and genotype data from UKB to perform polygenic risk score (PRS) analysis for four traits: coronary artery disease (CAD), type 2 diabetes (T2D), low-density lipoprotein (LDL), and body mass index (BMI). Of the participants included in the analysis, 4,765 individuals were diagnosed with CAD, and 40,361 individuals were diagnosed with T2D. The mean LDL level was 3.37 (*mmol/L*) with a standard deviation of 1 and the mean BMI was 31.23 (*kg/m*^2^) with a standard deviation of 5.8.

For real data analysis, in addition to the UKB dataset, we also used data from SPARK (Simons Foundation Powering Autism Research)[27] for autism spectrum disorder (ASD) patients. We accessed the first release of the combined multi-batch SPARK WES dataset, which includes phenotype data for the SPARK Collection Version 7. The details of these samples are available on the SFARI website, https://www.sfari.org/resource/spark/. This dataset includes 69,592 samples processed on the Illumina Global Screening Array and is provided in PLINK[28] format. After removing samples with estimated ancestry other than European (EUR) and missing genotype data, 51,658 samples remained for further analysis. We applied pre-imputation quality control using PLINK[28], including the removal of SNPs with low genotype call rates (<0.95), minor allele frequencies (<0.01), or deviations from Hardy-Weinberg equilibrium(<1e-06), as well as samples with high missing genotype rates (>0.05). This left us with 455,444 SNPs and 43,891 samples. The genotype data were then phased and imputed to the HRC reference panel using the Michigan Imputation Server[29]. After imputation, we applied additional quality control, including the removal of SNPs with low imputation quality (<0.8) or minor allele frequency (<0.01). Finally, the SPARK study data contained 7,194,844 SNPs on the GRCh37/hg19 build, of which 5948,083 SNPs were also included in the EUR 1KG Phase 3 data. We then retained 12,264 individuals who were ASD probands and also had intelligence quotient(IQ) scores to assess the association between PRSs and IQ scores in ASD probands. In our analysis, we quantified cognitive performance using full-scale IQ, verbal IQ, and non-verbal IQ. There were 1,026, 785, and 830 ASD probands in SPARK, who had both these IQ scores and qualify-controlled genotype data, respectively.

### 2.2 Genome partition

Both *ρ*-hess and SUPERGNOVA use LDetect[30] to partition the genome into non-overlapping blocks with an average width of 1.6 cM per block. However, LDetect is a heuristic method that may not always produce optimal results. In contrast, LAVA divides the genome by recursively splitting the largest block into two smaller blocks, selecting a new breakpoint that minimizes local LD between the resulting blocks. To compare the performance of different partitions fairly, we used snp ldsplit[31] to partition the genome, which uses dynamic programming to minimize the sum of squared correlations between variants in different blocks. We compared the performance of different genome partitions and found that the partitions generated from snp ldsplit led to smaller sum of squared correlations between SNPs in different blocks (Supplementary Material, Appendix A).

We used the 1KG Phase 3[26] reference panels for our analysis. We selected the European samples by the SuperPopulation information provided by 1KG and then excluded all duplicated and ambiguous SNPs. We applied quality control to the 1KG data for EUR ancestry using PLINK[28] (–geno 0.05 –hwe 1e-10 –mind 0.05 –maf 0.05) and generated a genetic map using the https://plink.readthedocs.io/en/latest/plink_mani/ website. In addition, we conducted quality control on the UKB data, creating two UKB reference panels with 503 randomly selected non-overlapping samples from the unrelated White British individuals (the same sample size as the EUR 1KG Phase 3 reference panel).

To partition the genome, we excluded the MHC block on chromosome 6 (30-31Mb) and applied snp ldsplit to each chromosome in parallel. To avoid LD leakage and biased estimates, we set the minimum size of each block to be at least 0.5 cM. We adaptively searched for the optimal values of max_r2 (the maximum squared correlation allowed for one pair of variants in two different blocks) and max_size (the maximum number of variants in each block) for each chromosome to make the LD blocks as independent as possible. It is important to note that a larger block may result in increased computational and memory requirements, and obscure local signals. To find the minimal combination of max_r2 and max size that can generate partitions with a mean block size smaller than 1.6cM, we searched for values of max_r2 from 0.3 to 0.72 and max size from 5cM to 13cM. From the partitions found, we selected the partitions that resulted in the minimal cost (the sum of squared correlations between SNPs at different blocks). The final max_r2 and max size values for each chromosome are shown in Supplementary Table 1 and the partitions used in this analysis are in Supplementary Table 2. (Note that genomic coordinates for this paper are in reference to the human genome build 37.)

### 2.3 Methods for genetic correlation estimation

We compared the performance of three local genetic correlation estimation methods: *ρ*-hess, SUPERGNOVA, and LAVA. These three approaches are based on the analysis of summary statistics, with *ρ*-hess and LAVA using fixed effects models and SUPERGNOVA adopting a random effects model. In the following, we first briefly introduce the concept of local genetic covariance and then describe the underlying statistical framework for these methods.

Let *X*_*i*_ denote the standardized genotype vector of size *m*_*i*_ in block *i*, where *m*_*i*_ is the number of markers in this block, i = 1, …, I, and *β*_*i*_ and *γ*_*i*_ are the effect size vectors of the *m*_*i*_ markers within block *i* for two traits, then the local genetic contributions for the two traits in block *i* are 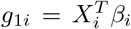 and 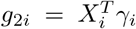, respectively. Local genetic covariance is defined as *ρ*_*i*_ =*Cov*(*g*_1*i*_, *g*_2*i*_). The local genetic correlation *r*_*i*_ can be estimated as 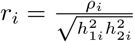 where 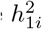 and 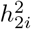 are the local heritability in block *i*. Furthermore, the global genetic covariance matches the sum of local genetic covariance when the genetic components in different partitions are independent:

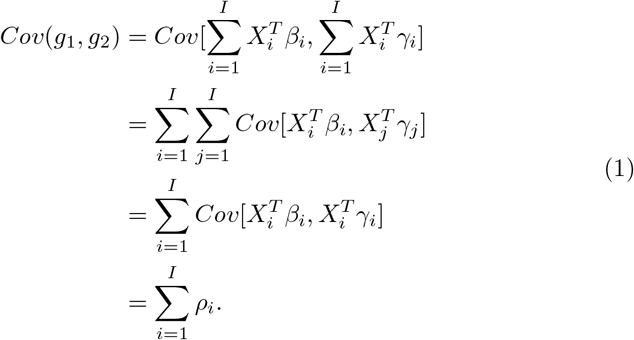

where *g*_1_ and *g*_2_ are the genetic contributions for traits 1 and 2, respectively. In the outputs of these three methods, *ρ*-hess and SUPERGNOVA give estimates of local genetic covariance, whereas LAVA provides estimates of local genetic correlation. All three methods yield estimates of local heritability with p-values provided by *ρ*-hess and LAVA, so we can obtain both local genetic correlation and covariance for all three methods.

#### 2.3.1 *ρ*-hess

Based on the definition of local genetic covariance introduced above, *ρ*- hess defines local genetic covariance in block *i* as 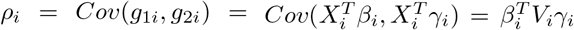 where *V*_*i*_ is the local LD matrix in this block, and *β*_*i*_ and *γ*_*i*_ are the fixed effect size vectors for two traits in block *i*.

When there are two GWASs, with *n*_1_ samples for trait 1 (*ϕ*_1_) and *n*_2_ samples for trait 2 (*ϕ*_2_), *ρ*-hess assumes that 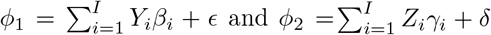, where *ϕ*_1_ is the vector of trait 1 for *n*_1_ samples and *ϕ*_2_ is the vector of trait 2 for *n*_2_ samples, *Y*_*i*_ and *Z*_*i*_ are the standardized genotypes in block *i* for *n*_1_ and *n*_2_ individuals, respectively, and *ϵ* and *δ* are the vectors of noises with 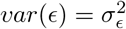 and 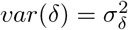. Assume the first *n*_*s*_ samples overlap, then

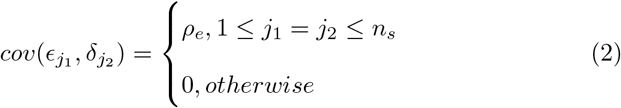

and 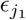 and 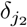 are the noises for individual *j*_1_ in trait 1 and for individual *j*_2_ in trait 2, respectively.

The marginal effect size estimates of SNPs in block *i* from GWAS, 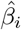 and 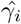, follow the normal distribution 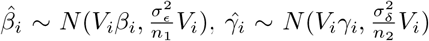, so in the absence of sample overlap, *ρ*-hess estimates the local genetic covariance in block *i* by

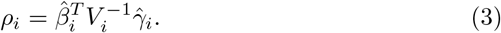

However, due to sample overlap, the estimation based on (3) using 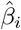 and 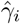 will have a bias term. Besides, in practice *ρ*-hess uses truncated SVD to address rank-deficiency of LD matrix *V*_*i*_ to improve stability, especially when only the external reference panel is available. By defining 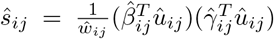, where 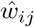 and 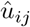 are the *j*th top eigenvalue and its corresponding eigenvector of local LD matrix in block *i* from the external reference panel, and 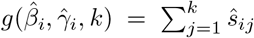 *ρ*-hess estimates local genetic covariance sfter correcting for the bias by

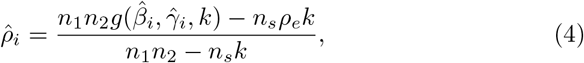

where *k* is the number of top eigenvalues and their corresponding eigenvectors used which can be input by the user and is the same for different blocks.

For testing significance, *ρ*-hess assumes that the sampling distributions of the local genetic correlation and covariance are normal, and uses a parametric bootstrap approach to estimating the standard errors.

In our real data analysis, we followed *ρ*-hess’s suggestion [24] to estimate *λ*_*GC*_ from GWAS data. We then used the estimated 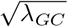 to re-inflate the effect sizes before estimating the local SNP heritability and genetic correlation. To improve the accuracy of *ρ*-hess, we used the global heritability and its standard error from LDSC as extra inputs. The number of shared samples used in our analysis was based on the consortium from which each GWAS was generated. As indicated in Supplementary Table 4, when two traits have samples from the same consortium, we fixed the shared sample size to the minimum sample size of the common consortium. We set the shared sample size to zero when two traits came from completely different consortia. All other parameters were kept at their default values.

#### 2.3.2 SUPERGNOVA

SUPERGNOVA also assumes traits follow the same linear models shown in *ρ*-hess. However, SUPERGNOVA models genetic effects *β*_*i*_ and *γ*_*i*_ as random rather than fixed. More specifically, *β*_*i*_ and *γ*_*i*_ in block *i* follow a multivariate normal distribution:

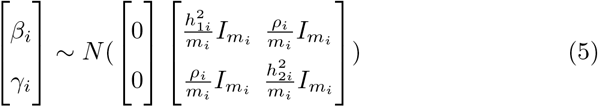

where 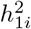 and 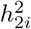 are the local heritability of traits 1 and 2 in block *i*; *ρ*_*i*_ is the local genetic covariance between traits 1 and 2; *I*_*m*_*i* is the identity matrix of size *m*_*i*_; and *m*_*i*_ is the number of SNPs in block *i* as defined before.

The estimator used in SUPERGNOVA is defined in terms of the marginal z-statistics of a single SNP *j* in block *i*, which is given by 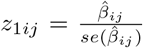 and 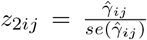, where 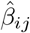 and 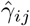 are the marginal effect sizes from GWAS. SUPERGNOVA performs eigen decomposition of the local LD matrix 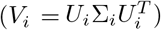 and chooses the first *K*_*i*_ eigenvectors to transform and decorrelate association statistics in a given block *i*, where *K*_*i*_ is determined adaptively. After decorrelation, local genetic covariance *ρ*_*i*_ is estimated by modeling the expected value of the products of the projected z-statistics,

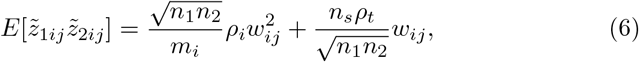

where *w*_*ij*_ is the *j*th eigenvalue, where 1 ≤ *j* ≤ *m*_*i*_, *ρ*_*t*_ is the sum of genetic covariances and non-genetic covariance, i.e., 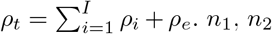, and *n*_*s*_ are the sample sizes for each trait and the sample size shared by two GWASs, respectively, as defined above. Besides, 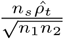 is the estimation of 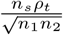 using the intercept of cross-trait LDSC[32]. Then a weighted least squares regression is used to regress 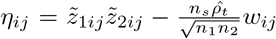 on predictor 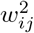 with the weight as the reciprocal of 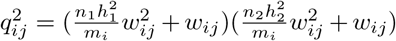, where 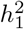 and 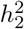 are the global heritability for traits 1 and 2, respectively.

SUPERGNOVA adopts an adaptive procedure to determine the number of eigenvalues/eigenvectors for each block. This is accomplished by choosing *K*_*i*_ which minimizes the maximum between the theoretical variance and the empirical variance of local genetic covariance:

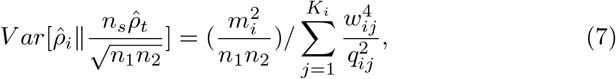

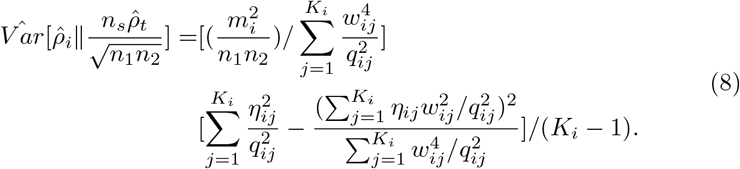

Finally, the variance of local genetic covariance has the following form:

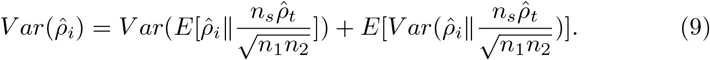

#### 2.3.3 LAVA

Same as *ρ*-hess, LAVA also assumes that the genetic effect sizes are fixed and denotes local genetic covariance in block *i* as 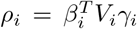. LAVA first applies singular value decomposition to the local LD matrix in block *i* which is 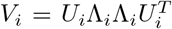, and then defines *U*_*i*\*_ as the *m*_*i*_ by *k*_*i*_ pruned eigenvector matrix and Λ_*i*\*_ as the corresponding *k*_*i*_ by *k*_*i*_ diagonal singular value matrix, where *m*_*i*_ is the number of SNPs in block *i* and *k*_*i*_ is the number of top eigenvalues that could explain 99% variance of the local LD matrix in block *i*. Thus, the inverse of *V*_*i*_ can be approximated as 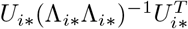.

Furthermore, LAVA defines the scaled principal component (PC) matrix 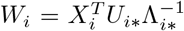 and the corresponding PC effects 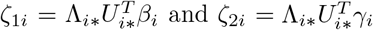, such that 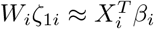 and 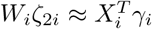. Thus, the local genetic covariance can be represented by the covariance of the PC effects:

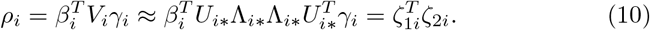

Assume that 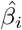 and 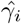 are the vectors of the marginal effects of two traits in block *i*, then based on the distribution of marginal effect sizes, PC effects can be estimated as 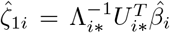 and 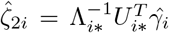, and 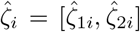 follows the distribution 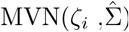, where *ζ*_*i*_ = [*ζ*_1*i*_, *ζ*_2*i*_] and 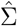 represents the sampling covariance matrix.

The method of moments can be used to estimate:

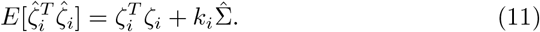

In the absence of sample overlap, 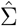 is defined as the diagonal matrix with diagonal elements as the sampling variances of trait 1 and trait 2. When accounting for sample overlap, LAVA first applies LDSC to create a covariance matrix with the intercepts for the global genetic covariance for the off-diagonal elements and each trait’s univariate LDSC intercept as the diagonal elements. Then LAVA converts this covariance matrix to a correlation matrix, C, and computes the sampling correlation matrix as 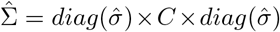, where 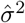 is a vector with the sampling variances of the traits.

Once estimated, the significance of *ρ*_*i*_ is evaluated using simulation-based P values. Based on the definition of non-central Wishart distribution and 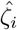 follows 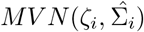, the statistic 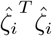 has a non-central Wishart distribution with *k*_*i*_ degrees of freedom, scale matrix 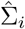 and non-centrality matrix 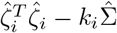.

### 2.4 Simulation settings

The genotype data in all the simulation settings are generated by white British individuals from UKB[25] while the reference panel is either the in-sample reference panel from UKB or the external reference panel from EUR 1KG Phase 3 data across the settings. We first conducted extensive simulations on blocks with different sizes to evaluate the performance of *ρ*-Hess, SUPERGNOVA, and LAVA under 1) varying sample overlaps between two GWASs, 2) both continuous and binary traits, 3) both infinitesimal and non-infinitesimal models, 4) the presence or absence of correlations between effect sizes and LD, and 5) different reference panels. Then based on the above simulation results, we further investigated the impact of the number of eigenvalues and eigenvectors on the stability and inference of *ρ*-hess and LAVA. We repeated each simulation setting 100 times and summarize the simulation settings in Table 1 and describe the details below.

**Table 1.**
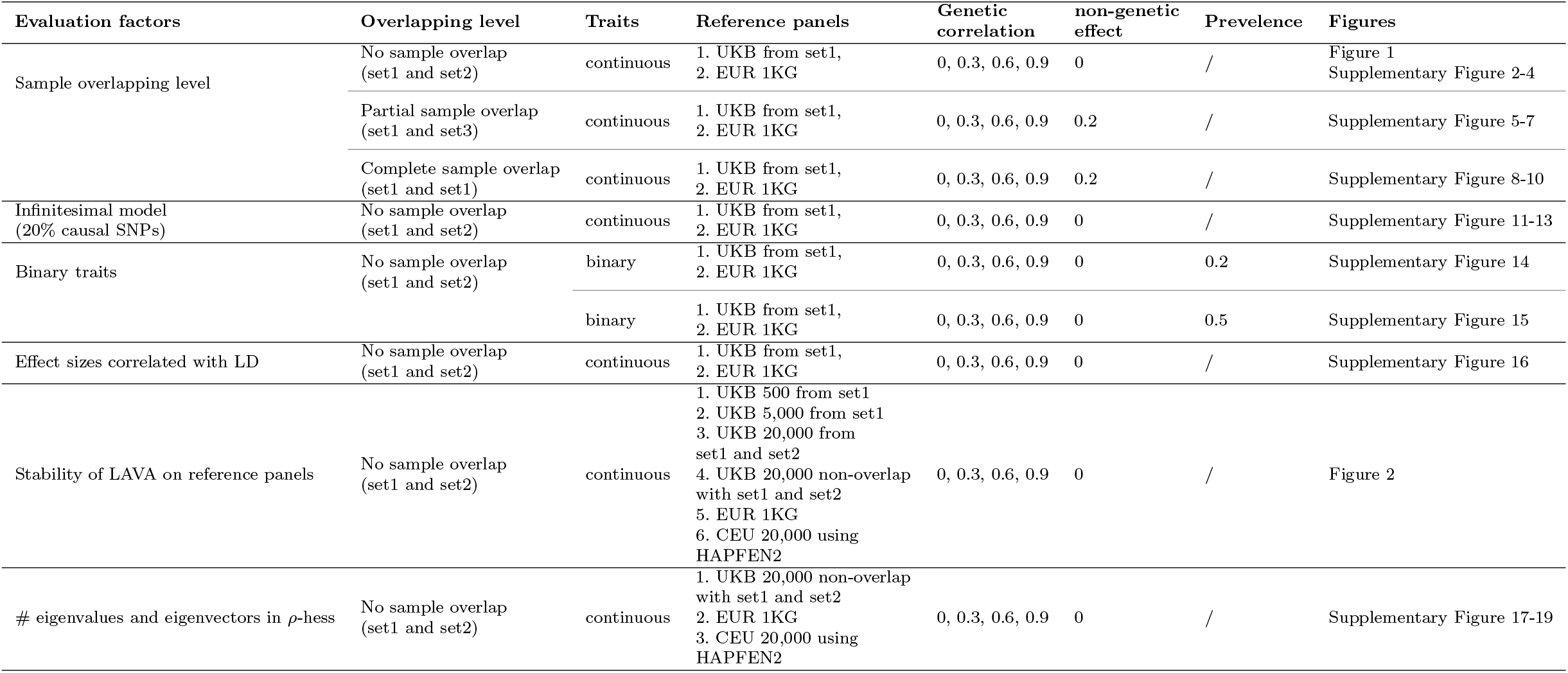
Details of each simulation setting

We selected overlapping SNPs from chromosome 1 in UKB, EUR 1KG Phase3, and HapMap3[33] datasets for efficient simulations and to ensure sufficient SNP coverage. We then selected SNPs with MAF > 5%, genotype missing rate < 5%, and Hardy-Weinberg equilibrium P-value > 1e-10. After removing SNPs with ambiguous alleles, 71,609 SNPs remained for our simulation. We randomly selected 20,000 unrelated white British individuals from UKB and divided them into two subgroups of 10,000 individuals each, labeled as set1 and set2, respectively. We formed another set3 with 5,000 individuals from set1 and 5,000 individuals from set2. We randomly selected four blocks on chromosome 1 having 525 SNPs (POS: 60197393-61754126), 743 SNPs (POS: 3264297-5311384), 1033 SNPs (POS: 245966297-249239303), and 2315 SNPs (POS: 113753415-146215362), respectively. We treated one block as the local region of interest in each simulation and the other SNPs as the background SNPs. We simulated two traits whose SNP effects followed the multivariate normal distribution, with correlation only for SNPs within the chosen region of interest. The correlation of the local genetic effects was set to be 0, 0.3, 0.6, and 0.9, respectively. The remaining SNPs on chromosome 1 were considered background SNPs without genetic correlation. We set the heritability of two traits to be 0.5 which was evenly distributed to all SNPs (71,609 SNPs), so the local heritability of the above four blocks was 0.0037, 0.0052, 0.0072, and 0.0162, respectively. Genome-wide Complex Trait Analysis (GCTA)[34] was applied to simulate continuous traits *ϕ*_1_ and *ϕ*_2_. We used PLINK[28] to analyze the simulated traits and generate GWAS summary statistics.

We considered no sample overlap, partial sample overlap, and complete sample overlap. When there was no sample overlap, two continuous traits, *ϕ*_1_ and *ϕ*_2_, were simulated on set1 and set2, respectively. For *ϕ*_1_ and *ϕ*_2_ with partial sample overlap, set1 and set3 were used and the covariance of non-genetic effects was set to 0.2. As for the case where the samples were completely overlapping, we used set1 to simulate both *ϕ*_1_ and *ϕ*_2_ and the covariance of non-genetic effects was still set to 0.2. Additionally, we considered the situation that 20% of the SNPs were causal SNPs, where we randomly chose 20% of the SNPs in the regions of interest and 20% of SNPs in the background to be causal. For this case, we considered the no-sample overlap case where the two traits were continuous. We also conducted a simulation where the two traits were binary with no sample overlap. We considered the same local regions of interest, heritability, and genetic correlation for continuous traits. We used a liability model to simulate the binary traits. We first simulated continuous traits *ϕ*_1_ and *ϕ*_2_ and then the binary traits were set to be *I*[*ϕ*_1_ > *γ*] and *I*[*ϕ*_2_ > *γ*], where *γ* was the quantile of standard normal distribution. Since we considered two simulation settings with *γ* to be 80% or 50%, the prevalence of the binary traits in the two simulations was 0.2 or 0.5. Besides, we also considered the situation where the effect sizes were correlated with local LD, which was the baseline in the LAVA simulation which was also mentioned in SUPERGNOVA. In the simulation setting, LAVA first decomposed the local LD matrix of the reference panel as 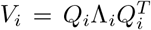 for block *i* and obtained the subset of eigenvalues 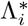 and eigenvectors 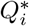 that explained 99% of the variance. We denote the number of eigenvalues thus selected as *q*. LAVA defined the projected genotype matrix in its simulation setting for block *i* as 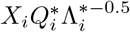 where *X*_*i*_ is the standardized genotype of the reference panel in block *i*. It then generated *δ*_*_, a *q*×2 matrix with 0 means and identity variance, and decomposed the variance-covariance matrix of the genetic components as Ω = *Q′* Λ*′ Q′*^*T*^ and set *δ* = *Q′* Λ*′*^0.5^ *δ*_*_. It simulated the genotype component for two traits as *G*_*i*_ = *W*_*i*_*δ*. Thus, the effects in the simulation settings of LAVA were 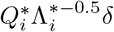 where the effect sizes were correlated with local LD. We note that SUPERGNOVA also conducted simulations when the effect sizes were associated with ldscore. Thus, we also considered the simulation setting where the effect sizes were generated similarly from the simulation setting in LAVA[21] so that the effect sizes were related to local LD. For each simulation setting described above, we used both the in-sample reference panel from the UKB set1 samples and one external reference panel from the 1KG Phase 3 data.

Across the above simulation settings, the performance of LAVA was sensitive to the choice of reference panels. To further investigate the stability of LAVA, we applied LAVA using six different reference panels, 1) EUR 1KG Phase3 reference panel, 2) UKB reference panel with 500 randomly selected individuals from set1, 3) UKB reference panel with 5,000 randomly selected individuals from set1, 4) UKB reference panel with all 20,000 individuals from set1 and set2, 5) UKB reference panel with 20,000 individuals randomly selected from unrelated white British populations in UKB which do not overlap with set1 and set2, and 6) 20,000 CEU individuals simulated using HAPGEN2[35](CEU refers to Northern Europeans from Utah).

Even though the *ρ*-hess-based estimates were more stable when using different reference panels, there was a substantial difference in statistical inference. As *ρ*-hess allows users to change the number of eigenvalues, we considered different reference panels and varied the number of eigenvalues to further investigate the performances of *ρ*-hess (the EUR 1KG Phase 3 reference panel; the UKB reference panel with samples from set1 and set2; and the CEU reference panel using HAPGEN2 with 20,000 individuals). For each reference panel, we varied the number of eigenvalues to explain 99%, 95%, 90%, 85%, 80%, and 70% variance in the above-selected blocks.

### 2.5 GWAS summary statistics

We analyzed 31 complex traits whose GWAS summary statistics are publicly available. These GWASs were primarily generated using individuals of European ancestry. The sources, sample sizes, and global heritability for these traits are listed in Table 2. To prepare data for analysis, we employed the munge sumstats.py script from LDSC[32] to reformat and conducted quality control on the datasets, including the elimination of strand-ambiguous SNPs and the intersection of the remaining SNPs with those from the 1KG Project. In our analysis, we considered only autosomal SNPs with MAF > 5% and excluded the MHC block on chromosome 6 (30-31Mb).

**Table 2.**
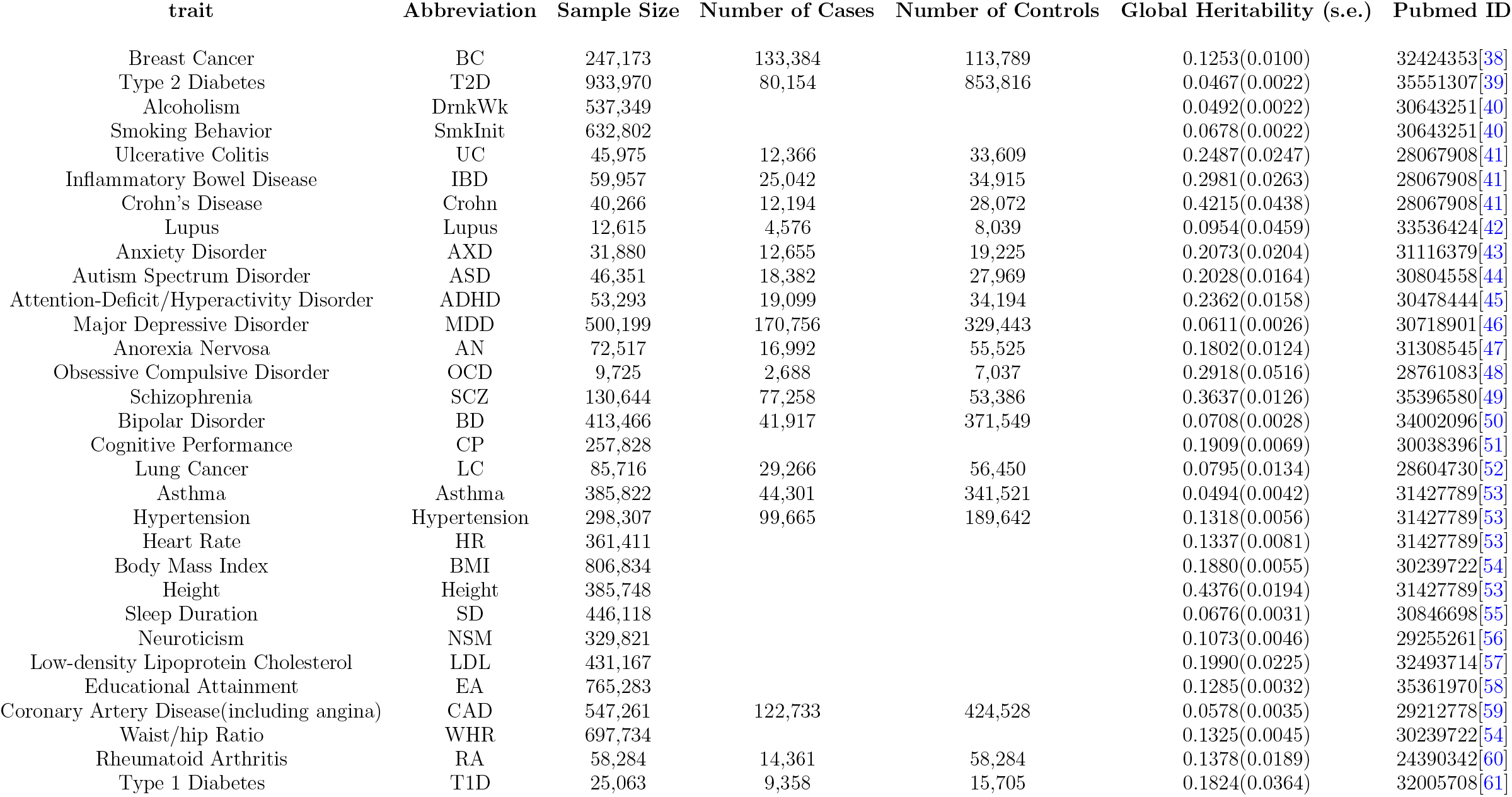
Overview of the traits included in this study

### 2.6 Polygenic risk score (PRS) analysis

We used the positively correlated and negatively correlated blocks from *ρ*- hess, SUPERGNOVA, and LAVA (with FDR < 0.1) between ASD and CP, CAD and LDL, and T2D and BMI to construct PRS+ and PRS- for ASD, CAD, and T2D, respectively. These SNPs were clumped using PLINK, with a significance threshold of 1 for index SNPs, an LD threshold of 0.1 for clumping, and a physical distance threshold of 250kb. PRSs were generated for ASD probands in the SPARK cohort and CAD and BMI cases in the UKB dataset. In addition, we compared CP (measured by IQ), LDL, and BMI between patients with high PRS+ and those with high PRS- for relevant disorders.

## 3 Results

### 3.1 Simulation Results

#### 3.1.1 Basic Simulation analysis

We compared the performance of *ρ*-hess, SUPERGNOVA, and LAVA by point estimation of local genetic correlation, local genetic covariance, local heritability, type I error, and statistical power. Since all three methods can use customized reference panels, we performed simulations on both the in-sample reference panel and the external reference panel with matched ancestry to investigate the robustness of these methods to the choice of LD reference panels.

We considered the simulation settings described in the Methods section to simulate traits based on the genotype data in the UKB and used the EUR 1KG Phase3 data and sample set1 from UKB as reference panels. For the continuous traits generated from non-overlapping samples (set1 and set2), SUPERGNOVA provided unbiased estimates for local genetic correlation, local genetic covariance, and local heritability (Figure 1A and Supplementary Figure 2 and 3) in all settings, and the results were robust to the choice of reference panel. However, SUPERGNOVA sometimes had inflated type-I error (Figure 1B and Supplementary Figure 4). When the EUR 1KG Phase 3 reference panel was used, which did not match the GWAS samples, LAVA overestimated local genetic covariance and local heritability and underestimated local genetic correlation (Figure 1A and Supplementary Figure 2 and 3), and the higher the local genetic covariance, the less accurate the point estimates obtained from LAVA. LAVA had a higher inflated type-I error (about 20%) (Figure 1B and Supplementary Figure 4) than SUPERGNOVA and *ρ*-hess. On the other hand, if the in-sample UKB reference panel was used, LAVA yielded unbiased estimates for local genetic covariance and local heritability, and more accurate local genetic correlation estimates with well-controlled type-I error (Figure 1 and Supplementary Figure 2-4). In contrast, regardless of the reference panels used, *ρ*-hess always underestimated local genetic covariance and local heritability, particularly when local genetic covariance and local heritability were high (Supplementary Figure 2 and 3), but it provided unbiased local genetic correlation estimation, which may be due to compensation for both underestimated local genetic covariance and local heritability. The statistical test based on *ρ*-hess was overly conservative, leading to reduced statistical power (Figure 1B, Supplementary Figure 4).

**Fig. 1.**
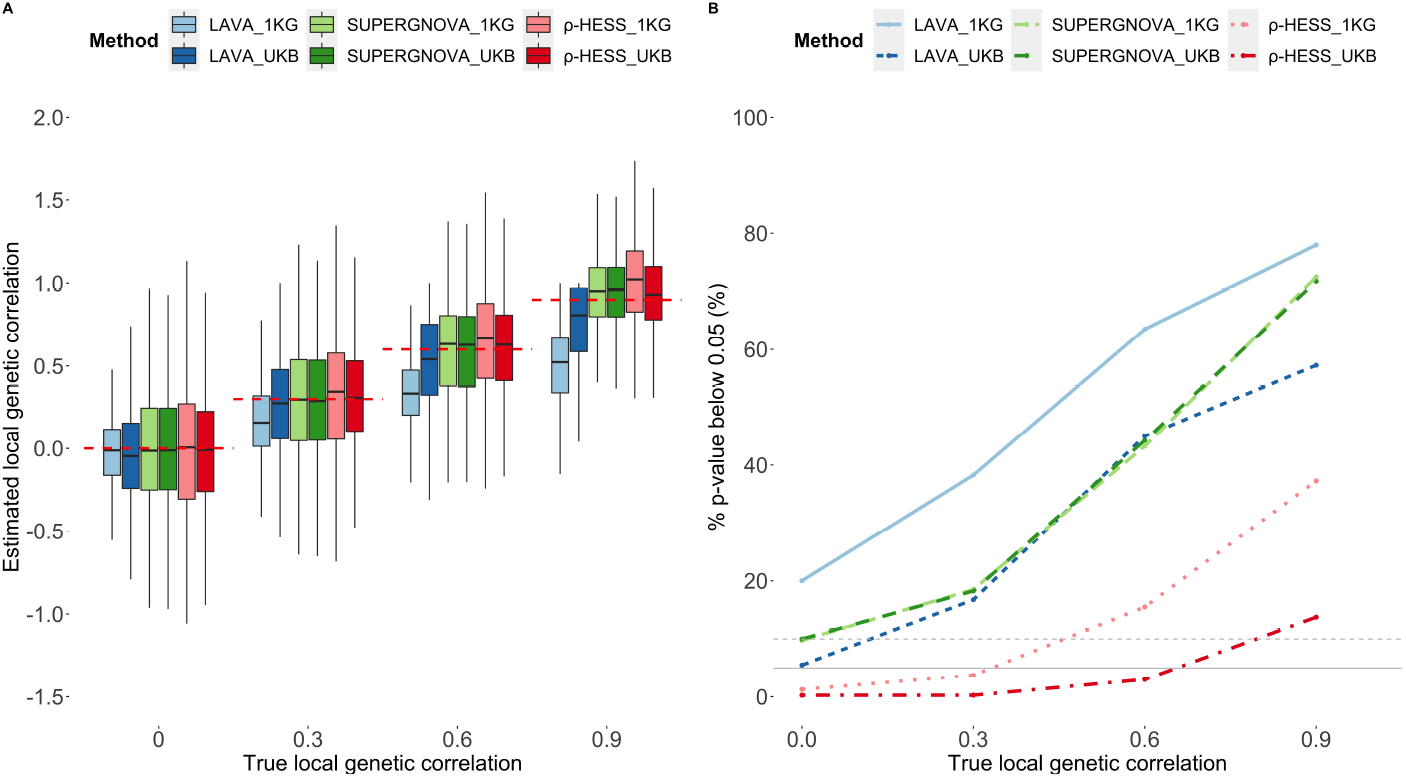
Evaluation of local genetic correlation/covariance methods on continuous traits from non-overlapping datasets (set1 and set2) using EUR 1KG Phase3 and UKB reference panel. (A) local genetic correlation estimates. The red dashed lines represent the true value of local genetic correlation. (B) type-I error and statistical power. The solid grey line represents 5% p-values below 0.05 in 100 repeats, and the grey dashed line represents 10% p-values below 0.05 in 100 repeats.

Since the shared sample size between two traits needs to be provided to *ρ*-hess, we used both the correct shared sample size and incorrect overlapping sample size (1,000) to investigate the impact of this parameter on *ρ*-hess for the partial and complete overlapping scenarios. In this case, the performance for point estimate and inference by SUPERGNOVA was the same (Supplementary Figures 5-10). However, LAVA did not have well-controlled type-I error with overlapping samples, even when the in-sample reference panel was used (Supplementary Figures 5 and 8). With an incorrect overlapping sample size, *ρ*-hess had much reduced statistical power (Supplementary Figures 5 and 8).

When only some SNPs were causal, the performance of different methods was similar when all the SNPs were set to be causal, except for LAVA having some inflated type-I error even using the in-sample reference panel. This suggests that the sparsity of causal SNPs does not have much impact on the performances of local genetic correlation/covariance estimation (Supplementary Figure 11-13).

Since all three methods can be applied to binary traits, we also considered binary traits in our simulation. Even though the genetic covariance is estimated on the observed scale, there is no distinction between observed- and liability-scale genetic correlation[14]. The estimates for the genetic correlation of binary traits had similar performances to that for continuous traits except with larger variations across the 100 repeats (Supplementary Figures 14A and 15A). However, the statistical power for binary traits was lower for all methods compared to continuous traits, especially *ρ*-hess which barely detected any significant blocks in our simulations (Supplementary Figures 14B and 15B).

#### 3.1.2 LD-related effect sizes

For simulations where the effect sizes were associated with the local LD structure, similar to the simulation setting in LAVA, there was a substantial underestimation of local genetic covariance (Supplementary Figure 16) with nearly no significant block detected by SUPERGNOVA and *ρ*-hess with the default settings. When we gave *ρ*-hess the number of eigenvalues that could explain 99% variance and used the UKB reference panel, the performance of *ρ*-hess improved but still had lower power than LAVA. In this setting, LAVA had the best performance in this simulation setting except for the inflated type-I error when using EUR 1KG Phase 3 data as the reference panel.

#### 3.1.3 Robustness to reference panels

Our simulation results suggest that with different reference panels, the results from LAVA can be unstable, thus we further investigated the choice of the reference panels on LAVA. We considered the EUR 1KG Phase 3 reference panel, four different UKB reference panels, and one CEU reference panel (Method). As shown in Figure 2, when we used two UKB reference panels with 20,000 participants, LAVA had unbiased estimations and well-controlled type-I errors. With smaller UKB reference samples, LAVA could not provide reliable local genetic correlation estimates and well-controlled type-I errors. By comparing the performance between using the CEU reference panel and the UKB reference panel with the same sample size (20,000), the sample size of the reference panel was not a key factor for the performance of LAVA. Our results suggest that LAVA only performs well with enough individuals from the genotype data cohorts used as the reference panel.

**Fig. 2.**
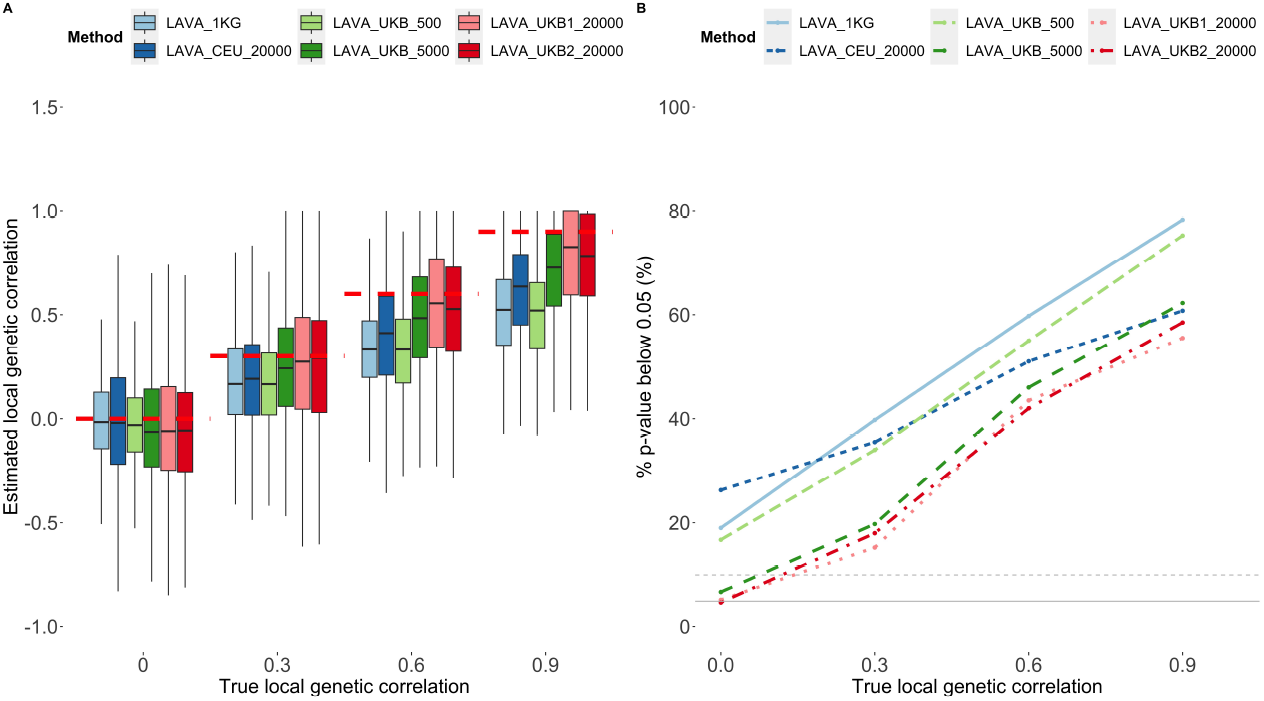
Evaluation of local genetic correlation estimated by LAVA on continuous traits from non-overlapping datasets (set1 and set2) using different reference panels. (A) local genetic correlation estimates. The red dashed lines represent the true value of local genetic correlation. (B) type-I error and statistical power. The solid grey line represents 5%, and the grey dashed line represents 10%. LAVA_1KGrepresents the EUR 1KG Phase3 reference panel; LAVA_CEU_20000 represents the CEU reference panel using HAPGEN2; LAVA_UKB_500 represents the UKB reference panel using 500 individuals randomly selected from set1; LAVA_UKB_5000 represents the UKB reference panel using 5,000 individuals randomly selected from set1; LAVA_UKB1_20000 represents the UKB reference panel using 20,000 individuals from set1 and set2; LAVA_UKB2_20000 represents the UKB reference panel using 20,000 individuals that do not overlap with set1 and set2.

#### 3.1.4 Number of eigenvalue input

By using the EUR 1KG Phase3, UKB samples from set1 and set2 and 20,000 CEU individuals(Method) as reference panels, we investigated whether the optimal number of eigenvalues used in *ρ*-hess stays more or less the same for different blocks and different reference panels. The optimal number of eigenvalues here is the one that could result in well-controlled type-I errors and higher powers, although the point estimates derived using these numbers could still be biased. When the reference panel was the same as that from the GWAS samples, the more eigenvalue used, the better the performance for *ρ*-hess, although it still had limited statistical power (Supplementary Figure 18 and Supplementary Figure 4). With an external reference panel that is different from the GWAS samples, there was no consistent pattern observed, and the optimal number of eigenvalues varied for blocks and reference panels (Supplementary Figures 17, 19 and Supplementary Table 3). When using the EUR 1KG Phase 3 reference panel, the optimal number of eigenvalues was 93 (95%), 153 (90%), 146 (95%), and 85 (70%), respectively. For the CEU reference panel, the optimal number was 93 (95%), 310 (99%), 42 (70%), and 572 (99%), respectively. These observations could explain the poorer performance of *ρ*-hess in Figure 1 when using the in-sample UKB reference panel than the external 1KG Phase3 reference panel. This is because when the in-sample reference panels were used, the larger the number of eigenvalues used, the better the performance, while in Figure 1, the default number was 50.

### 3.2 Local genetic correlation/covariance of 31 complex traits

We considered 31 complex disorders or traits to compare the performance of the three methods. Table 2 summarizes these traits, abbreviations, sample sizes (the number of cases and the number of controls for binary traits), global heritability and its standard error derived from LDSC[14], and the original papers.

#### 3.2.1 Stability based on different reference panels

When using only EUR 1KG Phase 3 genotype data as our reference panel to estimate the local genetic correlation for the above 31 complex traits, there were substantial differences in point estimates and inferences by *ρ*- hess, SUPERGNOVA, and LAVA (Supplementary Material Appendix B). we focused on comparing the point estimates and detecting significant blocks using different reference panels for the same method in this section because the simulation results showed the importance of the reference panels for both *ρ*-hess and LAVA. Since the heritability of Height is the largest among all the traits, for clearer and more efficient comparison we decreased the number of trait pairs estimated and compared the results of the genetic correlation between Height and other traits using two other reference panels which were generated using different randomly selected white British UKB samples (Methods) and EUR 1KG Phase 3 reference panel. We compared the local heritabilities and local genetic correlations in the same block for the same trait or trait pairs using the same methods but different reference panels. As seen in Supplementary Figures 26-31, SUPERGNOVA displayed the most stable point estimates for local genetic correlation and local heritability using different reference panels and the estimates from *ρ*-hess were more stable than LAVA. Even with two different references from the same cohort, i.e. the two UKB reference panels, LAVA resulted in different estimates for the same block for the same pair of traits (Supplementary Figures 30 and 31).

Since the sum of local heritability should equal global heritability and the local genetic covariance should equal global genetic covariance (Methods), we further compared the sum of local heritability and local genetic covariance with global heritability and global genetic covariance with different reference panels. As shown in Supplementary Figure 32, LAVA tended to overestimate local heritability which is consistent with simulation results when the samples of the reference panel and the GWAS did not match. The sum of local heritability was highly concordant with the estimated global heritability for *ρ*-hess. For SUPERGNOVA, except for three traits, Lupus, OCD, and T1D, the sum of local heritability was also highly concordant with the global heritability. The sum of local genetic covariance had a high correlation with the global genetic covariance for SUPERGNOVA and LAVA but was lower for *ρ*-hess (Figure 3). As shown in Figure 4 and Supplementary Figure 34, the significant blocks found by SUPERGNOVA and *ρ*-hess were consistently detected using different reference panels, while the results differed substantially for LAVA with different reference panels.

**Fig. 3.**
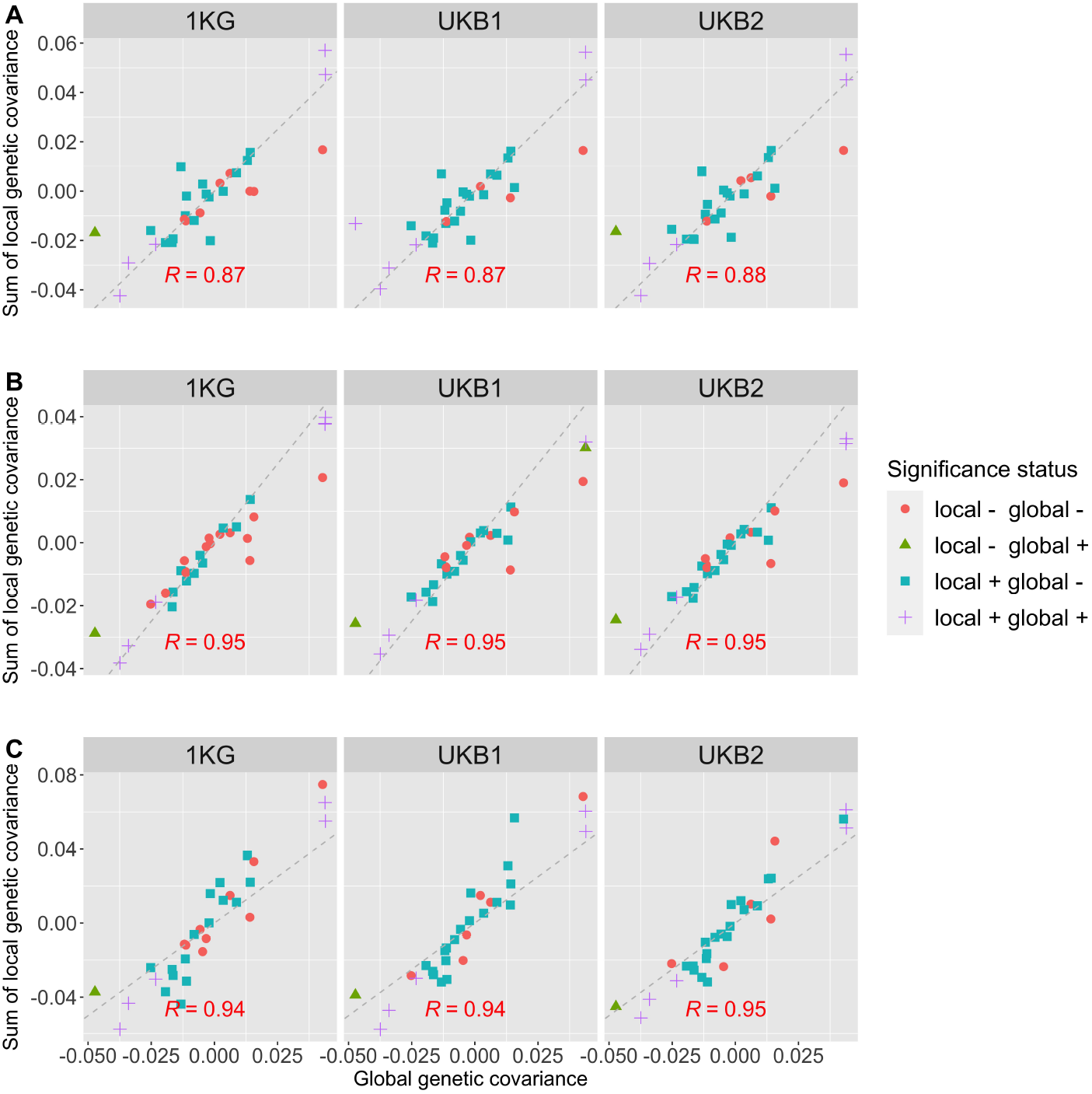
Comparisons of the sum of local genetic covariance for 465 trait pairs with the global genetic covariance derived from LDSC. A. Comparisons of the sum of local genetic covariances estimated from A *ρ*-hess, B SUPERGNOVA or C LAVA with the global genetic covariances using three different reference panels. Each point represents a trait pair. The color and shape of each data point denote the significance status in global and local correlation analyses. ‘local +’ denotes that there are significant blocks detected between that trait pair and ‘local -’ denotes that there are no corrected blocks detected. ‘global +’ denotes the global genetic correlation is significant, while ‘global -’ denotes that the global genetic correlation is not significant. The figures are divided into multiple panels, with each panel corresponding to different reference panels (EUR 1KG reference panel and two UKB reference panels with different samples). The ashed, grey reference line with a slope of 1 represents the line of perfect correlation in each panel. The strength of the relationship is indicated by Pearson correlation coefficients, which are displayed at the bottom of each panel.

**Fig. 4.**
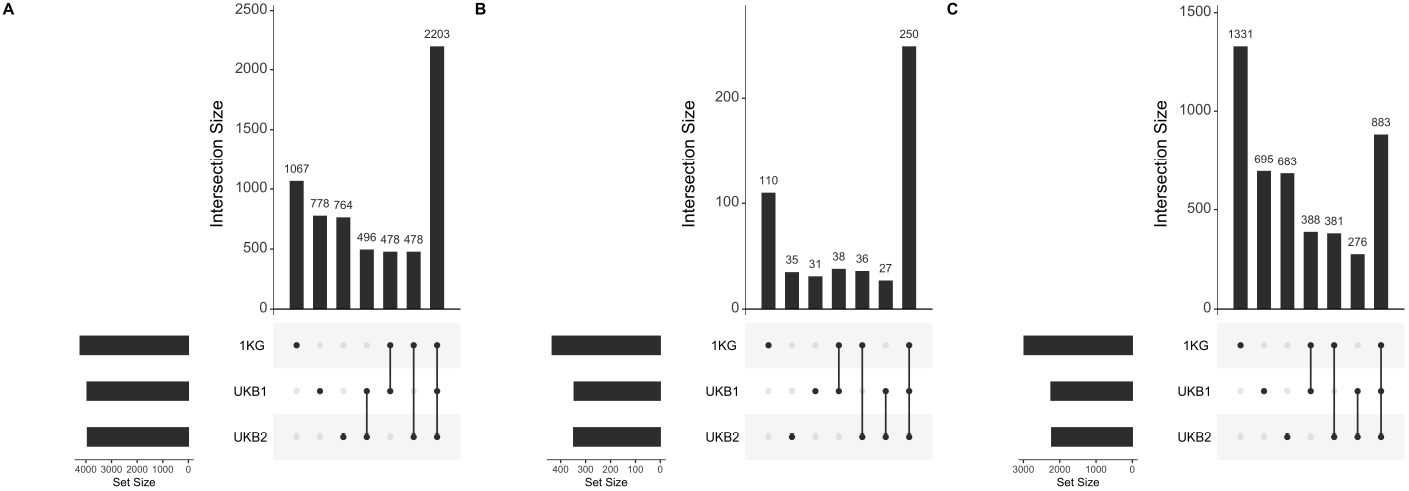
Comparisons of blocks with significant local genetic correlations when using different reference panels. These plots used bars to break down the Venn diagram of overlapped significant blocks using different reference panels using FDR at 0.1 level detected by A. *ρ*-hess, B. SUPERGNOVA, and C. LAVA.

#### 3.2.2 PRS analysis

Several studies[20, 36] including SUPERGNOVA have investigated the shared genetics among autism spectrum disorder (ASD), Attention-deficit/hyperactivity disorder (ADHD), and cognitive ability (CP) by utilizing local genetic information. To further compare the results of *ρ*-hess, SUPERGNOVA, and LAVA, we applied these methods to ASD, ADHD, and CP. By using a false discovery rate (FDR) cutoff of 0.1, we identified one block by *ρ*-hess, 55 blocks by SUPERGNOVA, and 126 blocks by LAVA with significant local genetic covariances between ADHD, ASD, and CP (Supplementary Table 8), respectively. The only block identified by all three methods was on chromosome 6 which was positively correlated between ASD and CP (POS: 97094444-98938023). Additionally, this is the same block that was significantly correlated between ASD and CP by both LAVA and SUPERGNOVA. The global genetic correlation between ASD and CP was 0.2 (p=1.8e-10), between ASD and ADHD was 0.36 (p=1.14e-11), and between ADHD and CP was -0.38 (p<1e-11) revealing that the local correlations of CP with ASD and ADHD were bidirectional. As in Supplementary Figure 33, there was no significant block with a negative correlation between ASD and ADHD identified using LAVA and there were only two such blocks detected by SUPERGNOVA. Besides, there was no block where ASD and ADHD showed opposite correlations with CP. SUPERGNOVA identified 12 blocks with positive correlations and four blocks with negative correlations between ASD and CP. LAVA identified 14 positively correlated blocks and 14 negatively correlated blocks between the two traits. We constructed positive and negative polygenic risk scores (Methods), referred to as PRS+ and PRS-, of ASD based on independent SNPs from blocks with significant positive or negative local correlations between ASD and CP detected by SUPERGNOVA or LAVA, respectively, for 1,026 ASD probands who had both genotypes and IQ scores in SPARK (Methods). We observed probands with high PRS+ had higher IQ than probands with high PRS- only in PRSs generated utilizing SUPERGNOVA (Figure 5A-I). No negative blocks were detected by *ρ*-hess, resulting in only PRS+ constructed based on *ρ*-hess (Figure 5C, F, I). When using PRS+ and PRS-based on SUPERGNOVA, there was a sharp change in the right tails of the PRS distribution analysis of the average full-scale IQ, from 84.7 and 83.1 in the 75th percentile to 89.9 and 75.0 in the 99th percentile for PRS+ and PRS-, respectively (Figure 5A). Similarly, the average non-verbal IQ (Figure 5D) and verbal IQ (Figure 5G) also showed a sharp change in the right tail of the PRS distribution, with respective changes from 93.2 and 92.5 in the 75th percentile to 101.7 and 84.0 in the 99th percentile, and from 94.9 and 91.4 in the 75th percentile to 102.1 and 80.4 in the 99th percentile for PRS+ and PRS-, respectively.

**Fig. 5.**
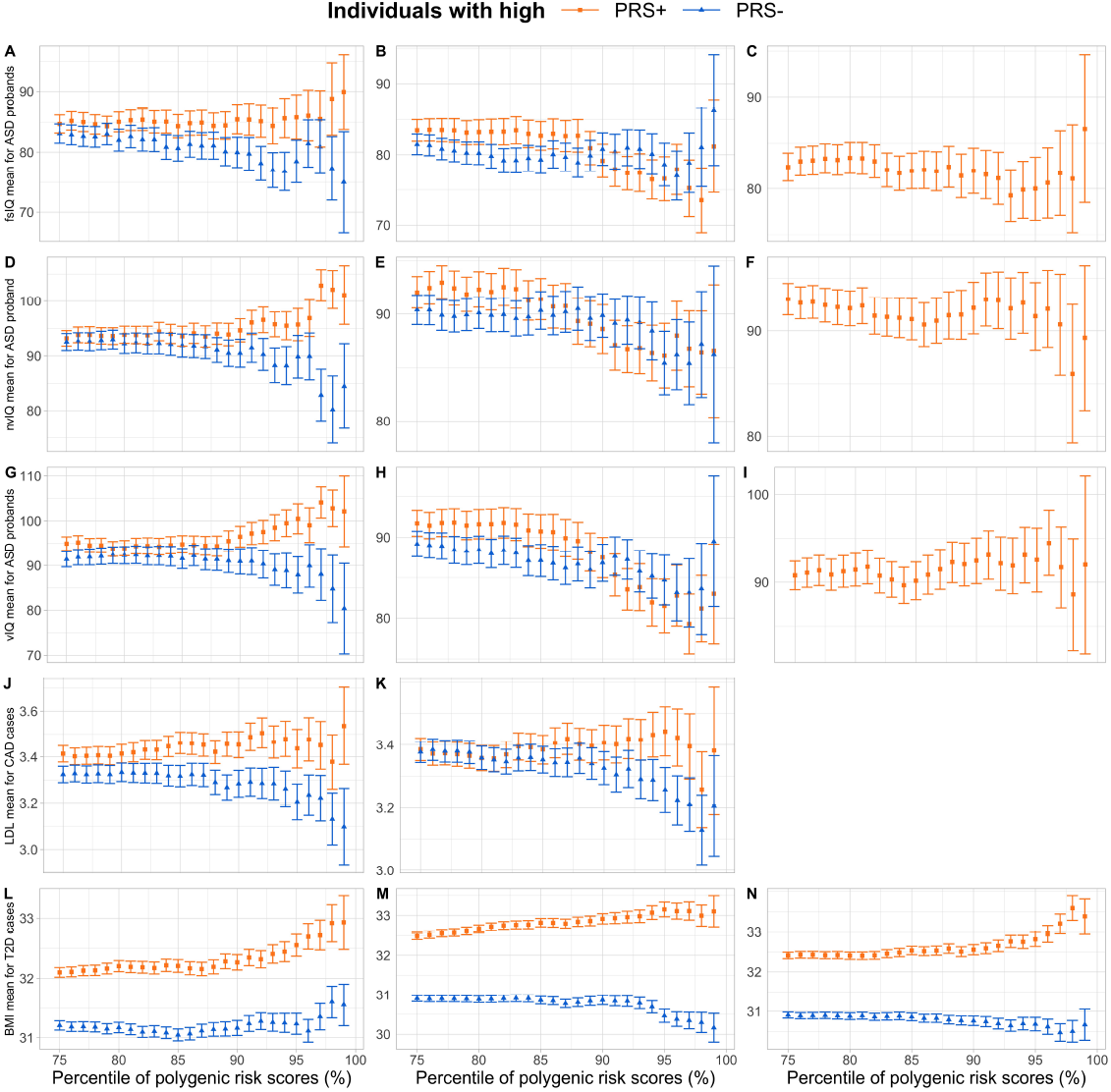
Phenotype heterogeneity of ASD probands, CAD and T2D patients with high PRS+ and PRS. Average full-scale IQ is computed for different groups defined by PRS based on the significant blocks found by A. SUPERGNOVA B. LAVA and C. *ρ*- hess. Average non-verbal IQ is computed for different groups defined by PRS based on the significant blocks found by D. SUPERGNOVA E. LAVA and F. *ρ*-hess. Average verbal IQ is computed for different groups defined by PRS based on the significant blocks found by G. SUPERGNOVA H. LAVA and I. *ρ*-hess. Average LDL is computed for different groups defined by PRS based on the significant blocks found by J. SUPERGNOVA K. LAVA. Average BMI is computed for different groups defined by PRS based on the significant blocks found by L. SUPERGNOVA M. LAVA and N. *ρ*-hess. Each interval indicated the standard error of the average values.

The LAVA study[21] explored the relationship between LDL and CAD, and between BMI and T2D from the angle of multivariate correlation. Here we conducted a PRS analysis using the results of the bivariate local genetic correlation between LDL and CAD, and BMI and T2D. The global genetic correlation between LDL and CAD was 0.3 (p< 1 × 10^−15^). SUPERGNOVA identified 36 positive blocks and 5 negative blocks with significant local genetic correlations between LDL and CAD at an FDR level of 0.1, and LAVA identified 108 positive blocks and 30 negative blocks (Supplementary Table 9). No significant block was identified using *ρ*-hess. SUPERGNOVA and LAVA identified 22 common blocks that had consistent correlation directions, including 21 positive blocks and one negative block on chromosome 5. As displayed in Figure 5J-5K, CAD cases with high PRS+ had higher LDL than cases with high PRS- for both SUPERGNOVA and LAVA, with an average LDL changing from 3.41 and 3.32 for the 75 percentile to 3.54 and 3.10 for the 99 percentile when using SUPERGNOVA. However, the trend in LAVA was less apparent, with the average LDL moving from 3.39 and 3.38 for the 75 percentile to 3.38 and 3.21 for the 99 percentile.

When analyzing the local correlations between T2D and BMI whose global genetic correlation was 0.57 (p< 1 × 10^−15^), *ρ*-hess identified 279 significant blocks, SUPERGNOVA identified 176 ones, and LAVA identified 589 blocks (Supplementary Table 10). A total of 93 blocks were found by all three methods with just one block on chromosome 3 showing a negative correlation between T2D and BMI, and all these 93 blocks had consistent correlation direction. Among the significant blocks, *ρ*-hess found 271 that were positively correlated and eight that were negatively correlated, SUPERGNOVA identified 170 that were positively correlated and 6 that were negatively correlated, and LAVA identified 66 that were positively correlated and 23 that were negatively correlated. As demonstrated in Figure 5L-5N, T2D cases with high PRS+ had a greater BMI than cases with high PRS- for all three methods. For SUPERGNOVA, the average LDL changed from 32.1 to 31.2 for the 75 percentile to 32.9 and 31.6 for the 99 percentile. For LAVA, the average LDL changes from 32.5 to 30.9 for the 75 percentile to 33.1 and 30.2 for the 99 percentile. For *ρ*-hess, the average LDL changes from 32.4 and 30.9 for the 75 percentile to 33.4 and 30.7 for the 99 percentile.

## 4 Discussion

In recent years, there has been an increasing interest in inferring local genetic correlation in post-GWAS analyses in addition to global genetic correlation. This trend can be attributed to advancements in methodologies for estimating local genetic correlation and detecting locally significant blocks, as well as a growing knowledge of the limitations of global genetic correlation for revealing the underlying genetic similarity between complex traits. Local genetic correlation has also been utilized to improve association studies and PRS prediction.

The first step for local genetic correlation is determining how to partition the whole genome into approximately independent blocks. The larger the blocks, the more independent the partitions, but larger blocks may mask local information in the same way that global genetic correlation does. On the other hand, smaller blocks may result in LD leakage and biased estimates. The three methods compared in this paper all provide their own partitions, but also allow users to use their own partitions. Another issue that needs to be addressed for local genetic correlation is also considered for global correlation, i.e. how to deal with pervasive sample overlap across GWASs. The common solution for these three methods is to utilize the cross-trait LDSC intercept to calculate the phenotypic correlation. *ρ*-hess is the only method that requires the shared sample size between two GWASs as input. However, as the number of GWASs generated by meta-analysis grows, the exact number of overlapping sample sizes is difficult to obtain. Our simulation results suggest that the power of *ρ*-hess will decrease if an incorrect number of shared sample size is given. The other two methods have more stable performances in terms of the sample-overlapping level. The third and most crucial challenge with these three methods is estimating the local LD structure using external reference panels. Ideally, the external reference panels applied should have the same LD structure as the genotype data used to calculate summary statistics. In the real world, because access to individual-level data from the GWAS dataset is typically limited due to practical constraints, it is common to choose an external reference panel. Through extensive simulations and real data analysis, we have demonstrated that the choice of the local LD matrix is critical for both estimation and inference. SUPERGNOVA is the most robust method for the choice of reference panels because it has an adaptive procedure to choose the number of eigenvalues and eigenvectors used for different blocks and different reference panels. However, the type-I error of SUPERGNOVA is still inflated in some simulation settings which indicates a better adaptive procedure is still needed. LAVA recommends using the number of eigenvalues and eigenvectors that explain 99% of the variances and performed the best when the genotype data and the reference panel were perfectly matched. However, with different reference panels, LAVA could provide different estimations, and the significant blocks detected were also not consistent. *ρ*-hess needs to be given the number of eigenvalues as input and the default number is set to be 50. However, the optimal number of eigenvalues and eigenvectors depends on both the local LD structure of the reference panels and the LD structure of the blocks in the genotype data. In summary, *ρ*-hess can provide unbiased estimates if the proper number of eigenvalues is selected based on different reference panels, while SUPERGNOVA yields unbiased estimates assuming the underlying assumption holds true. Additionally, LAVA produces unbiased estimates when an in-sample reference panel with sufficient sample sizes is utilized. While *ρ*- hess generally has well-controlled type-I error rates, it may have lower power. SUPERGNOVA is generally more stable across different reference panels, but may have slightly inflated type-I error at times. LAVA only produces well-controlled type-I error rates when an in-sample reference panel with sufficient sample sizes is used.

Despite extensive simulation settings and real data sets considered, there are limitations in our study. First, the methods compared in this study are those that can reveal correlated blocks between two traits within a single population (e.g. European). All these three methods can provide both estimates and references with user-defined partitions. However, there are other methods that could detect corrected blocks between different populations (e.g. European and African) for the same trait[37] or evaluate the concordance of two traits on the method-defined regions[22]. Thus, a more general comparison or review is needed. Secondly, there is no gold standard to compare these methods in real-world data applications since true local genetic correlations or significantly correlated blocks between phenotypic pairs are unknown. Even though we have conducted PRS analysis to help assess the performances of these methods, other downstream analyses can be done to compare the performance of different methods.

## Data Availability

All data produced in the present work are contained in the manuscript

## 5 Acknowledgments

We conducted the research using the UKBB resource under approved data requests (access ref: 29900). This study makes use of summary statistics from many GWAS consortia. We thank the investigators in these GWAS consortia for generously sharing their data. We are grateful to all the families participating in the Simons Foundation Powering Autism Research for Knowledge (SPARK) study.

## Notes

### Competing Interest Statement

The authors have declared no competing interest.

### Funding Statement

This study was funded by NIH grants R01 GM134005 and HG012735.

### Author Declarations

The North West Multi-Centre Research Ethics Committee (MREC) of UK Biobank gave ethical approval for this work.

