## Supplementary Material for "Benchmarking local genetic correlation estimation methods using summary statistics from genome-wide association studies"

### 1. Supplementary Figures

#### Supplementary Figure 1

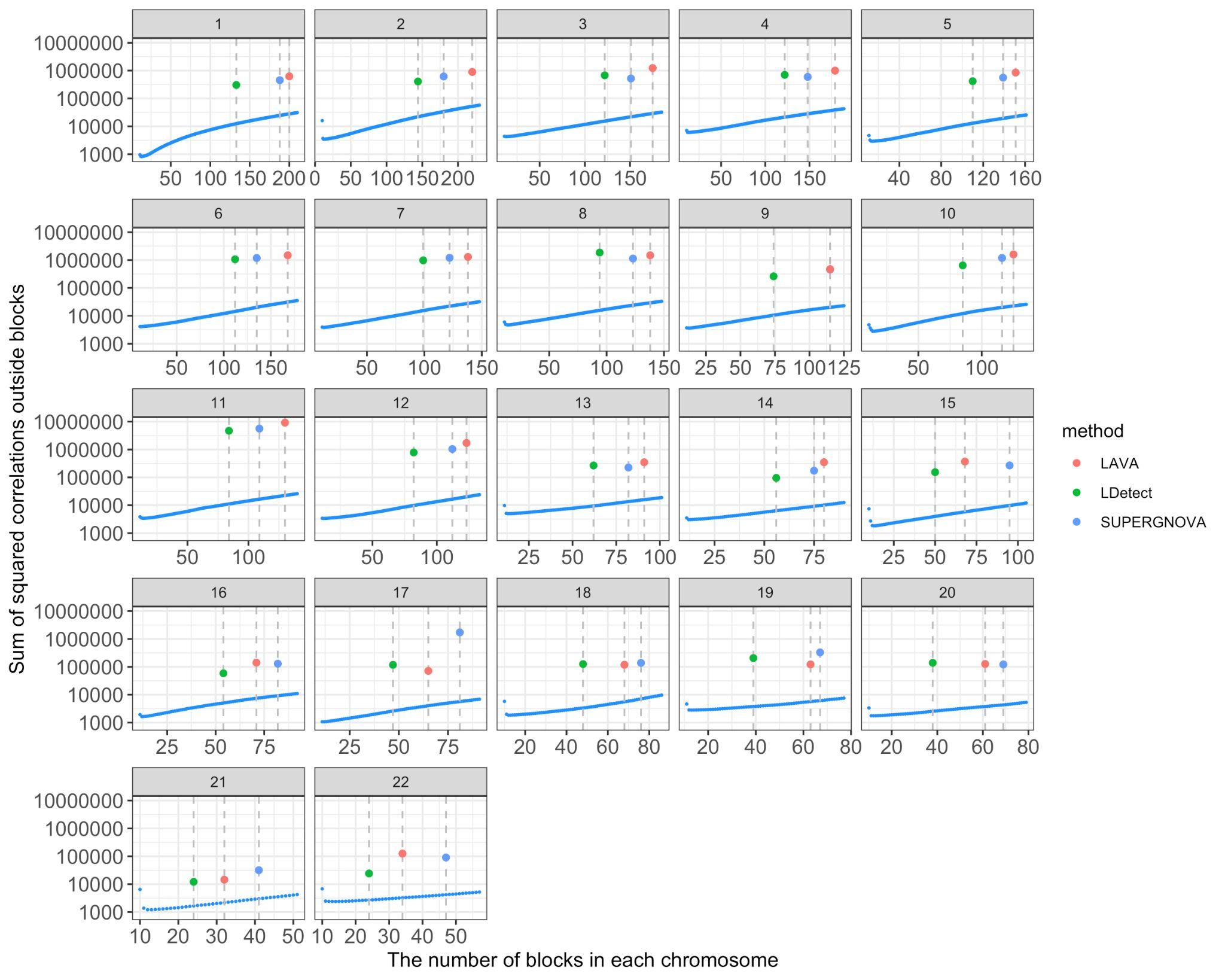

**Supplementary Figure 1: The sum of squared correlations between SNPs from different blocks for different partitions.** The red points represent the sum of the squared correlation using the partition provided by LDetect, SUPERGNOVA, and LAVA. The blue points represent the sum of squared correlation using partition generated by snp_ldsplit with different settings of the number of blocks.

#### Supplementary Figure 2

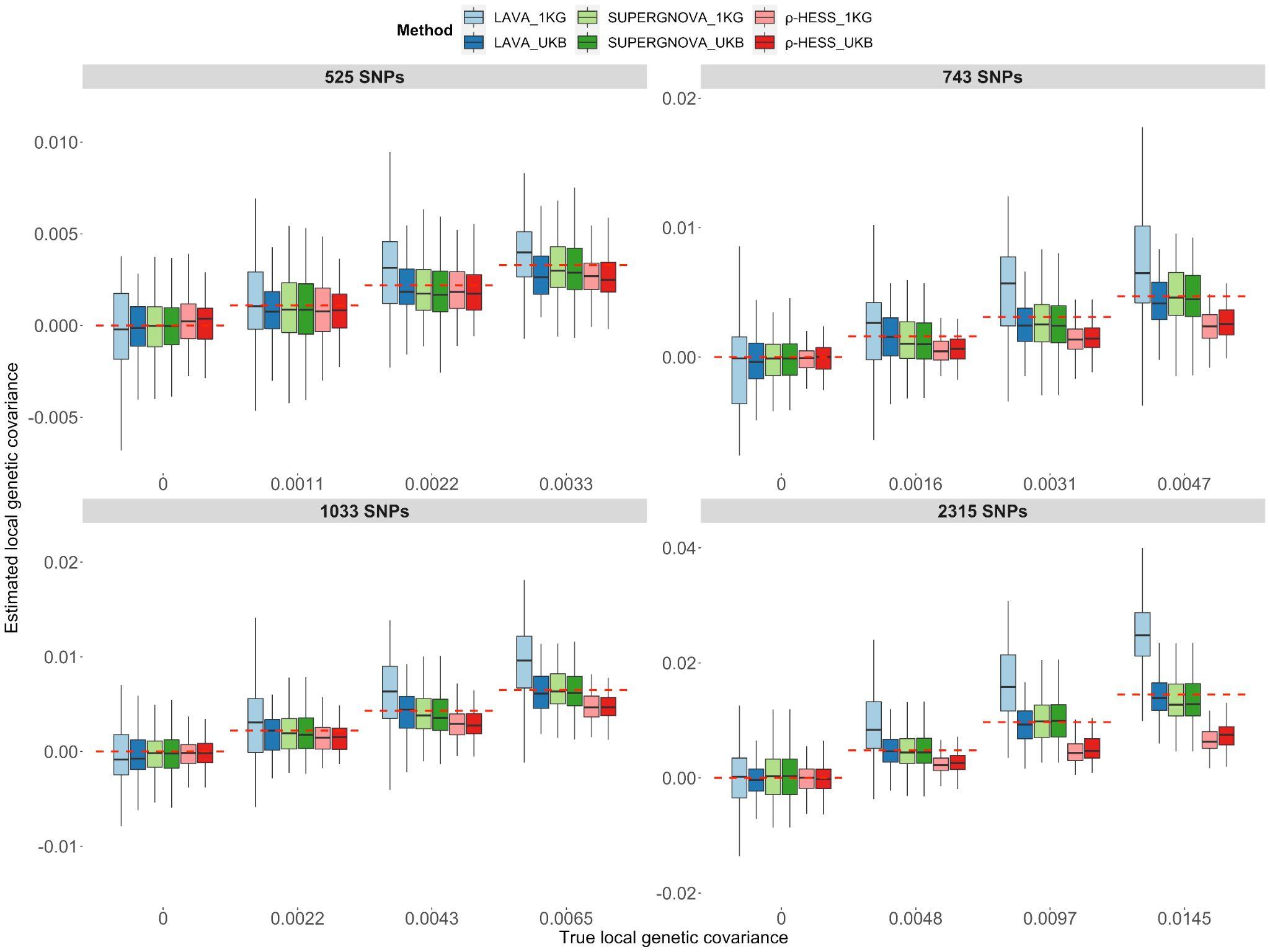

-

**Supplementary Figure 2: Evaluation of local genetic correlation/covariance methods on continuous phenotypes from non-overlapping datasets (set1 and set2) using an external reference panel with matched ancestry (EUR 1KG Phase 3) and an in-sample UKB reference panel (set1).** Local genetic covariance estimates on different loci. The red dashed lines represent the true value of local genetic covariances. Each panel represents the results of the loci with different numbers of SNPs.

#### Supplementary Figure 3

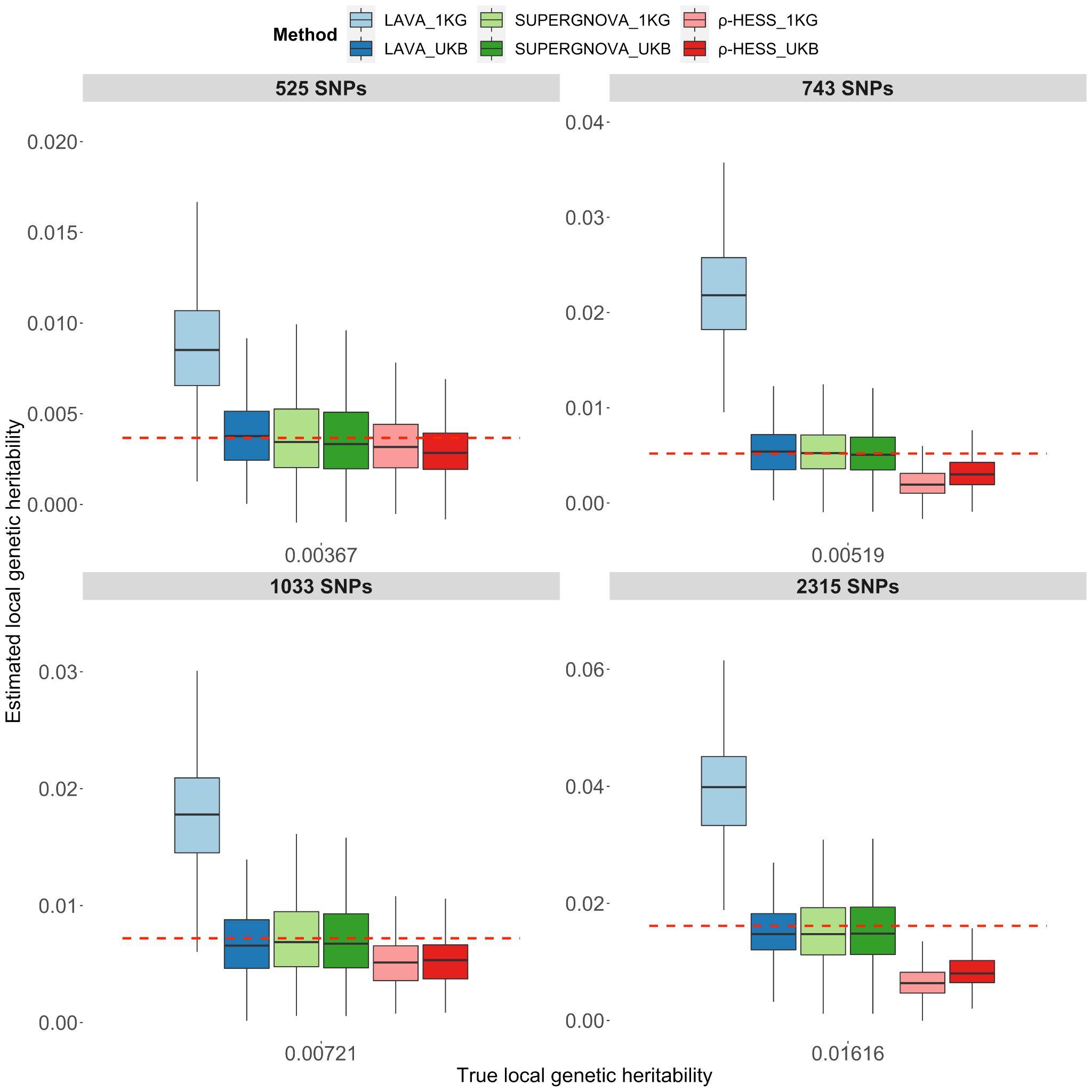

**Supplementary Figure 3: Evaluation of local genetic correlation/covariance methods on continuous phenotypes from non-overlapping datasets (set1 and set2) using an external reference panel with matched ancestry(EUR 1KG Phase 3) and an in-sample UKB reference panels(set1).** Local heritability estimation on different loci. The red dashed lines represent the true value of local heritability. Each panel represents the results of the loci with different numbers of SNPs.

#### Supplementary Figure 4

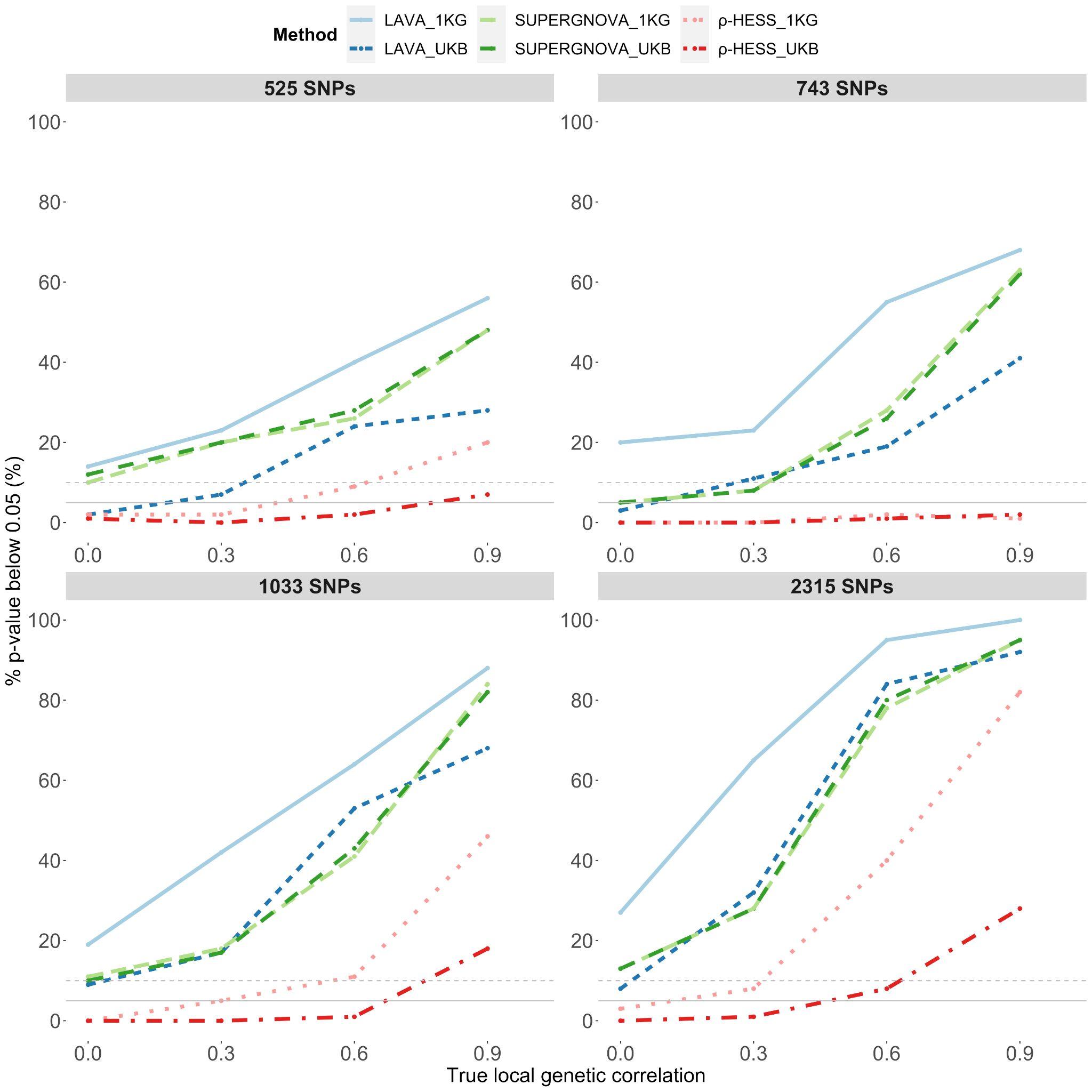

**Supplementary Figure 4: Evaluation of local genetic correlation/covariance methods on continuous phenotypes from non-overlapping datasets (set1 and set2) using an external reference panel with matched ancestry (EUR 1KG Phase 3) and an in-sample UKB reference panels(set1).** Type-1 Error and power on different loci. The grey solid line represents 5% and the grey dashed lines represent 10%. Each panel represents the results of the loci with different numbers of SNPs.

#### Supplementary Figure 5

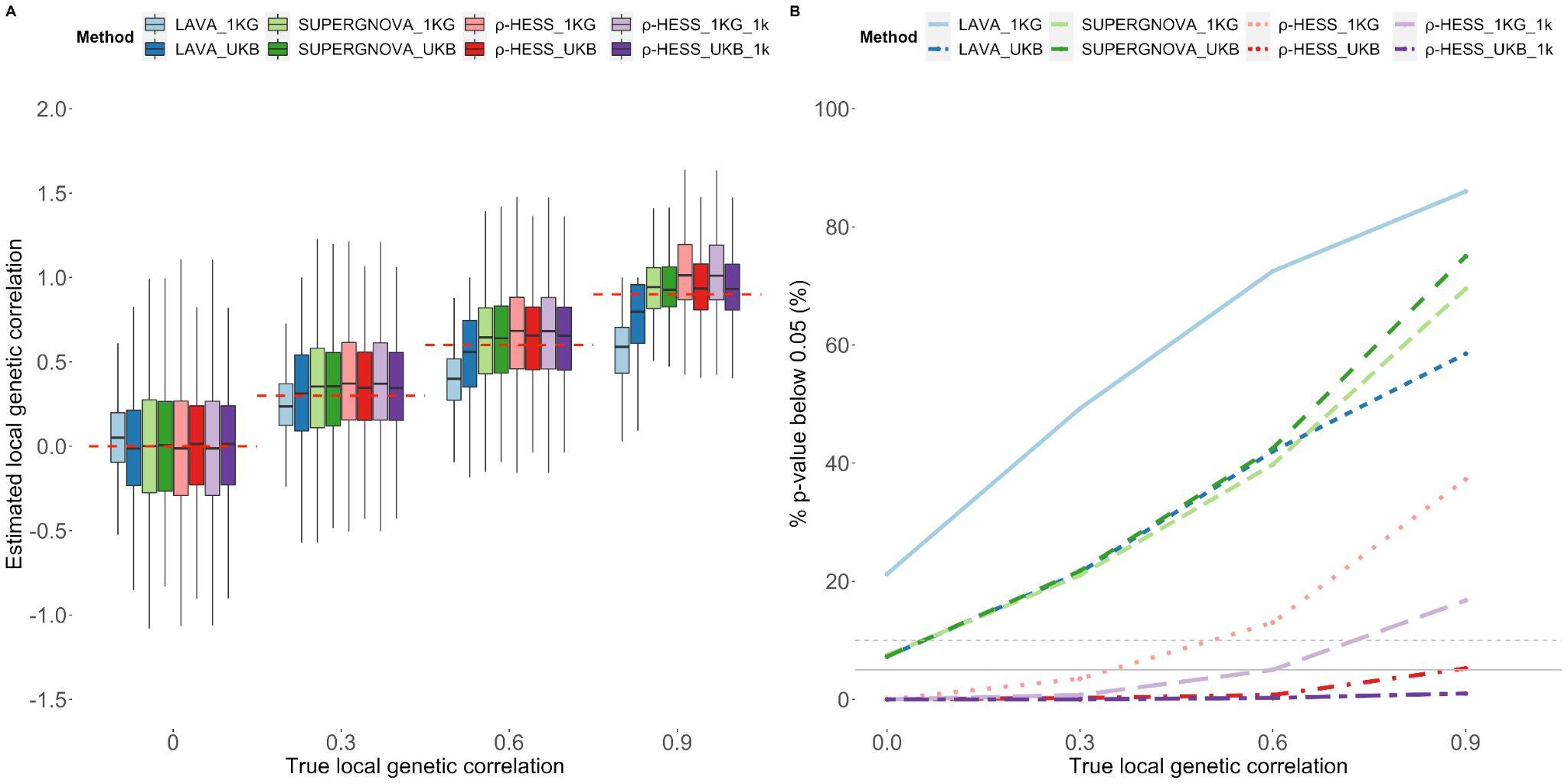

**Supplementary Figure 5: Evaluation of local genetic correlation/covariance methods on continuous phenotypes from partial-overlapping datasets (set1 and set3) using an external reference panel with matched ancestry (EUR 1KG Phase 3) and an in-sample UKB reference panel (set1). A.** Local genetic correlation estimates for different methods and different reference panels. **B**. Type-1 error and power for different methods and different reference panels. When using $\rho$-hess, we gave it either a true overlap sample size ($\rho$-HESS_1KG and $\rho$-HESS_UKB) or a wrong overlap sample size ($\rho$-HESS _1KG_1k and $\rho$-HESS _UKB_1k). The red dashed lines represent the true value of local genetic correlation.

#### Supplementary Figure 6

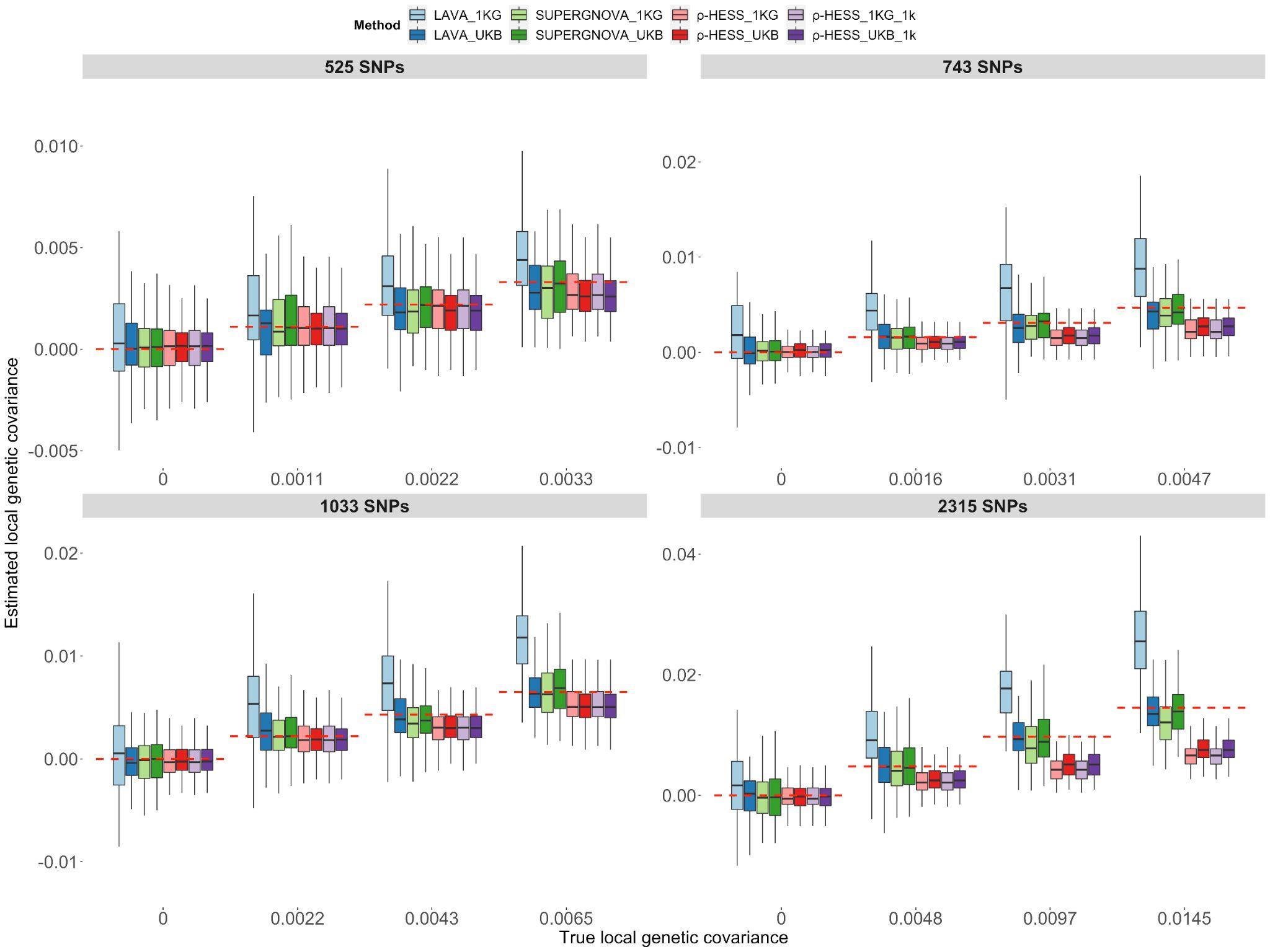

**Supplementary Figure 6: Evaluation of local genetic correlation/covariance methods on continuous phenotypes from partial-overlapping datasets (set1 and set3) using an external reference panel with matched ancestry(EUR 1KG Phase 3) and an in-sample UKB reference panel (set1).** Local genetic covariance estimates on different loci. The red dashed lines represent the true value of local genetic covariances. Each panel represents the results of the loci with different numbers of SNPs.

#### Supplementary Figure 7

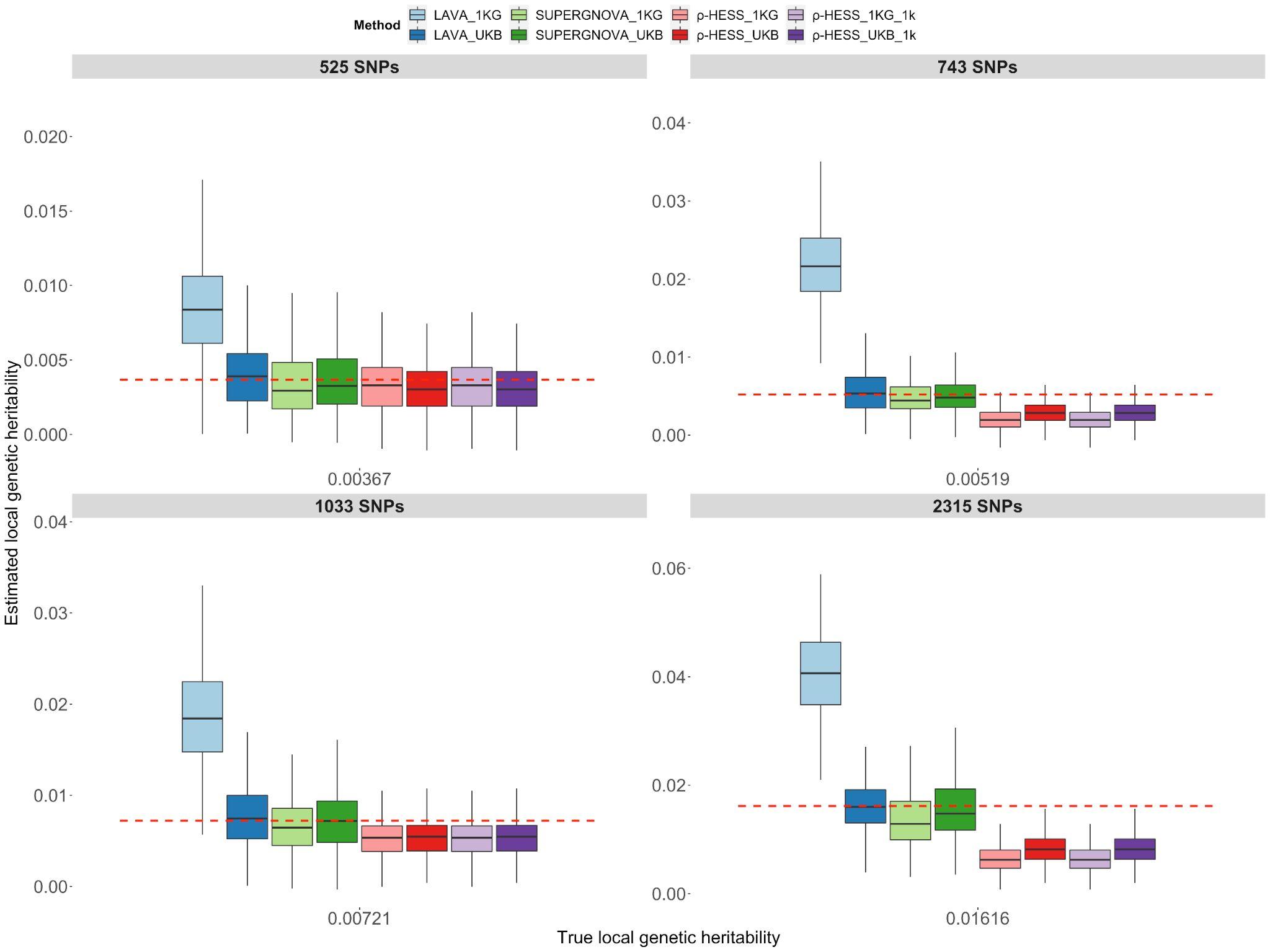

**Supplementary Figure 7: Evaluation of local genetic correlation/covariance methods on continuous phenotypes from partial-overlapping datasets (set1 and set3) using an external reference panel with matched ancestry(EUR 1KG Phase 3) and an in-sample UKB reference panel (set1).** Local heritability on different loci. The red dashed lines represent the true value of local heritability. Each panel represents the results of the loci with different numbers of SNPs.

#### Supplementary Figure 8

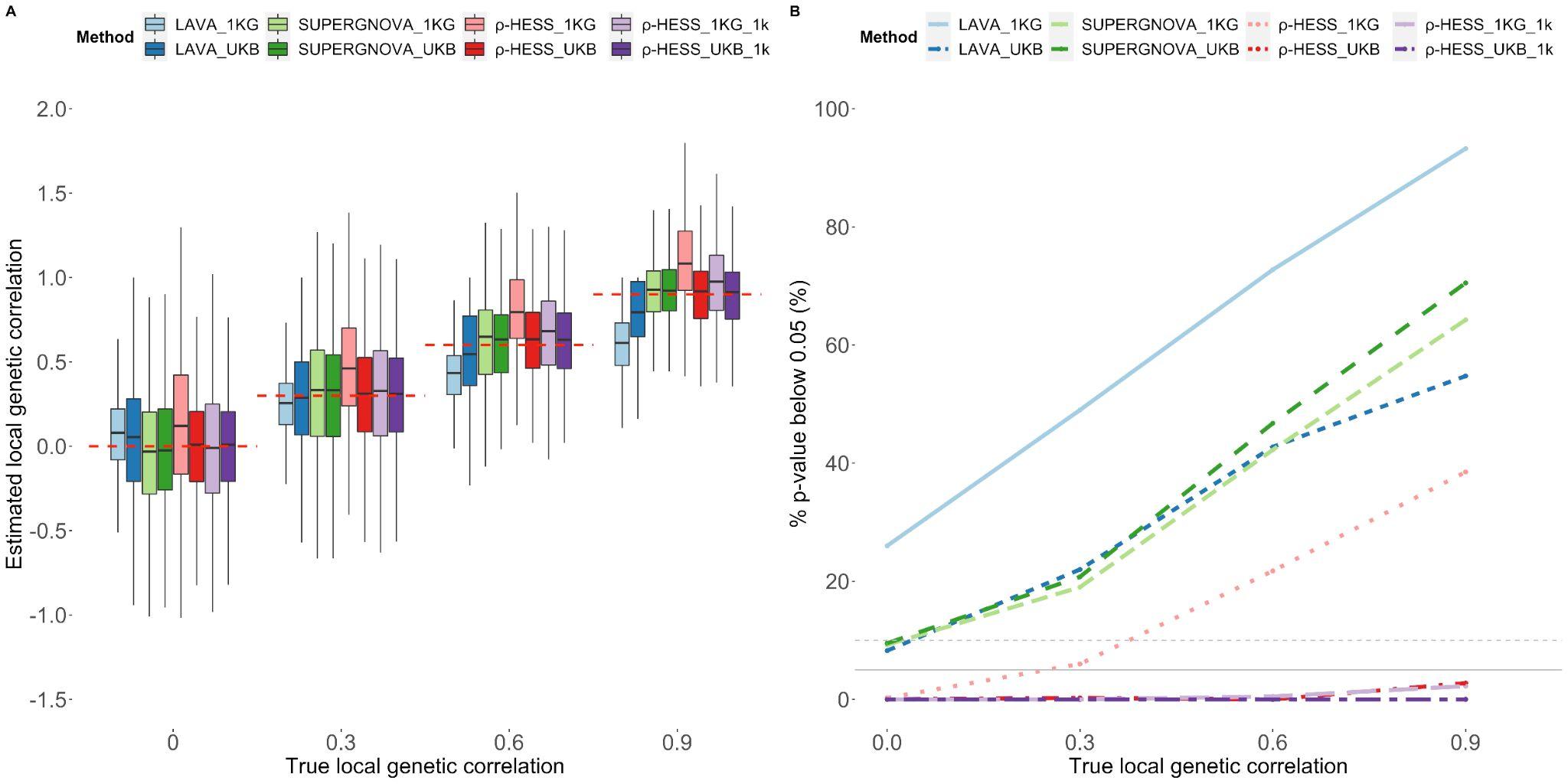

**Supplementary Figure 8: Evaluation of local genetic correlation/covariance methods on continuous phenotypes from completely overlapping datasets (set1 and set1) using an external reference panel with matched ancestry(EUR 1KG Phase 3) and an in-sample UKB reference panel (set1). A.** Local genetic correlation estimates for different methods and different reference panels. **B**. Type-1 error and power for different methods and different reference panels. When using p-hess, we gave it either a true overlap sample size ($\rho$-HESS_1KG and $\rho$-HESS_UKB) or a wrong overlap sample size($\rho$-HESS_1KG_1k and $\rho$-HESS_UKB_1k). The red dashed lines represent the true value of local genetic correlation.

#### Supplementary Figure 9

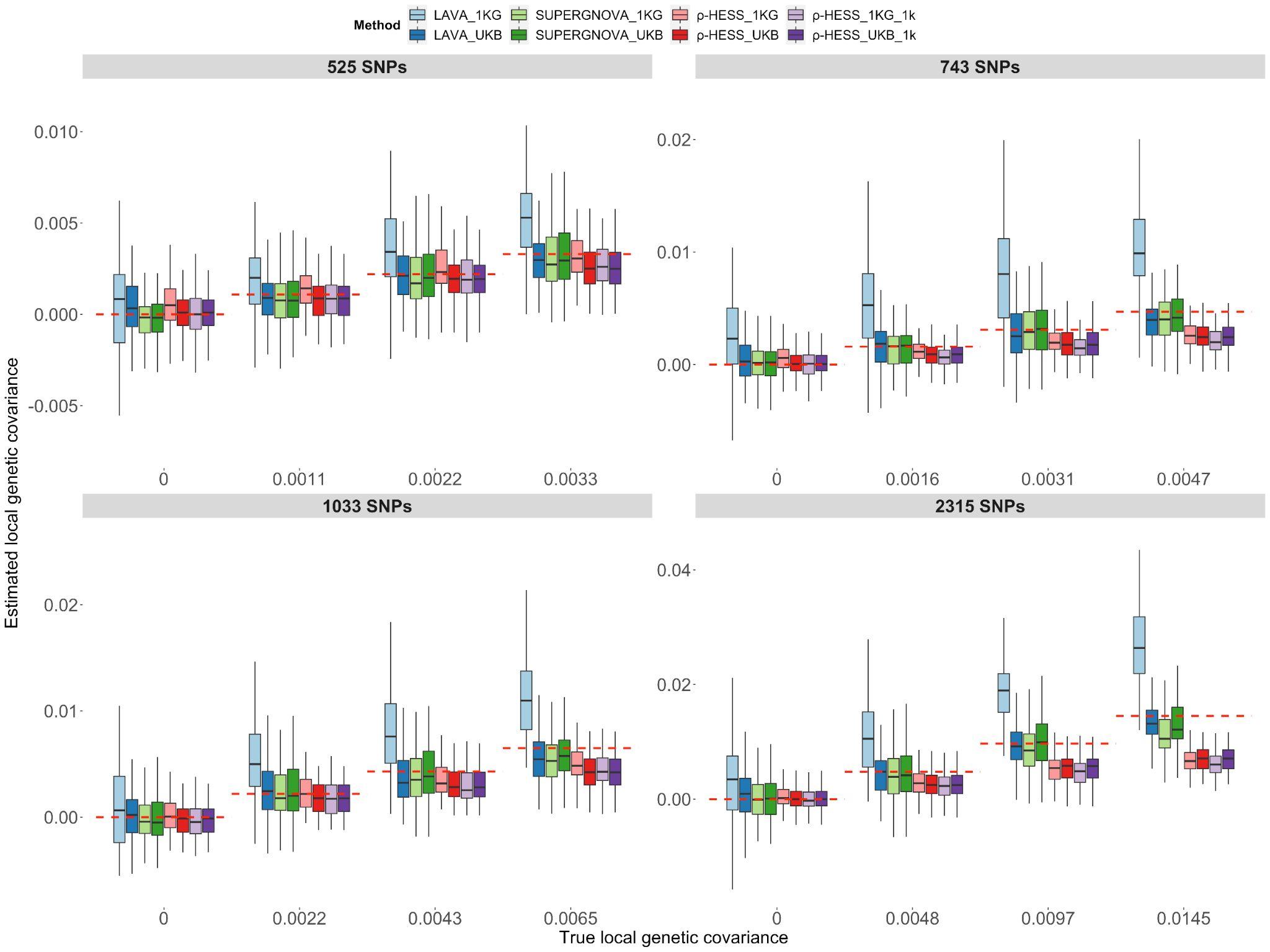

**Supplementary Figure 9: Evaluation of local genetic correlation/covariance methods on continuous phenotypes from completely overlapping datasets (set1 and set1) using an external reference panel with matched ancestry(EUR 1KG Phase 3) and an in-sample UKB reference panel (set1).** Local genetic covariance estimates on different loci. The red dashed lines represent the true value of local genetic covariances. Each panel represents the results of the loci with different numbers of SNPs.

#### Supplementary Figure 10

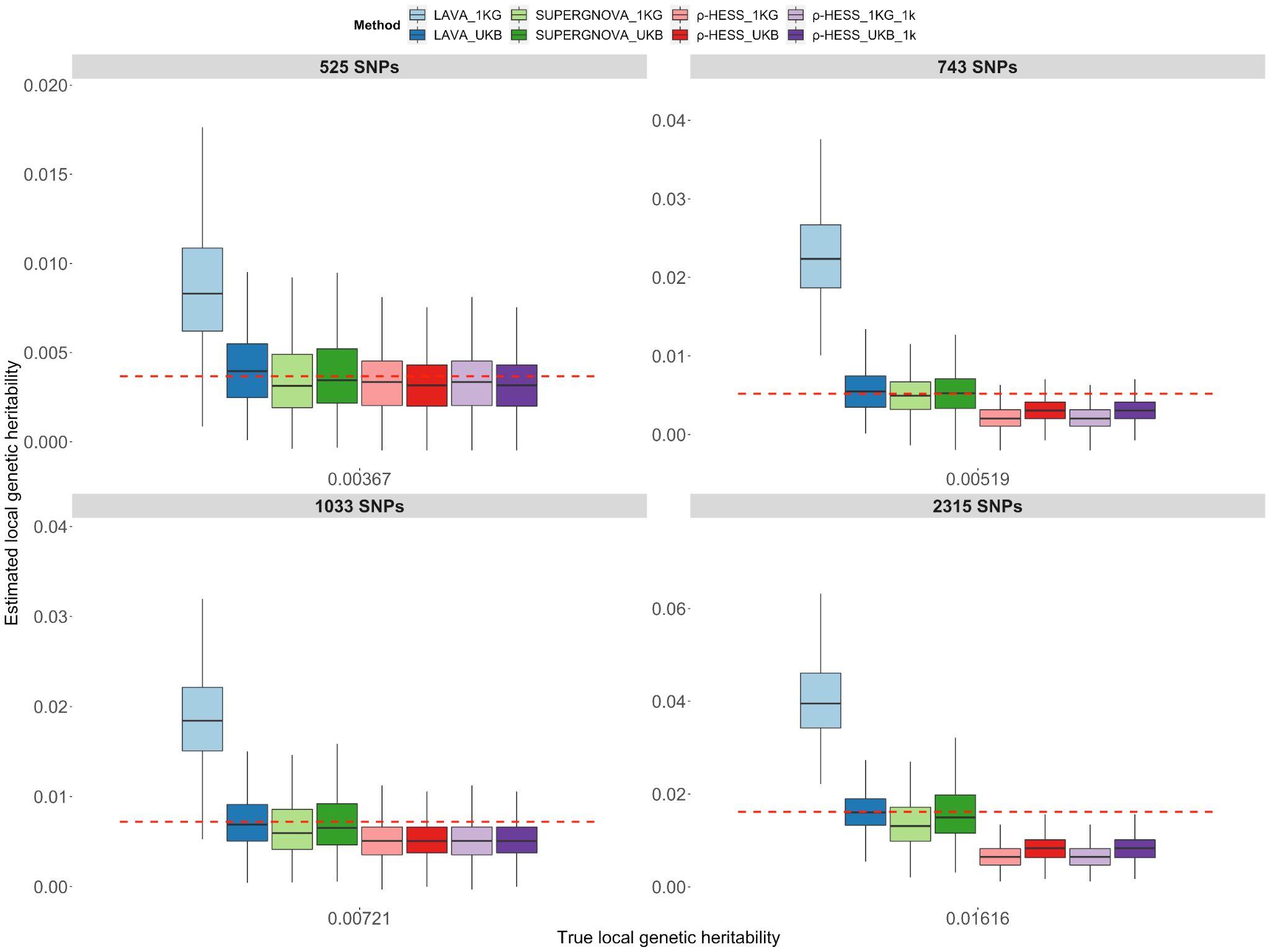

**Supplementary Figure 10: Evaluation of local genetic correlation/covariance methods on continuous phenotypes from completely overlapping datasets (set1 and set1) using an external reference panel with matched ancestry(EUR 1KG Phase 3) and an in-sample UKB reference panel (set1).** Local heritability estimates on different loci. The red dashed lines represent the true value of local heritability. Each panel represents the results of the loci with different numbers of SNPs.

#### Supplementary Figure 11

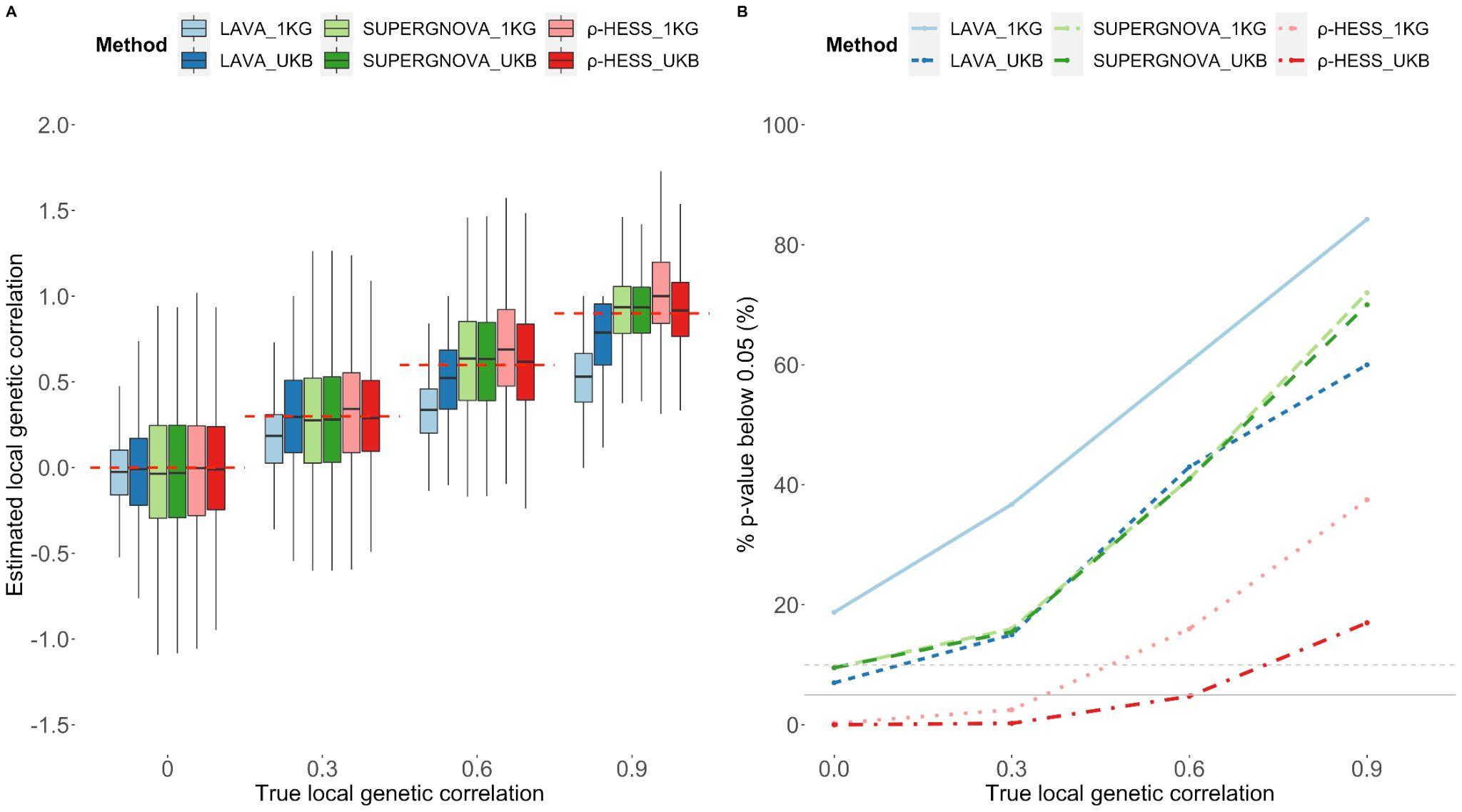

**Supplementary Figure 11: Evaluation of local genetic correlation/covariance methods when only a fraction of SNPs were chosen as causal SNPs (20%) for continuous phenotypes from non-overlapping datasets (set1 and set2) using an external reference panel with matched ancestry( EUR 1KG Phase 3) and an in-sample UKB reference panel (set1). A.** Local genetic correlation estimates for different methods and different reference panels. **B**. Type-1 error and power for different methods and different reference panels. When using $\rho$-hess, we gave it either a true overlap sample size($\rho$-HESS_1KG and $\rho$-HESS_UKB) or a wrong overlap sample size ($\rho$-HESS_1KG_1k and $\rho$-HESS_UKB_1k). The red dashed lines represent the true value of local genetic correlation.

#### Supplementary Figure 12

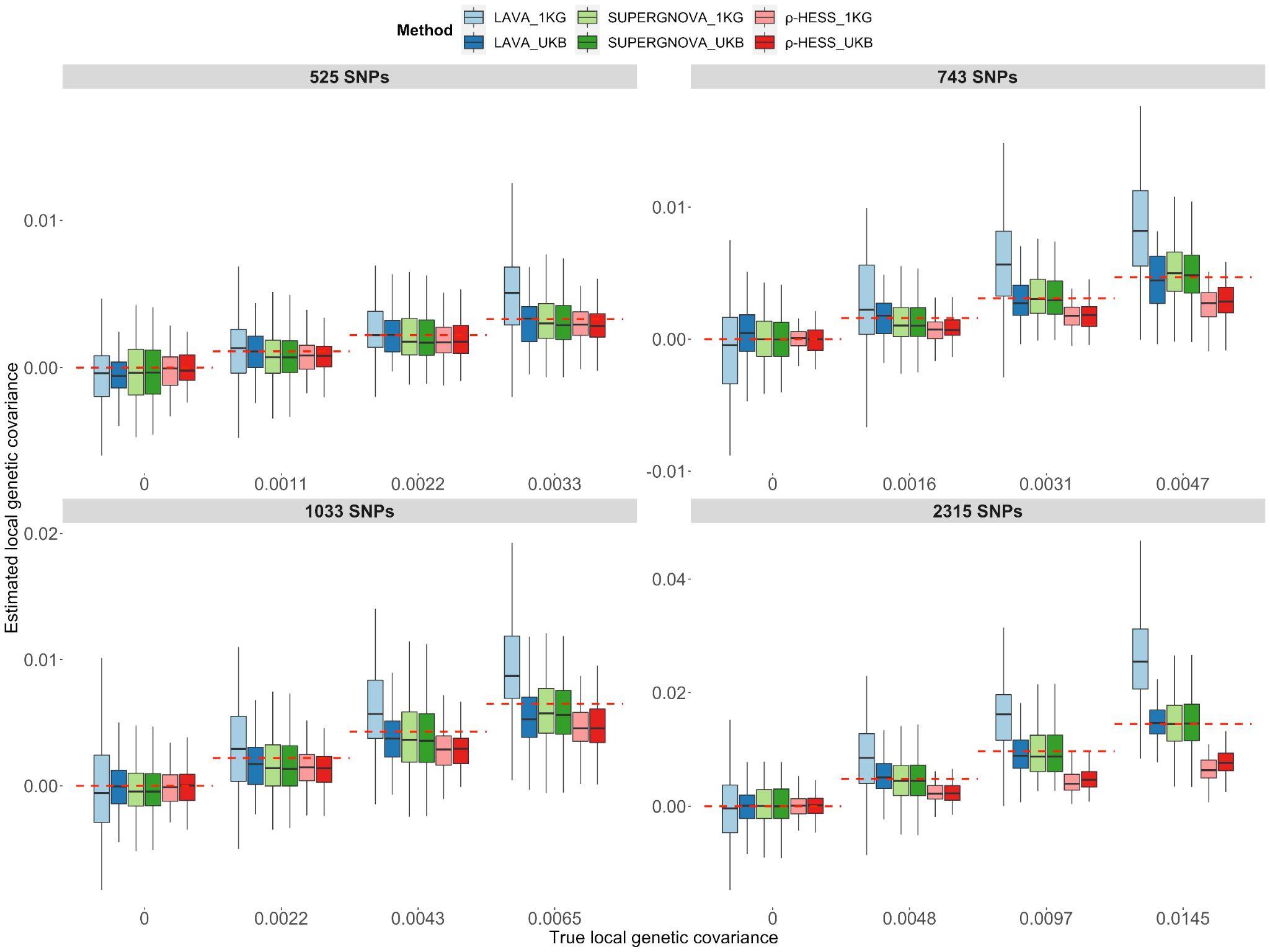

**Supplementary Figure 12: Evaluation of local genetic correlation/covariance methods when only a fraction of SNPs were chosen as causal SNPs (20%) for continuous phenotypes from non-overlapping datasets (set1 and set2) using an external reference panel with matched ancestry( EUR 1KG Phase 3) and an in-sample UKB reference panel (set1).**Local genetic covariance estimates on different loci. The red dashed lines represent the true value of local genetic covariance.Each panel represents the results of the loci with different numbers of SNPs.

#### Supplementary Figure 13

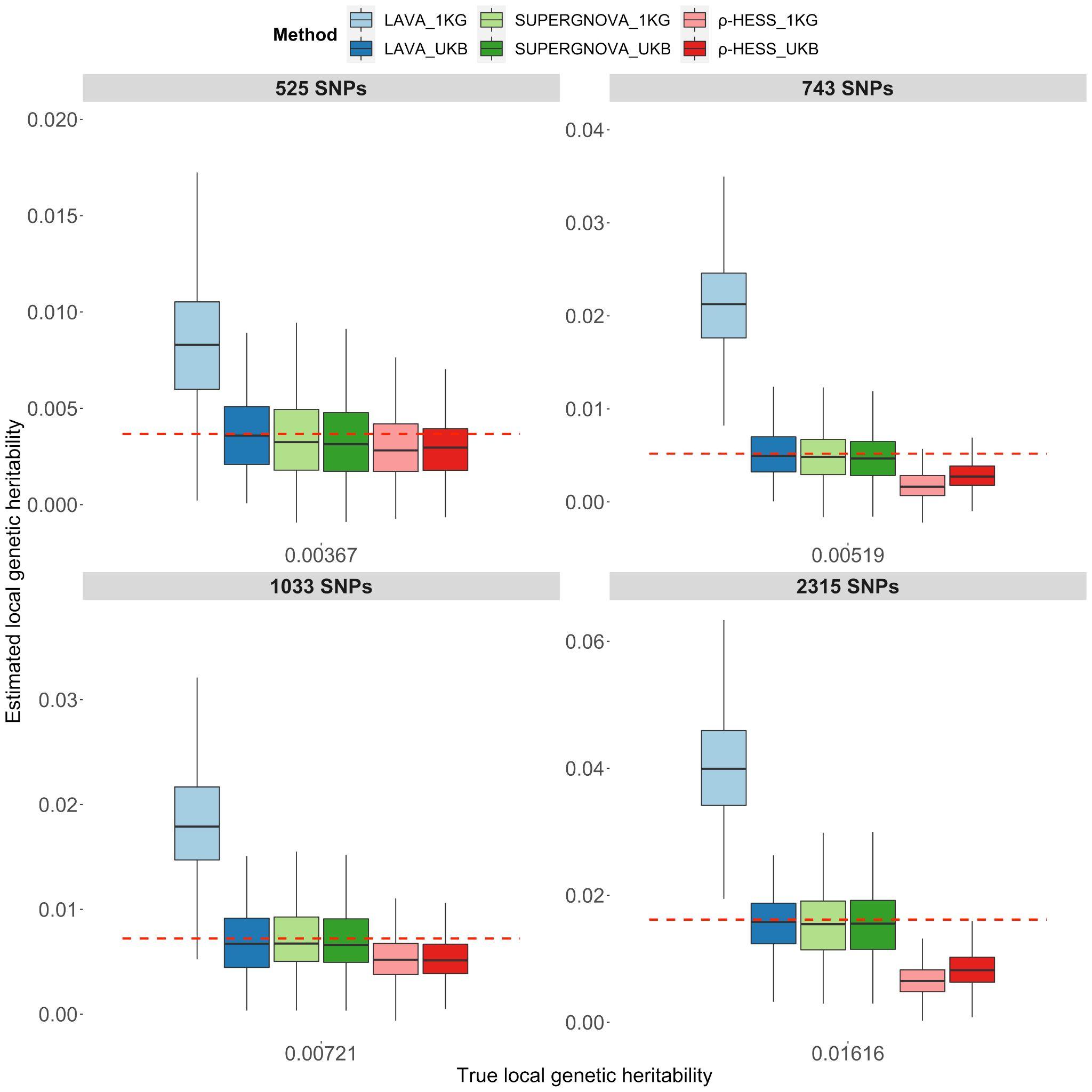

**Supplementary Figure 13: Evaluation of local genetic correlation/covariance methods when only a fraction of SNPs were chosen as causal SNPs (20%) for continuous phenotypes from non-overlapping datasets (set1 and set2) using an external reference panel with matched ancestry( EUR 1KG Phase 3) and an in-sample UKB reference panel (set1).**Local heritability estimates on different loci. The red dashed lines represent the true value of local heritability. Each panel represents the results of the loci with different numbers of SNPs.

#### Supplementary Figure 14

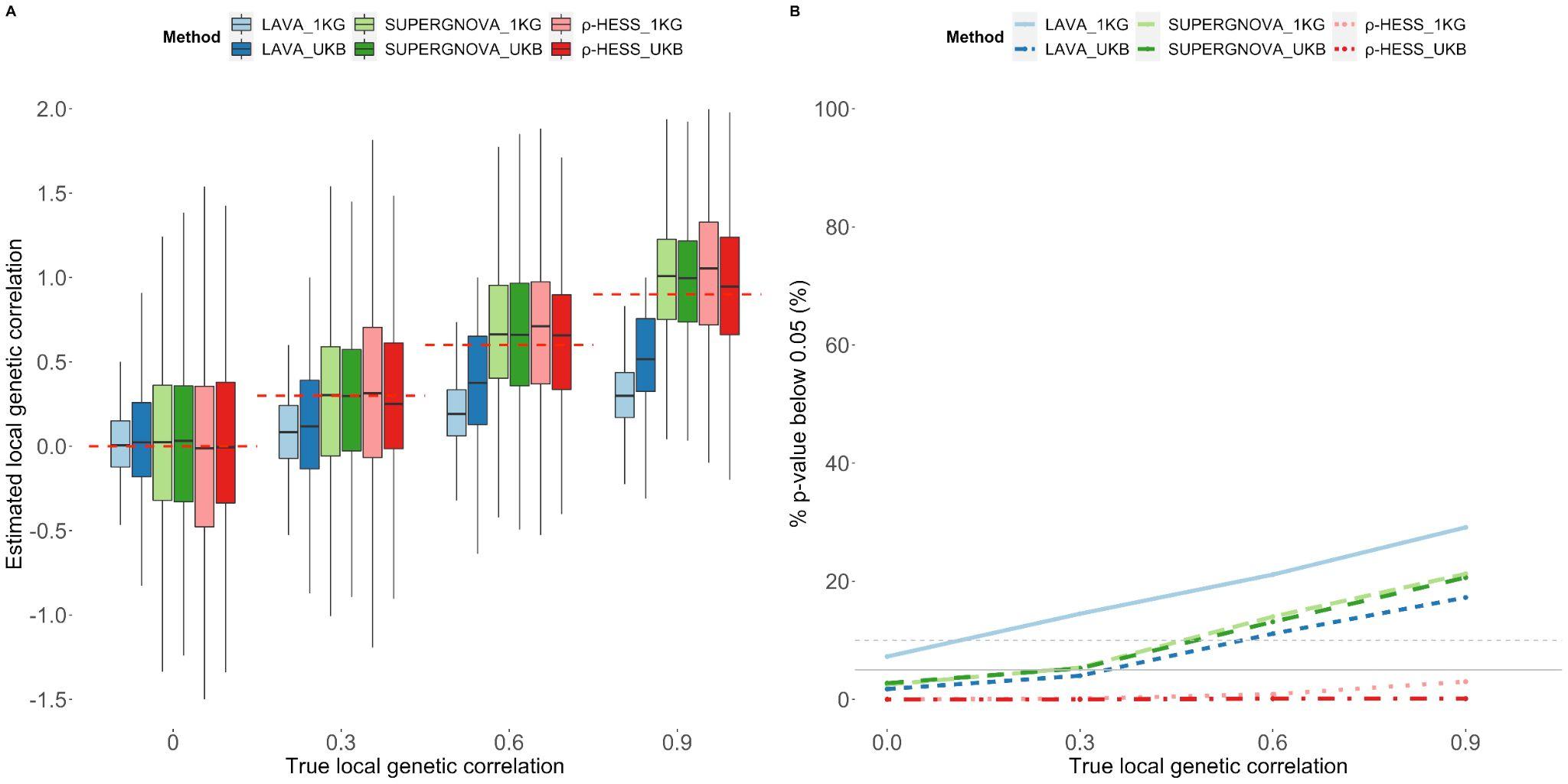

**Supplementary Figure 14: Evaluation of local genetic correlation/covariance methods for binary phenotypes whose prevalence is 0.2 from non-overlapping datasets (set1 and set2) using an external reference panel with matched ancestry (EUR 1KG Phase 3) and an in-sample UKB reference panel (set1). A.** Local genetic correlation estimates for different methods and different reference panels. **B**. Type-1 error and power for different methods and different reference panels.

#### Supplementary Figure 15

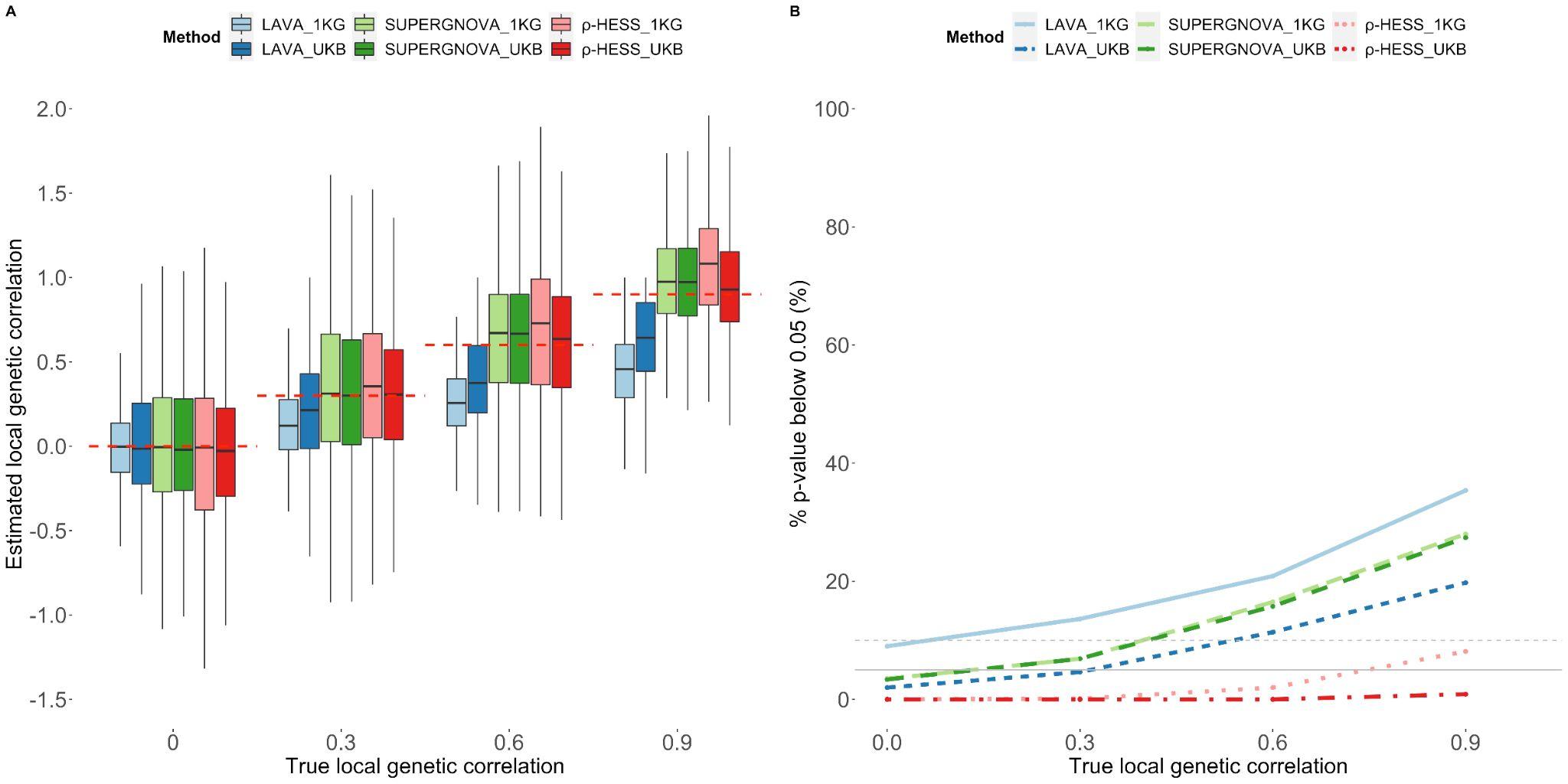

**Supplementary Figure 15: Evaluation of local genetic correlation/covariance methods for binary phenotypes whose prevalence is 0.5 from non-overlapping datasets (set1 and set2) using an external reference panel with matched ancestry(EUR 1KG Phase 3) and an in-sample UKB reference panel (set1). A.** Local genetic correlation estimates for different methods and different reference panels. **B**. Type-1 error and power for different methods and different reference panels.

#### Supplementary Figure 16

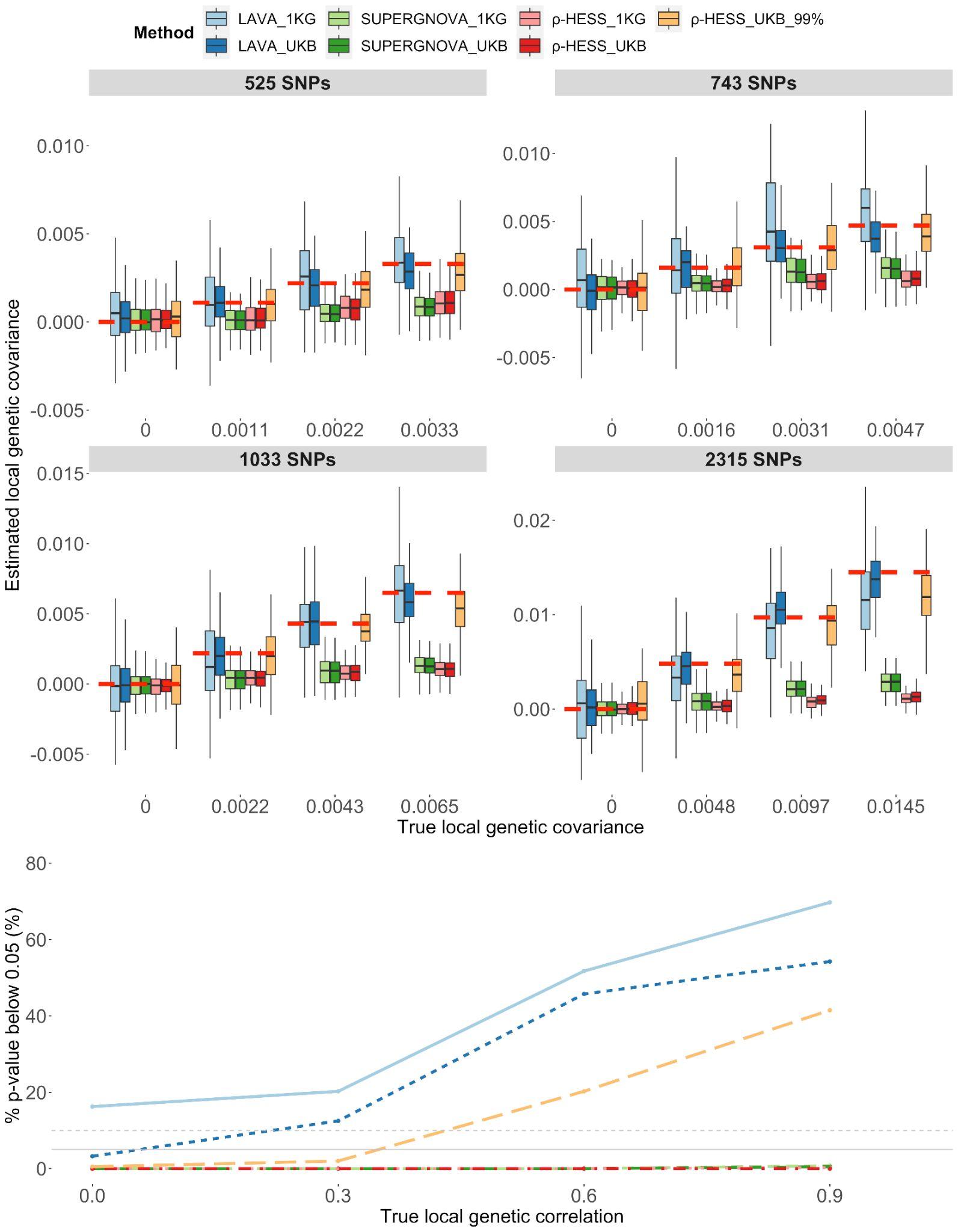

**Supplementary Figure 16: Evaluation of SUPERGNOVA when the effect sizes associated with local LD matrix for continuous phenotypes from non-overlapping datasets (set1 and set2) using the UKB reference panel (set1 and set2). A.** Local genetic covariance estimates. The red dashed lines represent the true value of local genetic covariance. **B**. Type-1 error and power. The grey solid lines represent 5% and the grey dashed lines represent 10%.

#### Supplementary Figure 17

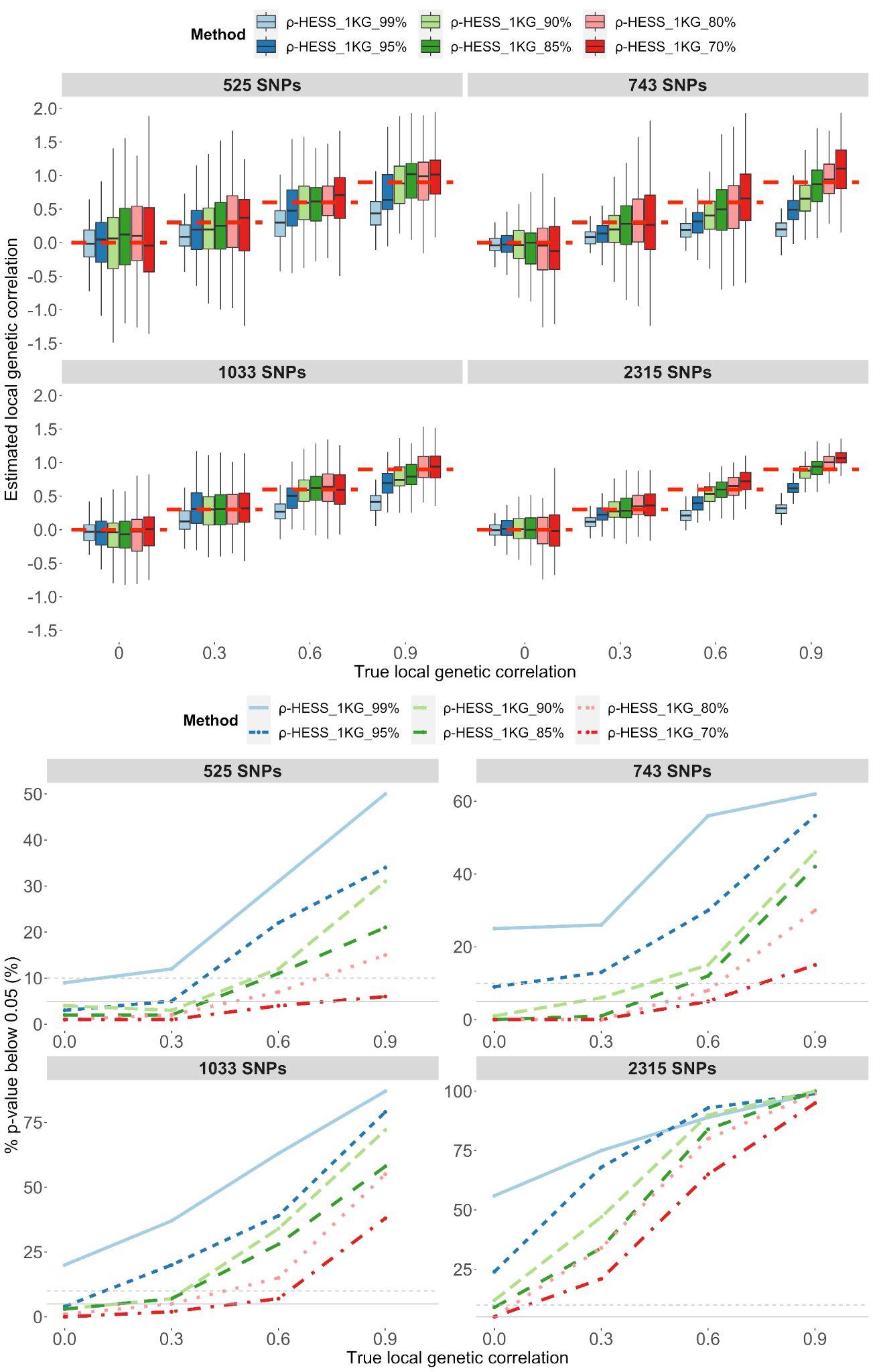

**Supplementary Figure 17: Evaluation of** $\boldsymbol{\rho}$**-hess using the different number of eigenvalues for continuous phenotypes from non-overlapping datasets (set1 and set2) using EUR 1KG Phase 3 reference panel. A.** Local genetic correlation estimates. The red dashed lines represent the true value of local genetic correlation. **B**. Type-1 error and power. The grey solid lines represent 5% and the grey dashed lines represent 10%.

#### Supplementary Figure 18

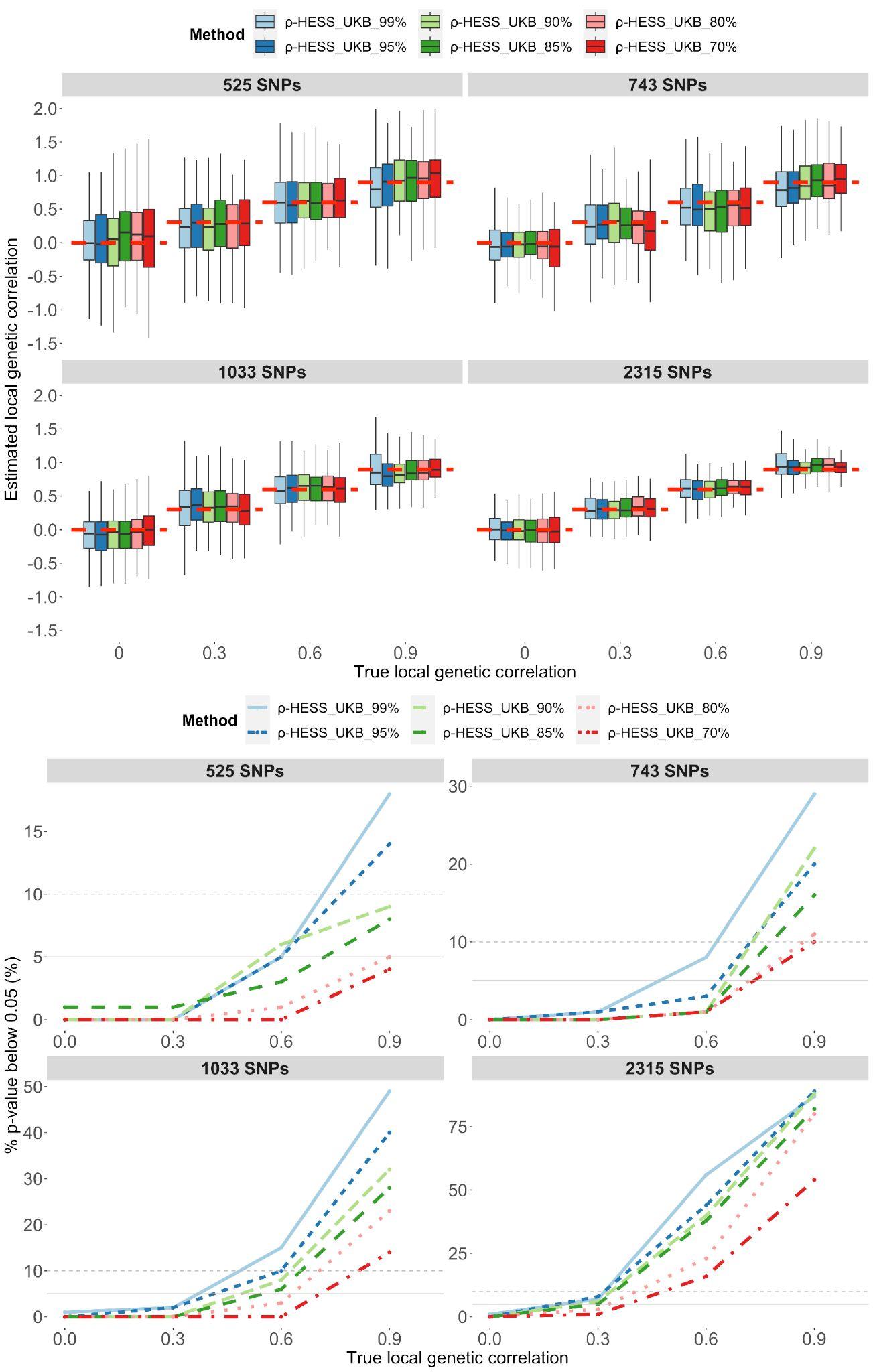

**Supplementary Figure 18: Evaluation of** $\boldsymbol{\rho}$**-hess using the different number of eigenvalues for continuous phenotypes from non-overlapping datasets (set1 and set2) using the UKB reference panel (set1 and set2). A.** Local genetic correlation estimates. The red dashed lines represent the true value of local genetic correlation. **B**. Type-1 error and power. The grey solid lines represent 5% and the grey dashed lines represent 10%.

#### Supplementary Figure 19

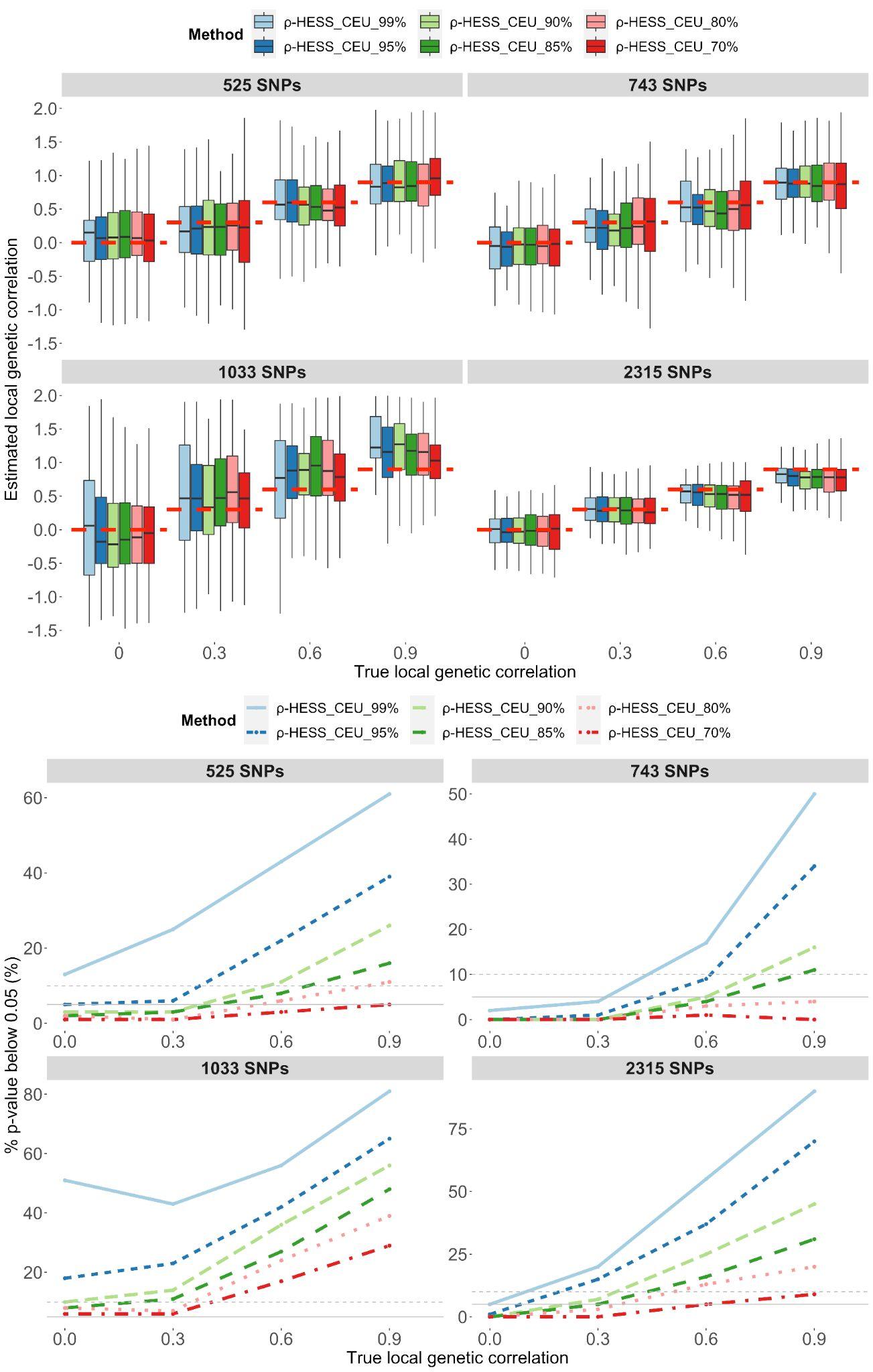

**Supplementary Figure 19: Evaluation of** $\boldsymbol{\rho}$**-hess using the different number of eigenvalues for continuous phenotypes from non-overlapping datasets (set1 and set2) using the CEU reference panel with 20,000 individuals. A.** Local genetic correlation estimates. The red dashed lines represent the true value of local genetic correlation. **B**. Type-1 error and power. The grey solid lines represent 5% and the grey dashed lines represent 10%.

#### Supplementary Figure 20

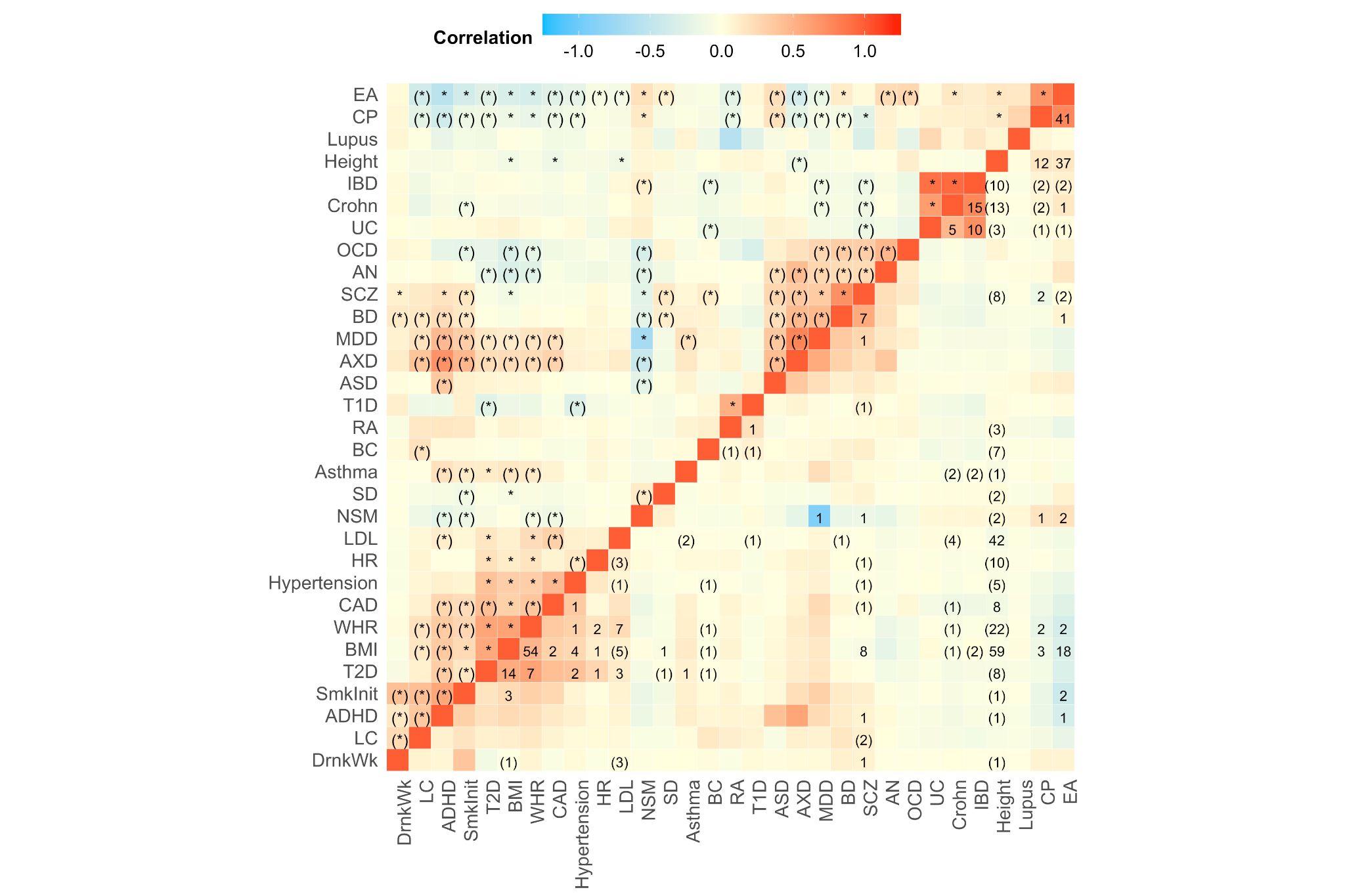

**Supplementary Figure 20: A comparison of the global genetic correlations calculated by LDSC and the mean local genetic correlations estimated by** $\boldsymbol{\rho}$**-hess for 31 phenotypes/465 pairs.** The global genetic correlations are displayed above the diagonal, with asterisks indicating a significant global genetic correlation estimated by LDSC (P= 0.05/465 = 1.07e-4). The mean local genetic correlations from $\rho$-HESS are displayed below the diagonal, with the numbers indicating the number of significant loci found (P =0.05/541,211 = 9.24e-8). Parentheses around an asterisk indicate that LDSC discovered a significant global correlation, but found no evidence of a significant local correlation, whereas parentheses around a number indicate that $\rho$-hess recognized at least one local genetic correlation despite no significant global genetic correlation.

#### Supplementary Figure 21

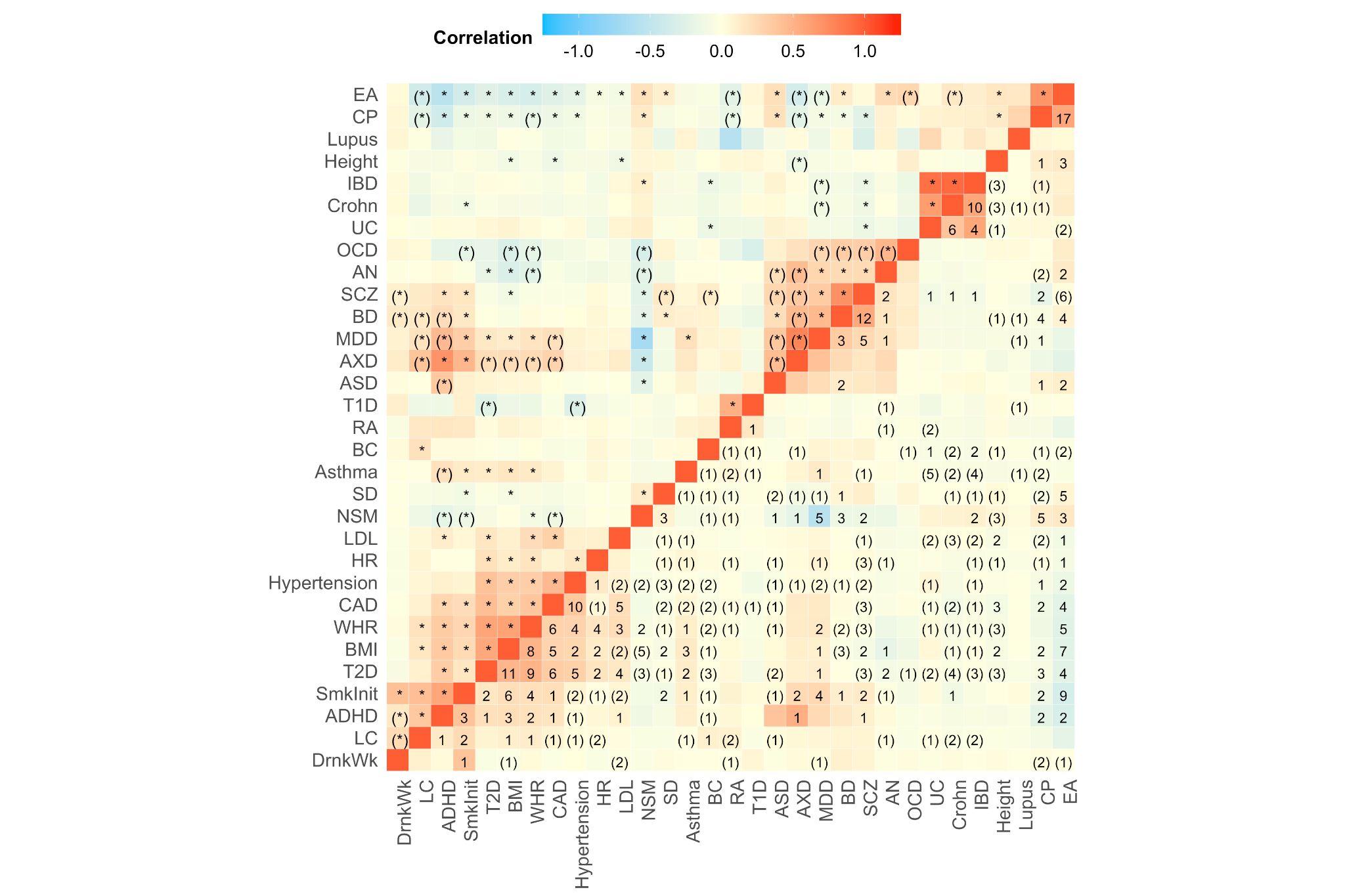

**Supplementary Figure 21: A comparison of the global genetic correlations calculated by LDSC and the mean local genetic correlations estimated by SUPERGNOVA for 31 phenotypes/465 pairs.** The global genetic correlations are displayed above the diagonal, with asterisks indicating a significant global genetic correlation estimated by LDSC (P= 0.05/465 = 1.07e-4). The mean local genetic correlations from SUPERGNOVA are displayed below the diagonal, with the numbers indicating the number of significant loci found (P =0.05/413,062 = 1.21e-7). Parentheses around an asterisk indicate that LDSC discovered a significant global correlation, but found no evidence of a significant local correlation, whereas parentheses around a number indicate that SUPERGNOVA recognized at least one local genetic correlation despite no significant global genetic correlation.

#### Supplementary Figure 22

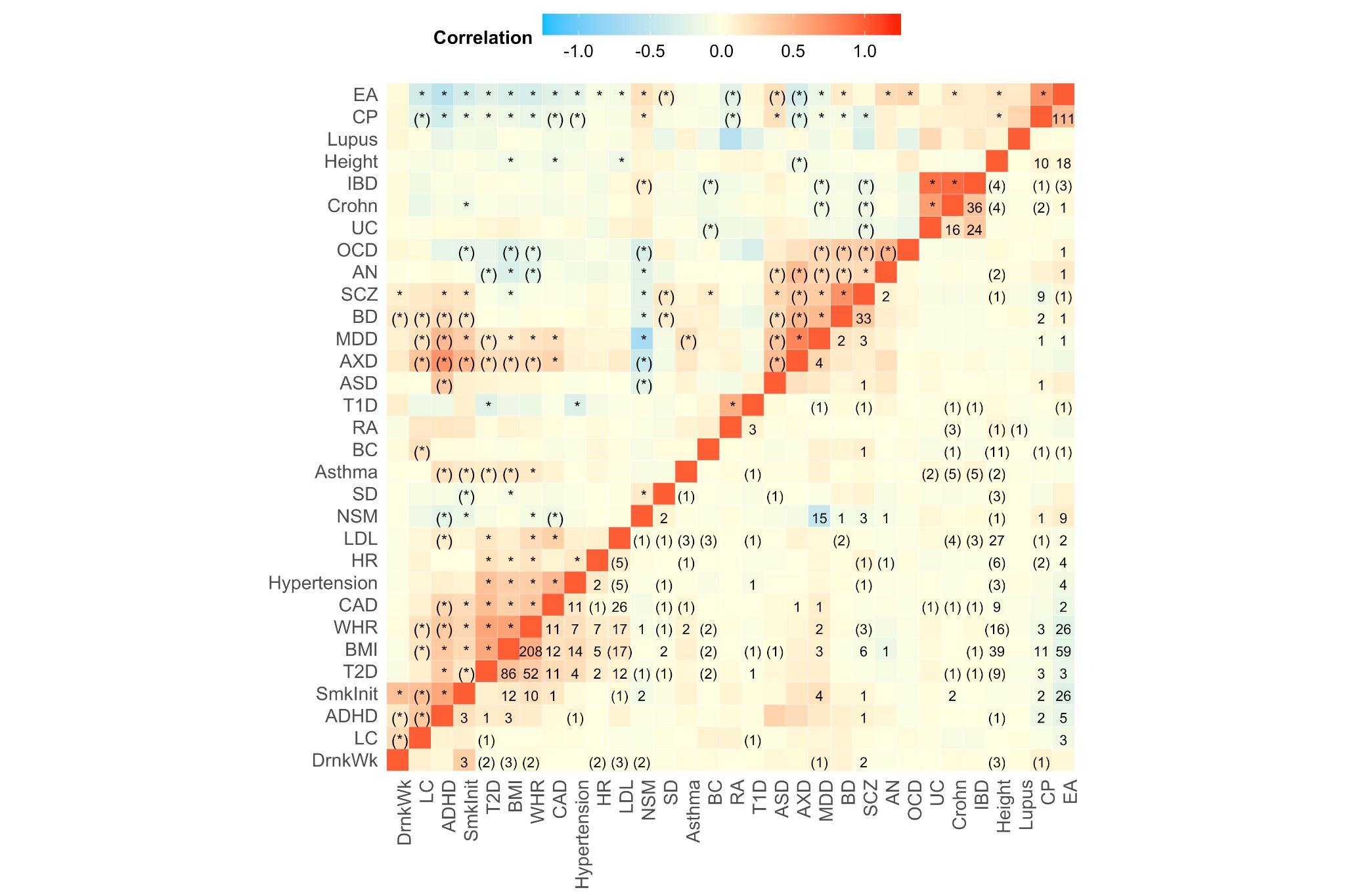

**Supplementary Figure 22: A comparison of the global genetic correlations calculated by LDSC and the mean local genetic correlations estimated by LAVA for 31 phenotypes/465 pairs.** The global genetic correlations are displayed above the diagonal, with asterisks indicating a significant global genetic correlation estimated by LDSC (P= 0.05/465 = 1.07e-4). The mean local genetic correlations from LAVA are displayed below the diagonal, with the numbers indicating the number of significant loci found (P =0.05/605,392 = 8.26e-8). Parentheses around an asterisk indicate that LDSC discovered a significant global correlation, but found no evidence of a significant local correlation, whereas parentheses around a number indicate that LAVA recognized at least one local genetic correlation despite no significant global genetic correlation.

#### Supplementary Figure 23

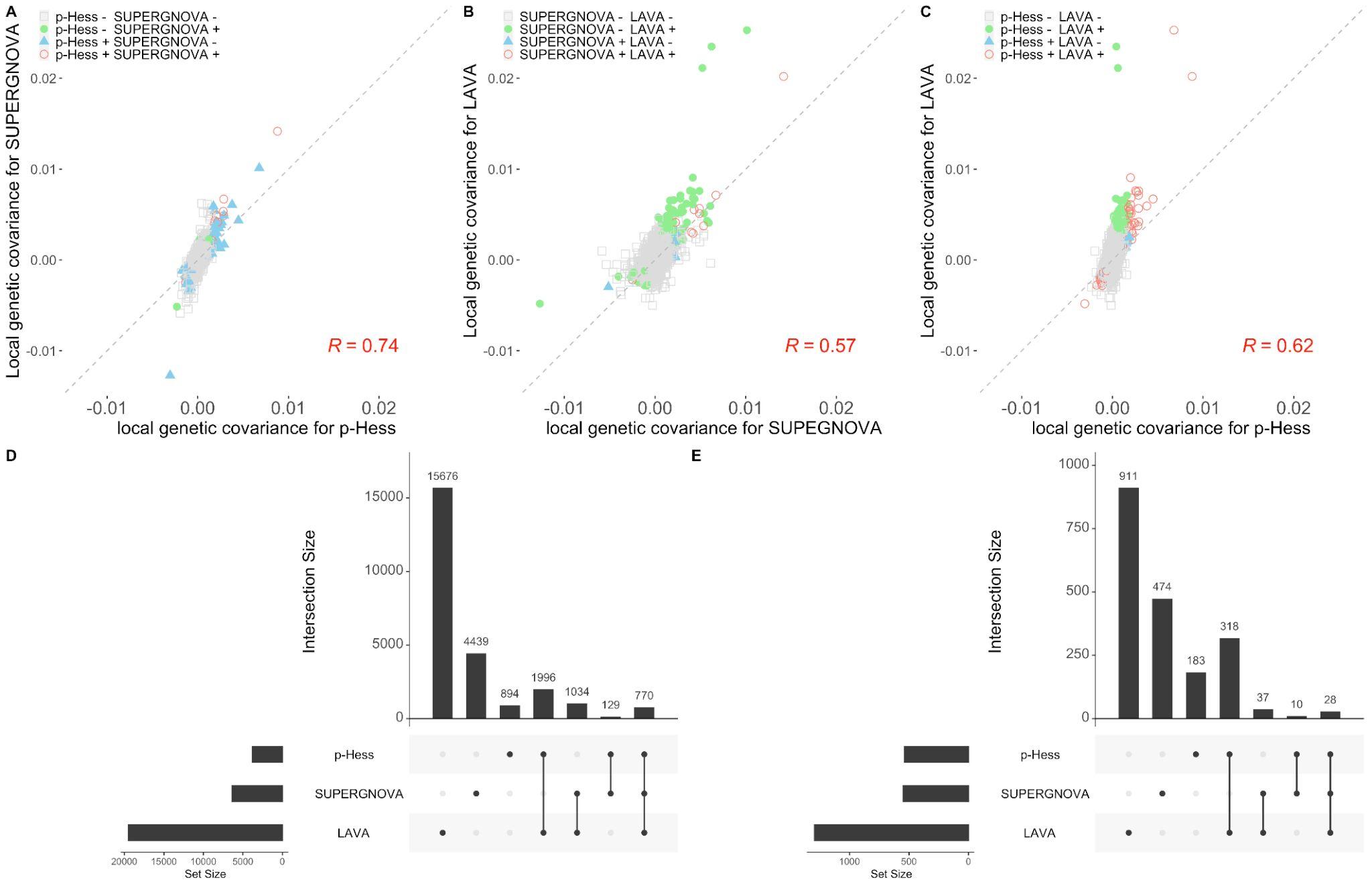

**Supplementary Figure 23: The comparison of local genetic covariances derived from** $\boldsymbol{\rho}$**-hess, SUPERGNOVA, and LAVA using EUR 1KG Phase3 reference panels across 2,048 blocks for 465 trait pairs, and the comparison of blocks with significant local genetic covariance for these three methods using different threshold** A. Local genetic correlations comparison between $\rho$-hess and SUPERGNOVA with Pearson correlation shown (R=0.74).B. Local genetic correlations comparison between LAVA and SUPERGNOVA with Pearson correlation shown (R=0.57). C. Local genetic correlations comparison between $\rho$-hess and LAVA with Pearson correlation shown (R=0.62). D. The Venn diagram of overlapping significant blocks in distinct methods using FDR at 0.1 level. E. using Bonferroni correlated p-value.

#### Supplementary Figure 24

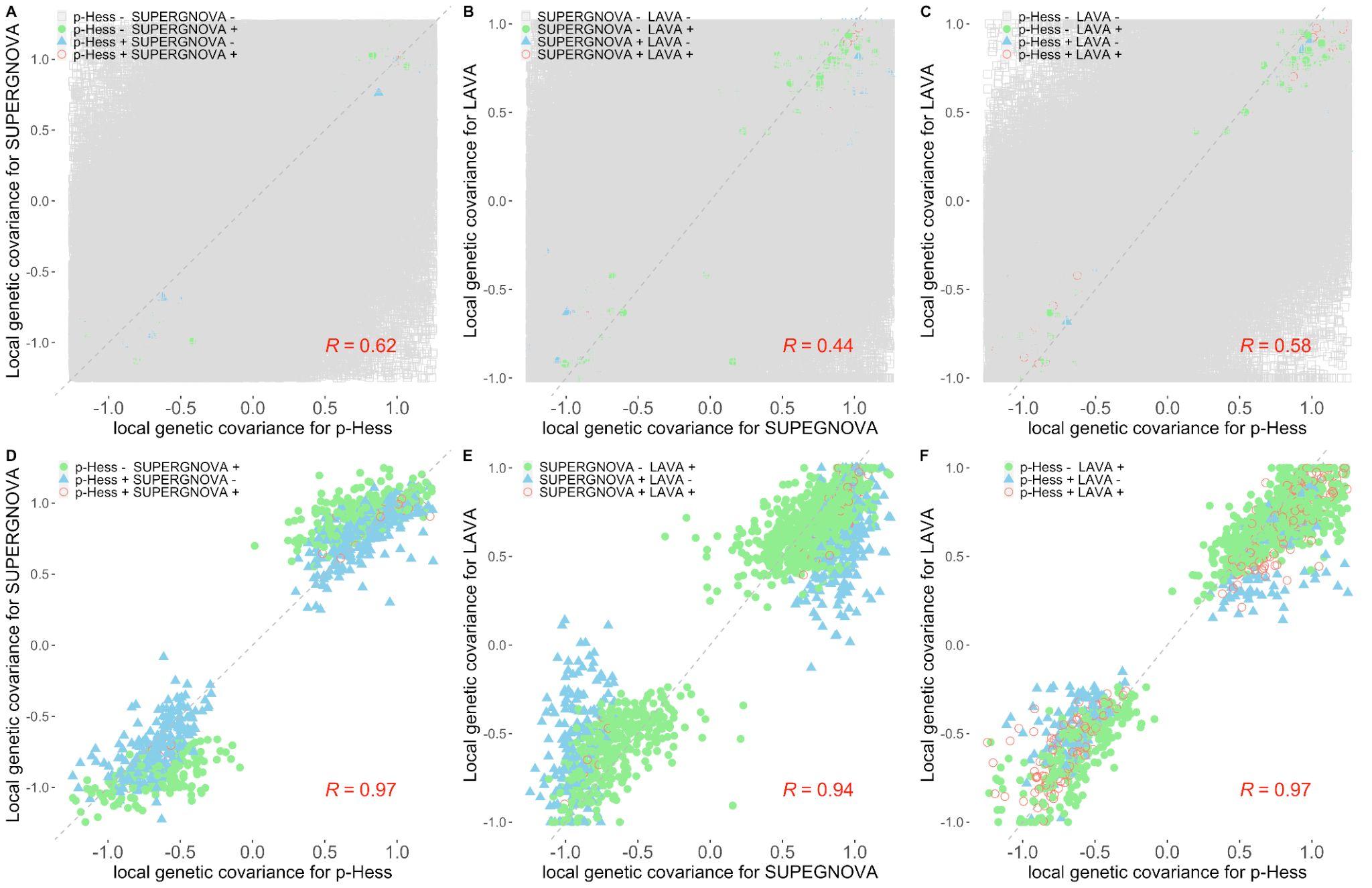

**Supplementary Figure 24: The comparison of local genetic correlations derived from** $\boldsymbol{\rho}$**-Hess, SUPERGNOVA, and LAVA using EUR 1KG reference panels across 2,048 loci for 465 phenotype pairs.** A. Local genetic correlations comparison between $\rho$-hess and SUPERGNOVA with Pearson correlation shown (R=0.62). B. Local genetic correlations comparison between SUPERGNOVA and LAVA with Pearson correlation shown (R=0.44). C. Local genetic correlations comparison between $\rho$-hess and LAVA with Pearson correlation shown (R=0.58). D. Local genetic correlations comparison between $\rho$-hess and SUPERGNOVA when estimates that are not significant for neither method with Pearson correlation shown (R=0.97). E. Local genetic correlations comparison between SUPERGNOVA and LAVA when estimates that are not significant for neither method with Pearson correlation shown (R=0.94). F. Local genetic correlations comparison between $\rho$-hess and LAVA when estimates that are not significant for neither method with Pearson correlation shown (R=0.97).

#### Supplementary Figure 25

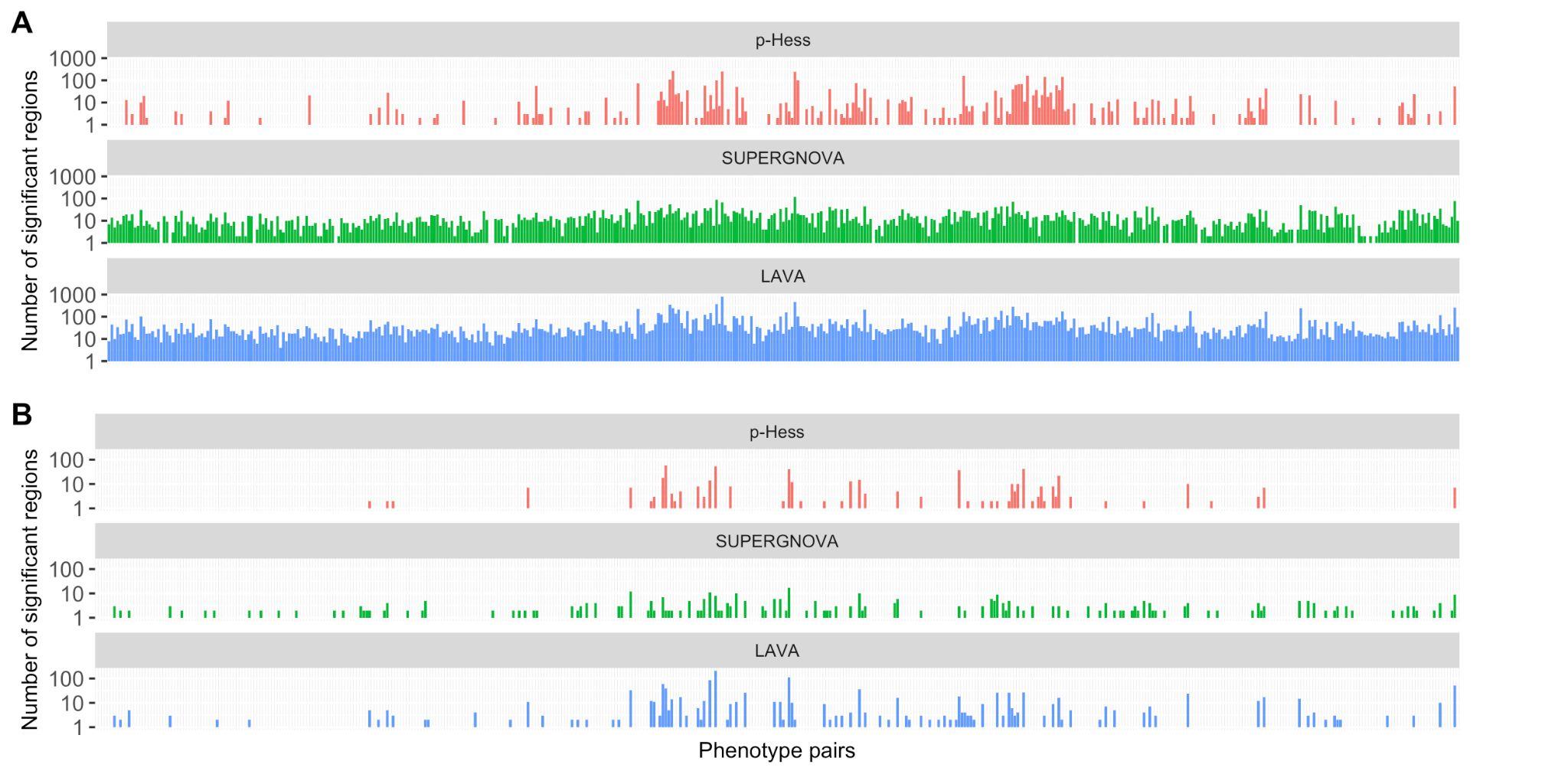

**Supplementary Figure 25: Distribution of the number of significant loci across 465 phenotype pairs detected by each method using different thresholds** A. FDR at 0.1 level and B. Bonferroni correction.

#### Supplementary Figure 26

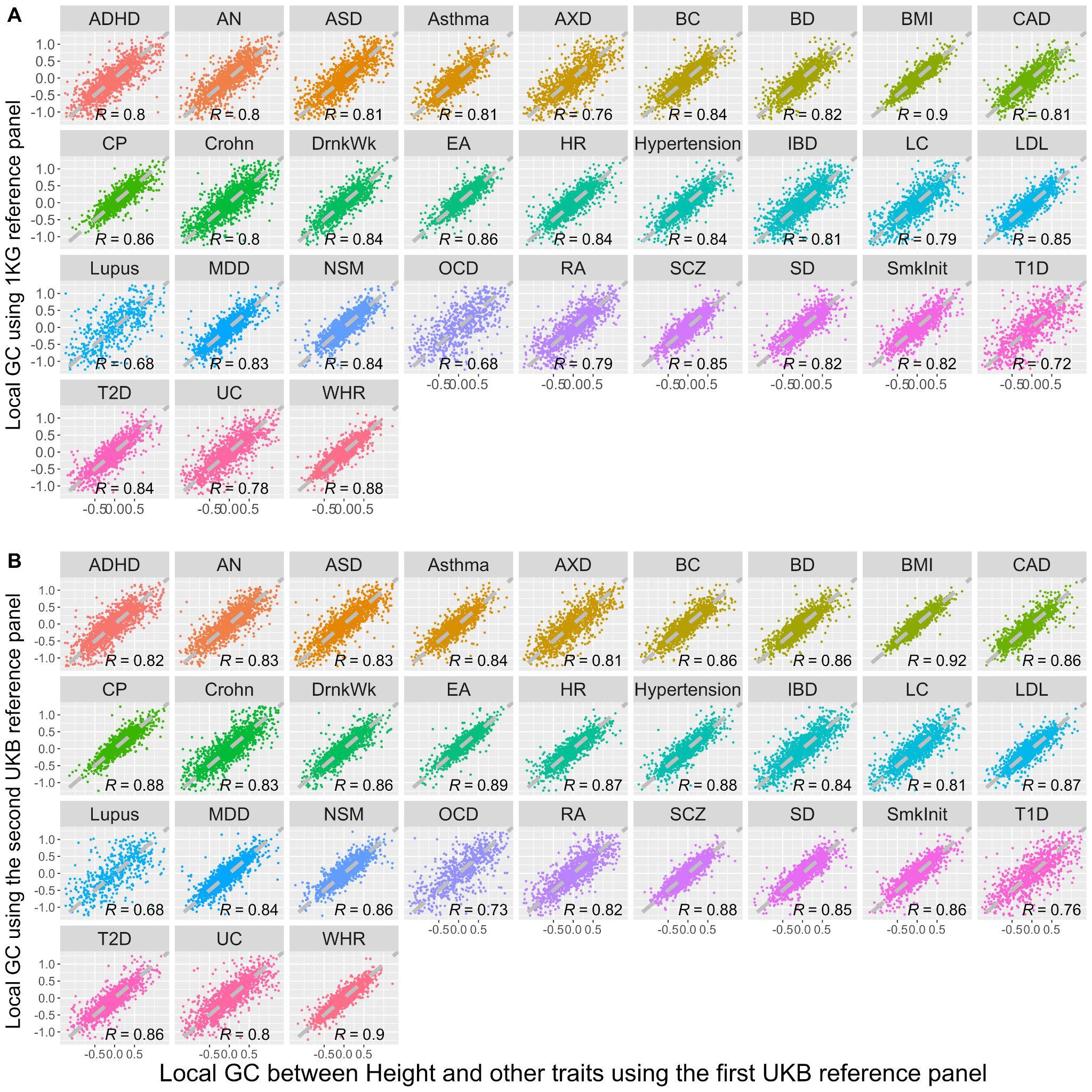

**Supplementary Figure 26: The comparison of local genetic correlation for height and other 30 traits between different reference panels using** $\boldsymbol{\rho}$**-hess.** A. The relationship between local genetic correlation for pairs of traits (height versus others) in certain blocks, as estimated using two different reference panels: a EUR 1KG reference panel and a UKB reference panel. B. The relationship between local genetic correlation for pairs of traits (height versus others) in certain blocks, as estimated using two different UKB reference panels with different samples. The two figures are divided into multiple panels, with each panel corresponding to a different pair of traits. A dashed, grey reference line with a slope of 1 represents the line of perfect correlation in each panel. The strength of the relationship is indicated by Pearson correlation coefficients, which are displayed on the bottom right of each panel.

#### Supplementary Figure 27

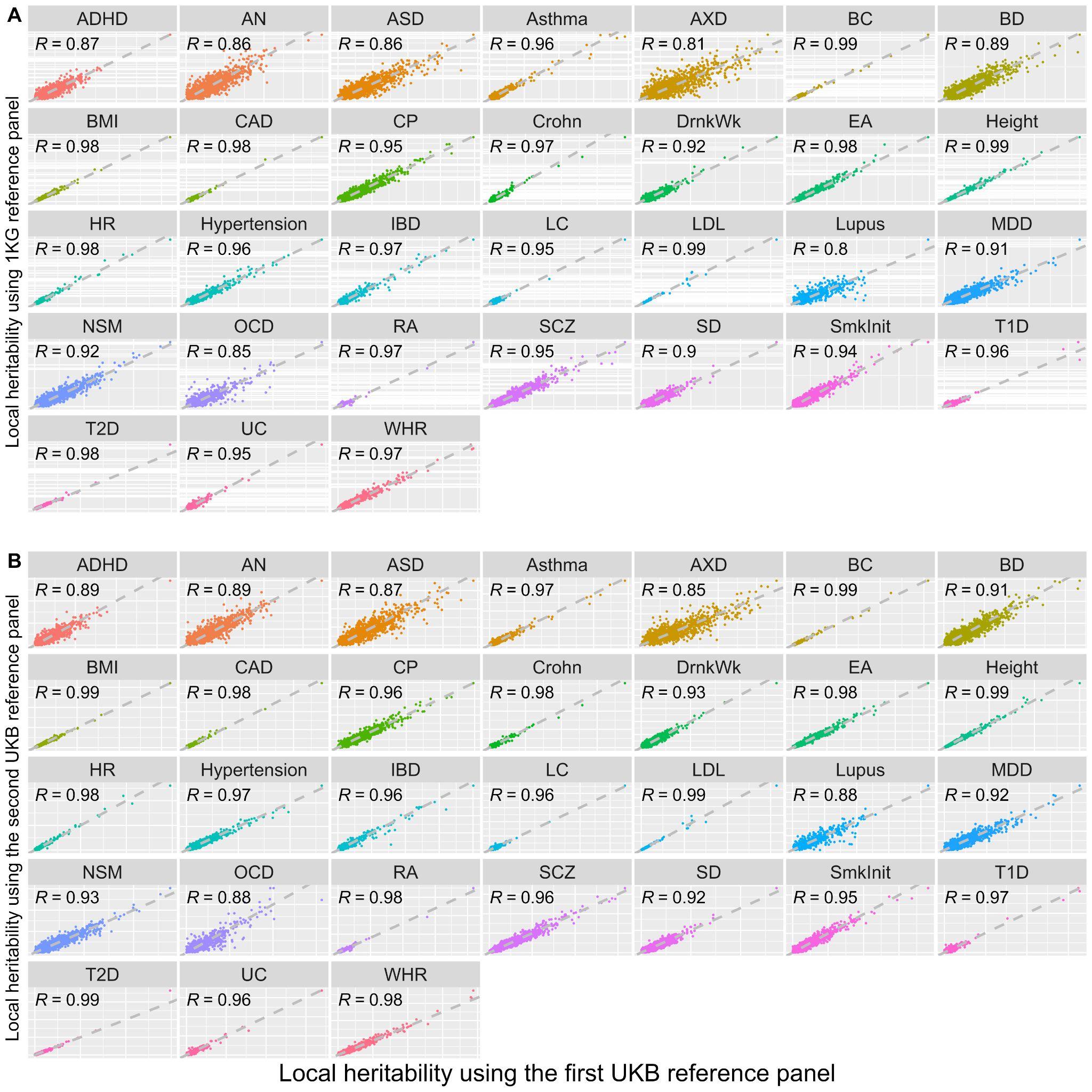

**Supplementary Figure 27: The comparison of local heritability for 30 traits between different reference panels using** $\boldsymbol{\rho}$**-hess.** For each trait, the local heritability was estimated using the overlap SNPs with height GWAS. A. The relationship between local heritability in certain blocks is estimated using two different reference panels: a EUR 1KG reference panel and a UKB reference panel. B. The relationship between local heritability in certain blocks, as estimated using two different UKB reference panels with different samples. The two figures are divided into multiple panels, with each panel corresponding to different traits. A dashed, grey reference line with a slope of 1 represents the line of perfect correlation in each panel. The strength of the relationship is indicated by Pearson correlation coefficients, which are displayed on the bottom right of each panel.

#### Supplementary Figure 28

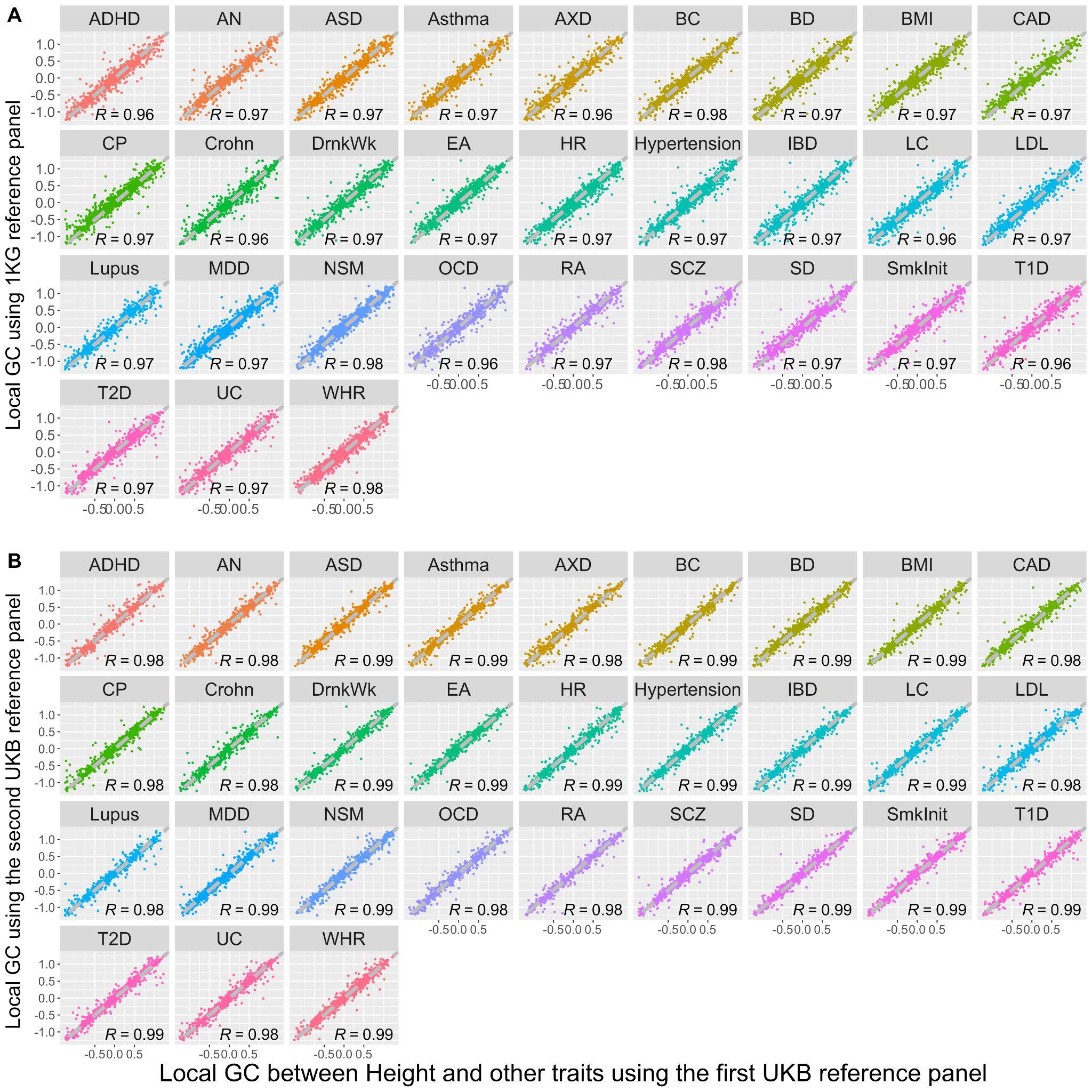

**Supplementary Figure 28: The comparison of local genetic correlation for height and other 30 traits between difference reference panels using SUPERGNOVA.** A. The relationship between local genetic correlation for pairs of traits (height versus others) in certain blocks, as estimated using two different reference panels: a EUR 1KG reference panel and a UKB reference panel. B. The relationship between local genetic correlation for pairs of traits (height versus others) in certain blocks, as estimated using two different UKB reference panels with different samples. The two figures are divided into multiple panels, with each panel corresponding to a different pair of traits. A dashed, grey reference line with a slope of 1 represents the line of perfect correlation in each panel. The strength of the relationship is indicated by Pearson correlation coefficients, which are displayed on the bottom right of each panel.

#### Supplementary Figure 29

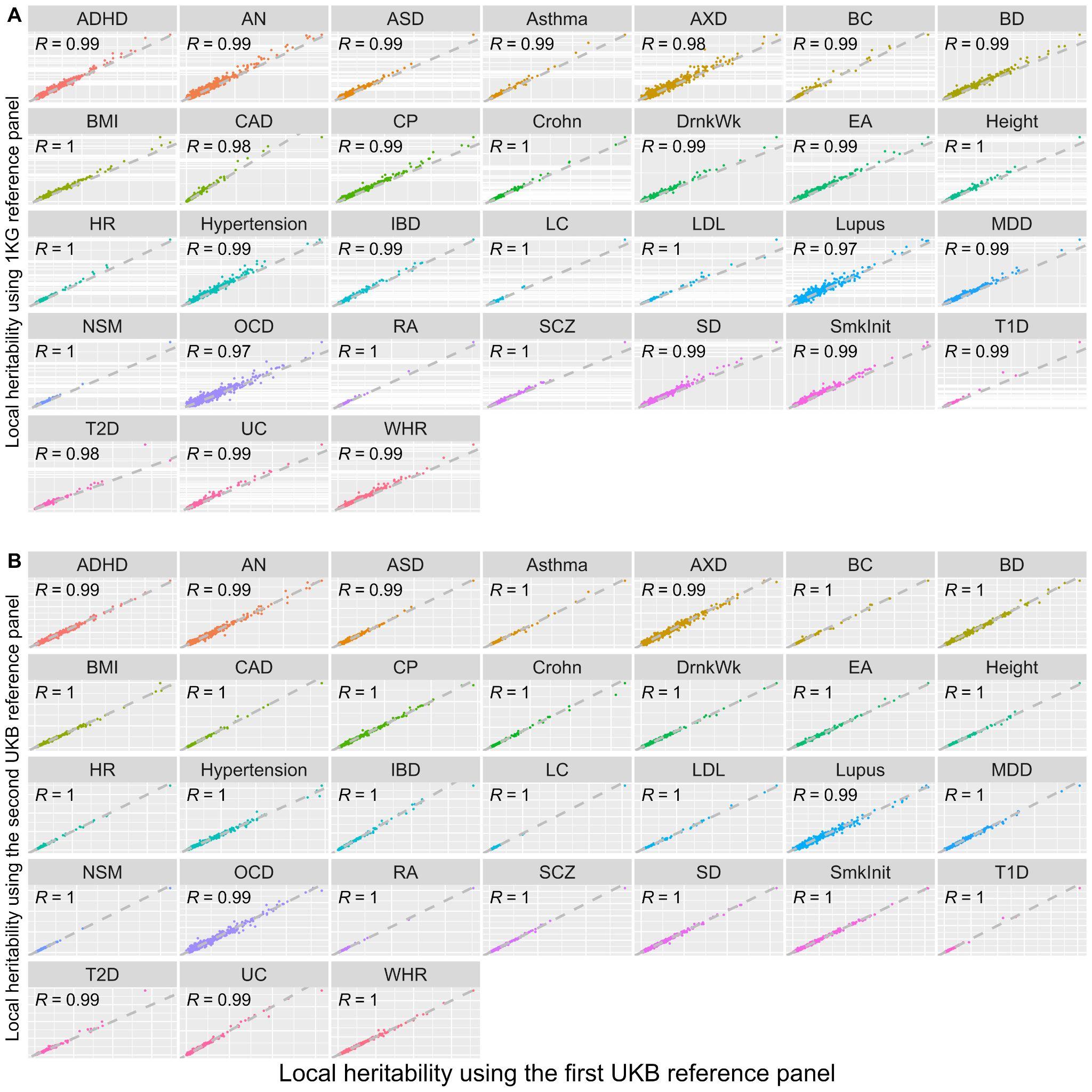

**Supplementary Figure 29: The comparison of local heritability for 30 traits between difference reference panels using SUPERGNOVA.** For each trait, the local heritability was estimated using the overlap SNPs with height GWAS. A. The relationship between local heritability in certain blocks is estimated using two different reference panels: a EUR 1KG reference panel and a UKB reference panel. B. The relationship between local heritability in certain blocks, as estimated using two different UKB reference panels with different samples. The two figures are divided into multiple panels, with each panel corresponding to a different trait. A dashed, grey reference line with a slope of 1 represents the line of perfect correlation in each panel. The strength of the relationship is indicated by Pearson correlation coefficients, which are displayed on the bottom right of each panel.

#### Supplementary Figure 30

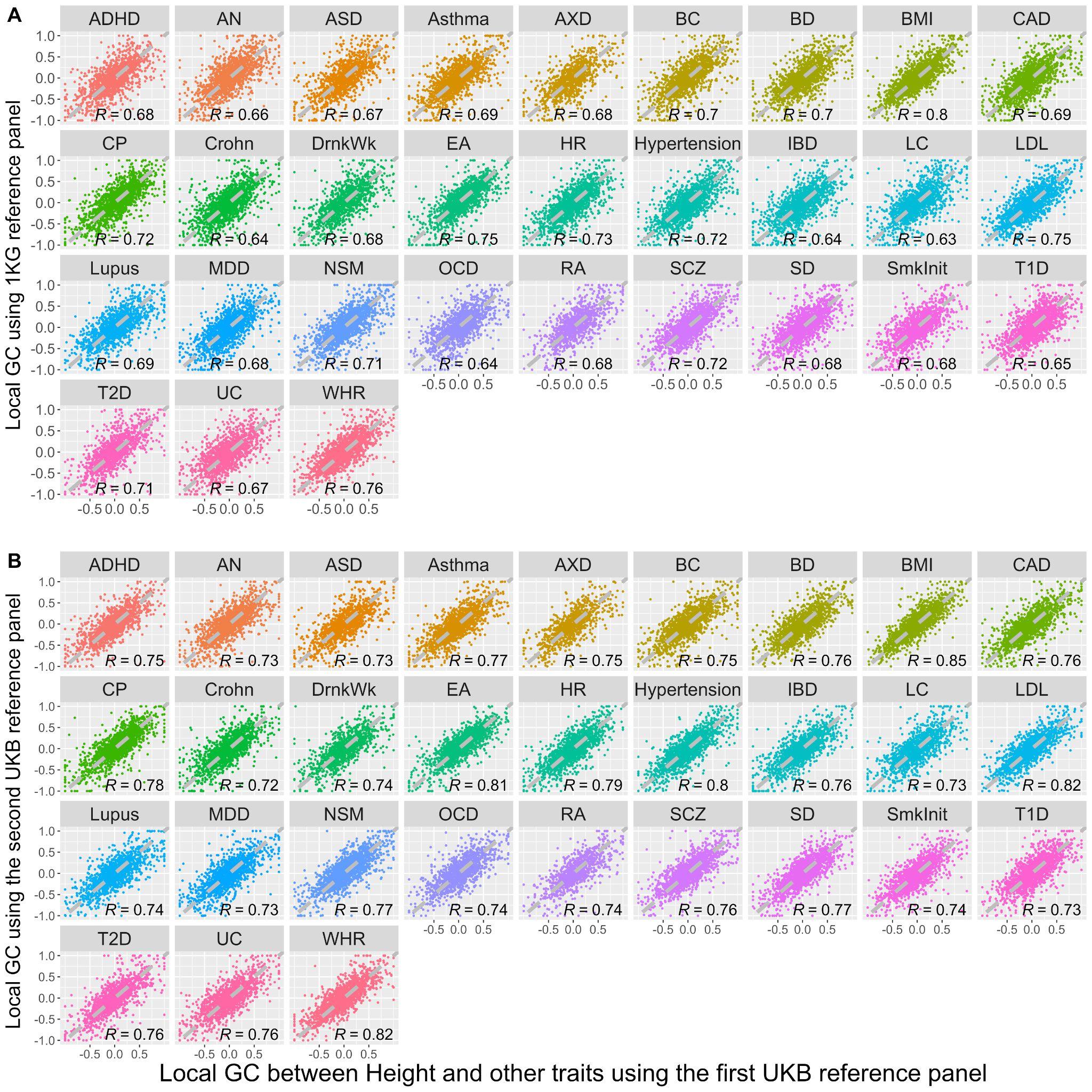

**Supplementary Figure 30: The comparison of local genetic correlation for height and other 30 traits between difference reference panels using LAVA.** A. The relationship between local genetic correlation for pairs of traits (height versus others) in certain blocks, as estimated using two different reference panels: a EUR 1KG reference panel and a UKB reference panel. B. The relationship between local genetic correlation for pairs of traits (height versus others) in certain blocks, as estimated using two different UKB reference panels with different samples.

The two figures are divided into multiple panels, with each panel corresponding to a different pair of traits. A dashed, grey reference line with a slope of 1 represents the line of perfect correlation in each panel. The strength of the relationship is indicated by Pearson correlation coefficients, which are displayed on the bottom right of each panel.

#### Supplementary Figure 31

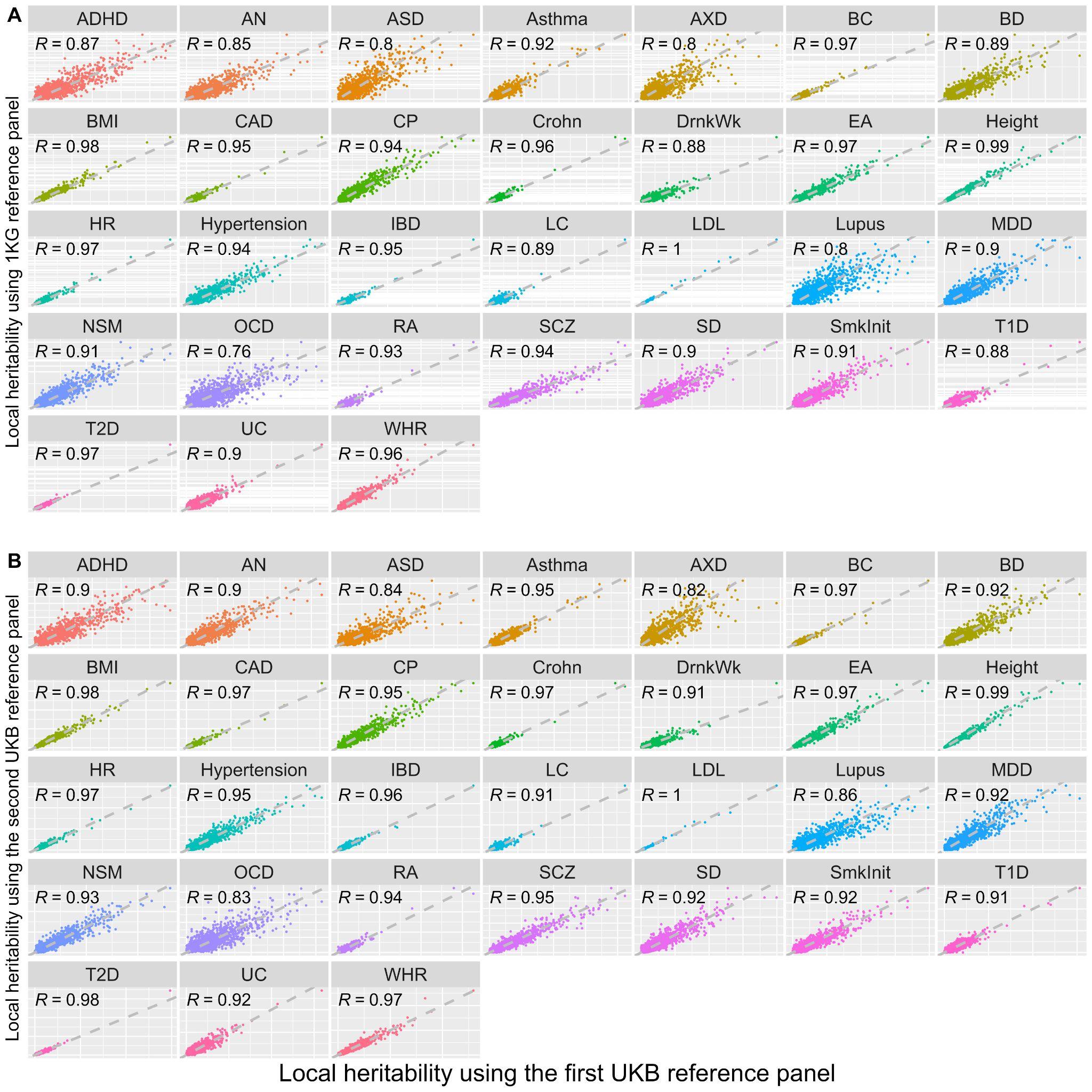

**Supplementary Figure 31: The comparison of local heritability for 30 traits between different reference panels using LAVA.** For each trait, the local heritability was estimated using the overlap SNPs with height GWAS. **A.** The relationship between local heritability in certain blocks is estimated using two different reference panels: a EUR 1KG reference panel and a UKB reference panel. **B.** The relationship between local heritability in certain blocks, as estimated using two different UKB reference panels with different samples.

The two figures are divided into multiple panels, with each panel corresponding to different traits. A dashed, grey reference line with a slope of 1 represents the line of perfect correlation in each panel. The strength of the relationship is indicated by Pearson correlation coefficients, which are displayed on the bottom right of each panel.

#### Supplementary Figure 32

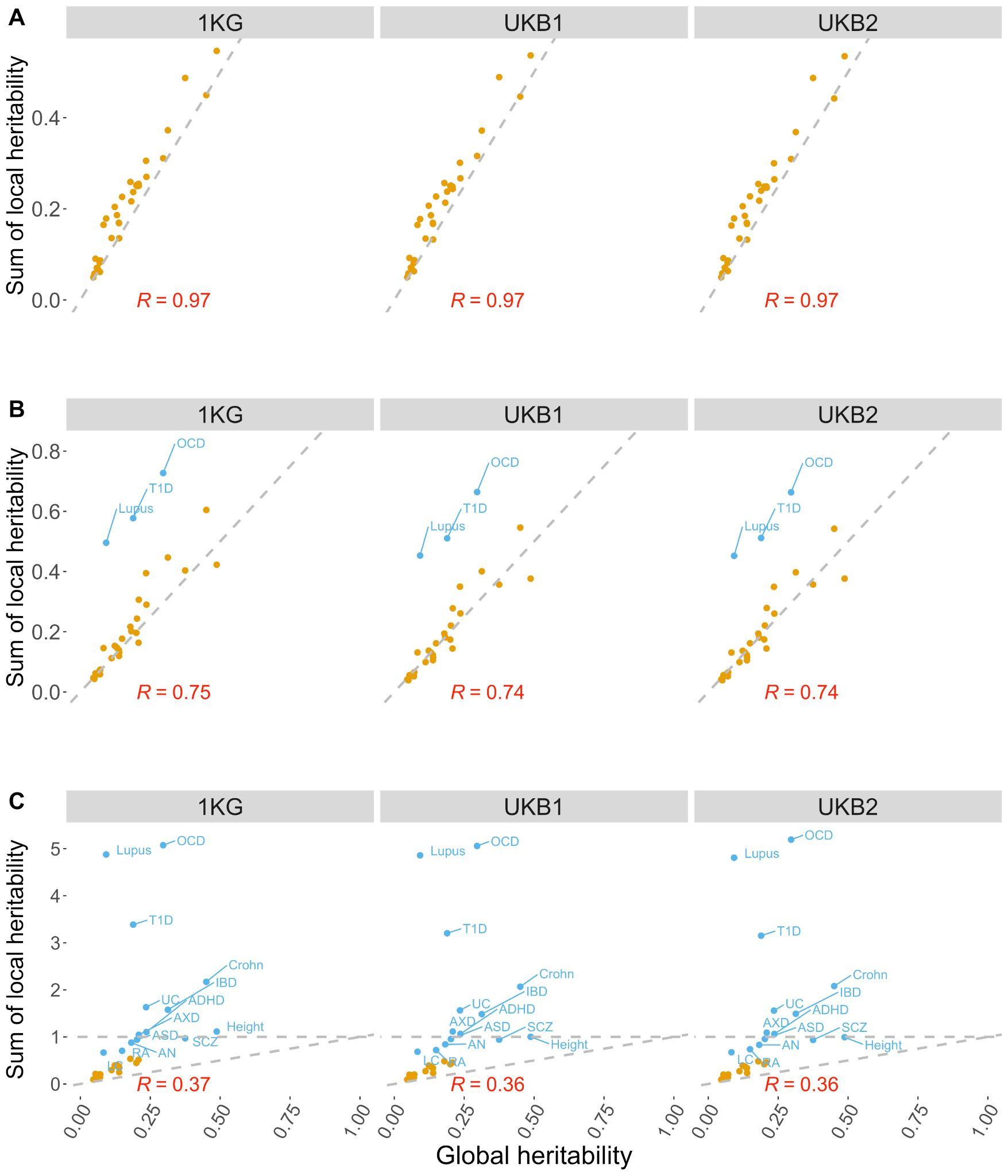

**Supplementary Figure 32: The comparison of the sum of local heritability for 31 traits with the global heritability derived from LDSC. A.** The comparison of the sum of local heritability was evaluated using $\rho$-hess with the global heritability derived from LDSC using three different reference panels. **B.** The comparison of the sum of local heritability was evaluated using SUPERGNOVA with the global heritability derived from LDSC using three different reference panels. **C.** The comparison of the sum of local heritability evaluated using LAVA with the global heritability derived from LDSC using three different reference panels.

The three figures are divided into multiple panels, with each panel corresponding to different reference panels (EUR 1KG reference panel and two UKB reference panels with different samples). A dashed, grey reference line with a slope of 1 represents the line of perfect correlation in each panel and in figure C the dashed and grey reference lines with an intercept of 1 and a slope of 0 are also displayed. The strength of the relationship is indicated by Pearson correlation coefficients, which are displayed at the bottom of each panel.

#### Supplementary Figure 33

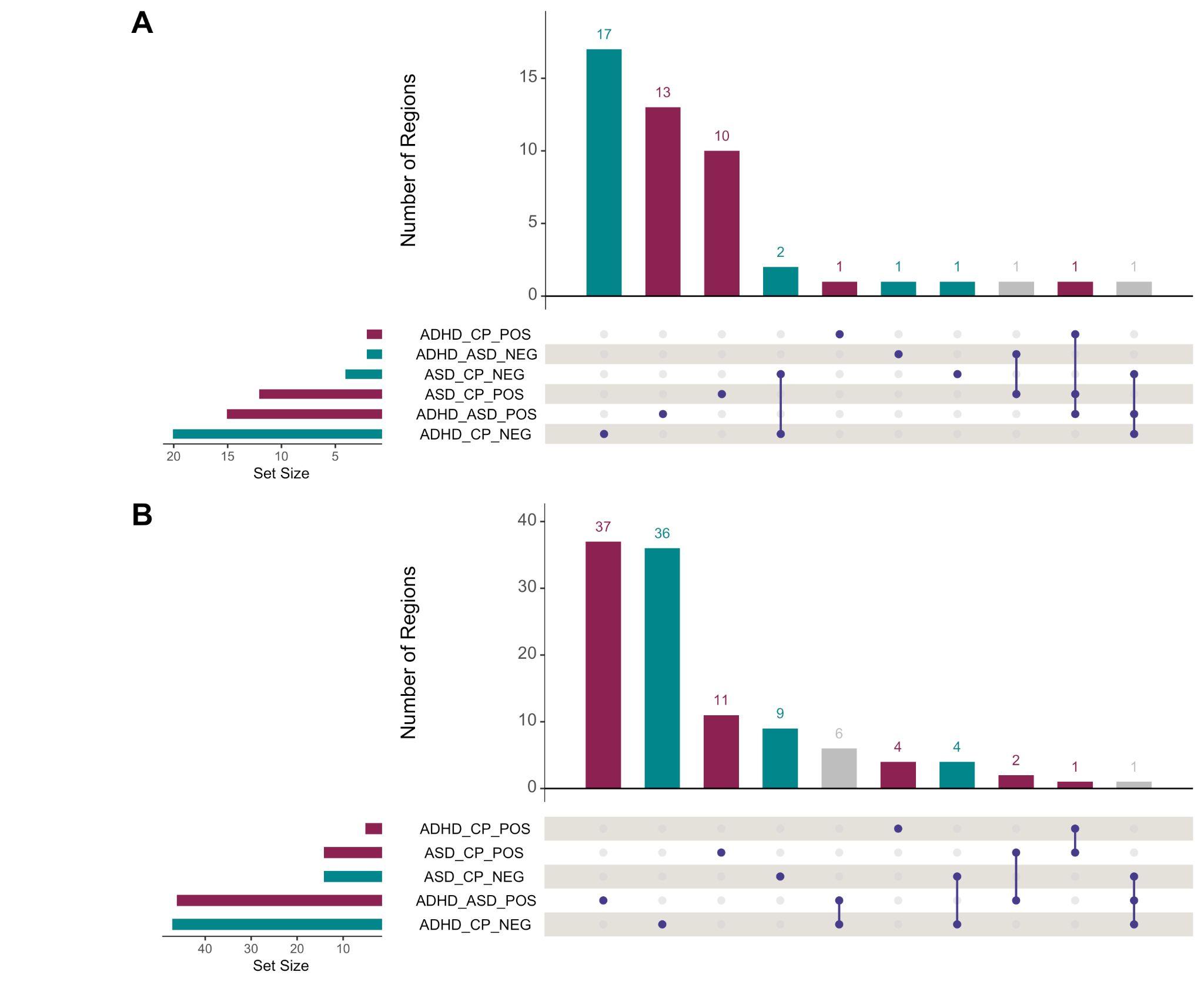

**Supplementary Figure 33: Regions with significant local genetic covariance among ADHD, ASD, and CP (FDR < 0.1) detected by A.** SUPERGNOVA, and **B.** LAVA. The Venn diagram of overlapping regions in distinct categories is decomposed using bars in these figures. The five categories in the lower panel represent detected significant loci of ADHD and CP (positive and negative), ASD and CP (positive and negative), and ASD and ADHD (positive and negative). We use red to annotate positively correlated regions, blue to annotate negatively correlated regions and gray to annotate regions with both positive and negative correlations.

#### Supplementary Figure 34

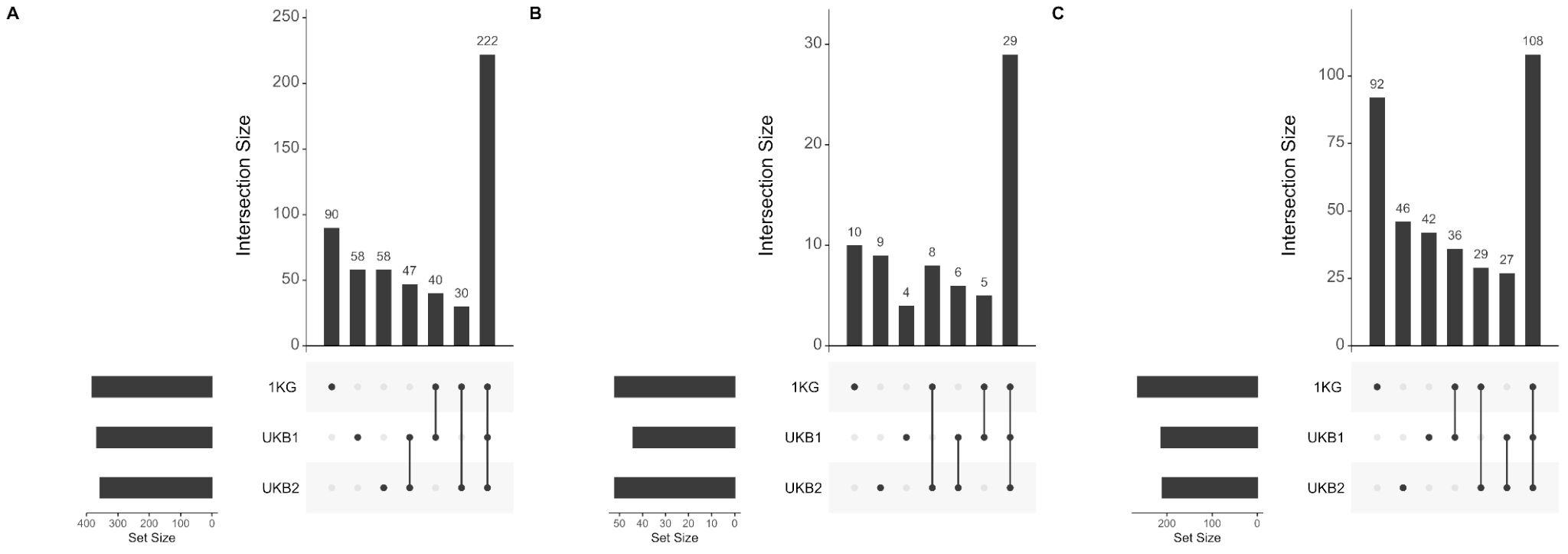

**Supplementary Figure 34: Comparisons of blocks with significant local genetic correlations when using different reference panels.** These plots used bars to break down the Venn diagram of overlapped significant blocks using different reference panels using Bonferroni corrected p-value detected by A. $\rho$-hess, B. SUPERGNOVA, and C. LAVA.

### Appendix A

We calculated the sum of squared correlations between SNPs at different blocks, referred to as the ‘cost’ using the compute_cost function from the bigsnpr package [1] on the quality-controlled EUR 1KG reference panel for each chromosome. We then applied the snp_ldsplit function to the same 1KG reference panel and compared the cost of partitions from SUPERGNOVA, LDetect, and LAVA with the cost of the partition from snp_ldsplit. As shown in Supplementary Figure 1, the partition produced by snp_ldsplit had a lower cost compared to those produced by LDetect, SUPERGNOVA, and LAVA, for the respective numbers of blocks in all the 22 chromosomes.

### Appendix B

We used LDSC, SUPERGNOVA, LAVA, and $\rho$-hess to compute the genetic correlation and genetic covariance between pairs of 31 complex traits first using the European 1KG Phase3 data as the LD reference panel. LDSC was used to obtain global genetic correlations and their significance. Using 2,048 blocks partitioned by the snp_ldsplit function, SUPERGNOVA had results for 950,925 comparisons, LAVA had 608,692 comparisons, and $\rho$-hess had 952,320 comparisons. While both LAVA and $\rho$-hess provided p-values for local heritability, SUPERGNOVA only provided values for local heritability. To ensure a fair comparison, we selected blocks with local heritability greater than zero for further analysis. This resulted in 520,133 comparisons for SUPERGNOVA, 605,392 comparisons for LAVA, and 681,153 comparisons for $\rho$-hess. Furthermore, we removed blocks whose estimated local genetic correlations were smaller than -1.25 or larger than 1.25 in SUPERGNOVA and $\rho$-hess because in default LAVA considered such estimates to be unreliable and set them to missing values. After that, the number of remaining comparisons was 541,211 for $\rho$-hess, 413,062 for SUPERGNOVA, and 681,153 for LAVA.

With a Bonferroni-corrected P-value threshold of 9.24e-8 (0.05/541,211), 1.21e-7 (0.05/413,062), and 8.26e-8 (0.05/605,392), we detected 539, 549, and 1,294 significant correlations across 279, 153, and 419 blocks using $\rho$-hess, SUPERGNOVA, and LAVA, respectively (Supplementary Table 5-7). Among 279 blocks detected by $\rho$-hess, 98 blocks were associated with more than one trait pair and 93 trait pairs had at least one block with significant correlations. Among 153 blocks detected by SUPERGNOVA, 79 blocks were associated with more than one trait pair and 245 trait pairs had at least one block with significant correlations. Among 1,294 blocks detected by LAVA, 242 blocks were associated with more than one trait pair and 172 trait pairs had at least one block with significant correlations.

We evaluated the local genetic correlations for these 465 trait pairs by computing the average of their estimated local genetic correlations across the remaining blocks selected above and compared them with the global genetic correlations (Supplementary Figures 20-22). The results revealed consistent, significant positive local genetic correlations for 21 trait pairs using all three methods, each of which also exhibited significant positive global correlations. There were also seven trait pairs, such as ADHD and EA, CP and SCZ, MDD and NSM, which exhibited both significant negative global genetic correlation and consistent, significant negative local genetic correlations for all three methods.

We also observed that for each method, there were some trait pairs that had both positively correlated blocks and negatively correlated blocks. For these trait pairs, there were more or equal numbers of significant positive blocks than negative blocks when the global correlation was positive, and vice versa when the global correlation was negative. Furthermore, we identified 27 trait pairs, for example, Asthma and IBD, Height and IBD, and DrnkWk and LDL, that did not show significant global genetic correlation, but had significant local genetic correlations for all three methods. These results suggest that only considering global genetic correlation may fail to capture heterogeneous genetic correlations and highlight the importance of considering local genetic correlations.

We compared the local genetic covariances estimated in the same block for the same trait pair among the three methods and found that $\rho$-hess and SUPERGNOVA had the highest correlation (R=0.74) whereas SUPERGNOVA and LAVA had the lowest correlation (R=0.57) (Supplementary Figures 23). We also compared the local genetic correlation displayed in Supplementary Figure 24. When all the estimates were considered, the Pearson correlation among the three methods was relatively low, ranging from 0.44 to 0.62. However, when we excluded estimates that did not show significance in both compared methods, the correlation increased significantly, ranging from 0.94 to 0.97. We also compared the number of significant blocks identified by the three methods under two threshold settings(Supplementary Figures 23). Regardless of the threshold used, we found that LAVA consistently identified more significant blocks than both SUPERGNOVA and $\rho$-hess. Among the three methods, LAVA and $\rho$-hess had the greatest overlap, however, even this overlap was much smaller than the number of blocks identified exclusively by LAVA. Additionally, we found that only a small proportion of significant blocks were detected by all three methods. Supplementary Figure 25 illustrates the distribution of the significant blocks detected by these three methods across all the trait pairs. Along with Supplementary Figure 20-22, it can be observed that $\rho$-hess detected significant correlations in fewer trait pairs compared to LAVA and SUPERGNOVA. However, SUPERGNOVA tended to detect fewer significant blocks for trait pairs but found more trait pairs with significant correlations.
